# Trends in medication use after the onset of the COVID-19 pandemic in the Republic of Ireland: an interrupted time series study

**DOI:** 10.1101/2023.06.09.23291202

**Authors:** Molly Mattsson, Jung Ah Hong, John Scott Frazer, Glenn Ross Frazer, Frank Moriarty

**Affiliations:** School of Pharmacy and Biomolecular Sciences, RCSI University of Medicine and Health Sciences, Dublin, Ireland; Somerville College, University of Oxford, Oxford, UK; Unaffiliated

**Keywords:** prescribing, trends, interrupted time series, COVID-19 pandemic

## Abstract

The COVID-19 pandemic had a substantial impact on healthcare delivery, particularly in general practice. This study aimed to evaluate how dispensing of medications in primary care in Ireland changed following the COVID-19 pandemic’s onset compared to expected trends. This interrupted time series study used data on medications prescribed in general practice 2016-2022 to patient eligible for state health cover, approximately one third of the population. Dispensing volumes for all therapeutic subgroups (ATC2 codes) and commonly dispensed medications were summarised. Pre-pandemic data was used to forecast expected trends (with 99% prediction intervals) using the Holt-Winters method, and these were compared to observed dispensing from March 2020 onwards. Most (31/77) therapeutic subgroups had dispensing significantly different from forecast in March 2020. Drugs for obstructive airway disease had the largest difference, with dispensing 26.2% (99%CI 19.5%-33.6%) higher than forecasted. Only two subgroups were significantly lower than forecasted, other gynaecologicals (17.7% lower, 99%CI 6.3%-26.6%) and dressings (11.6%, 99%CI 9.4%-41.6%). Dispensing of amoxicillin products and oral prednisolone were lower than forecasted in the months following the pandemic’s onset, particularly during winter 2020/2021. There was a spike in dispensing for many long-term medications in March 2020, while pandemic restrictions likely contributed to reductions for other medications.

## Introduction

The onset of the COVID-19 pandemic represented a substantial change to people’s lives, affecting all aspects of society, including the delivery of healthcare. In the Republic of Ireland, the first restrictions were introduced from March 12^th^ 2020, with the first “lockdown” introduced as of March 27^th^. The pandemic prompted changes in primary care, with a move towards telemedicine, affecting the processes of medication prescribing, dispensing, and monitoring.[1, 2] Patients also reported challenges in accessing healthcare services.[3]

The onset of the pandemic and restrictions across a variety of countries were followed by changes in how medications were prescribed or dispensed. Evidence from England showed lower than expected prescribing of some medications in March 2020, including oral steroids, contraceptive pills, travel vaccines, medications for dementia or Parkinson’s disease, and other medications that required face-to-face visits.[4] A US study on chronic medications from classes with significant population health impact, found that more prescriptions were filled in March 2020 than in any prior month, however this was followed by a significant drop in monthly dispensing and an increased likelihood of medication discontinuation.[5] In contrast in England, there was higher than expected dispensing of respiratory inhalers including corticosteroids, all types of insulins and diabetic related expendables, and immunomodulators, suggesting an increased immediate demand or precautionary stockpiling.[4] Furthermore, medications such as benzodiazepines, opioids, and analgesics used in end-of-life care showed increases in prescribing in April 2020, potentially indicating a peak of palliative care and death related to COVID-19.[4] A study of prescribing in nursing homes found an increase in drugs used at end of life, however the largest increases were seen in prescribing of antidepressants and antipsychotics.[6] Another study across eight countries in Europe, analysing all prescribing medicines based on the Anatomical Therapeutic Chemical (ATC) Classification, showed decreases in the antibiotic and nasal preparations dispensing, potentially due to lockdowns and social distancing resulting in lower chances of transmission of respiratory infections. This study also identified increased dispensing of some medications, again proposed as potential stockpiling to reduce the need to visit healthcare professionals or to avoid medication shortages.[7] To date, how medication use changed in the Republic of Ireland following the onset of the pandemic has yet to be evaluated.

Therefore, the aim of this study was to evaluate how dispensing of medications in primary care in the Republic of Ireland changed following the onset of the COVID-19 pandemic, and how this differed from expected trends in use.

## Methods

This was an interrupted time series study, and the protocol was pre-registered on OSF.[8] It used data from the Health Service Executive Primary Care Reimbursement Service (PCRS) in the Republic of Ireland. The PCRS publishes monthly data on the number of dispensings and costs of the top 100 individual medications, as well as by WHO Anatomical Therapeutic Chemical (ATC) classification code at the second level, i.e. indicating the pharmacological or therapeutic subgroup, of which there are 94, and 87 were included in PCRS data during the study period (including six subgroups within V07 “All Other Non-Therapeutic Products”). These data are published separately for different state drug schemes. For this study, we used data from the General Medical Services (GMS) scheme. Eligibility for this scheme is means tested and covers approximately 32% of the population, and therefore eligible persons tend to be more socioeconomically deprived than the general population.[9, 10] For adults aged 70 years and over, the scheme covers the vast majority of adults.

We included data from January 2016 (the earliest date for which PCRS have published monthly data at the time of writing) to July 2022. A cyberattack on the Republic of Ireland’s health service in May 2021 disrupted transmission of pharmacy claims for that month, resulting in a substantial drop in apparent dispensings, followed by a substantial increase for June 2021. The mean of dispensing for these two months was calculated and assigned to each month. The specific time point of interest is March 2020 (the initial onset of the COVID-19 pandemic restrictions in the Republic of Ireland). Data used in this study is available from https://doi.org/10.5281/zenodo.7999791.

As the primary outcome, we examined the number of dispensings in each pharmacological/therapeutic subgroup, and as the secondary outcome, the number of dispensings of the most common individual medications. In cases where data was reported for at least two individual medication within a therapeutic subgroup (e.g. atorvastatin and simvastatin within lipid-lowering drugs), we analysed the individual medications, and the residual dispensing within the subgroup (i.e. atorvastatin, simvastatin, and other lipid-lowering agents). Individual medications which did not remain in the top 100 for the study period up to March 2020 (i.e. where data on the quantity dispensed is missing for some months) were not considered. Of 133 individual medications in the top 100 at any point during the study period, 53 were excluded on this basis leaving 80 individual medications. Similarly, therapeutic subgroups were excluded if they were not dispensed in any months during the study period up to March 2020 (seven subgroups), or their frequency of dispensing remained <50 per month for this period (three subgroups), leaving 77 subgroups. The outcomes focused on the frequency of dispensing i.e. the number of times an individual medication or medication within a therapeutic subgroup was dispensed in that calendar month.

### Analysis

Frequency of dispensing of each therapeutic subgroup and included medication were plotted over the study period. Data was grouped into pre-pandemic data (January 2016 to November 2019), used for forecasting, and pandemic data (December 2019 onwards) which the forecast was compared against. Three months of data before the pandemic onset in the Republic of Ireland were included in the “pandemic data” to allow the forecasting model to be validated against data unaffected by the COVID-19 pandemic.

The time series was decomposed into seasonal, trend, and irregular components using locally estimated scatterplot smoothing to allow patterns to be identified. Forecasting was performed using the Holt-Winters method, a predictive technique that uses triple exponential smoothing to apply moving averages to observed data. Triple exponential smoothing amalgamates three separate smoothing methods: Simple Exponential Smoothing assumes that the data have no trend, producing a recursive average used as a baseline value to predict the next point; Holt’s Exponential Smoothing builds on the previous technique but also incorporates a trend component; Winter’s Exponential Smoothing includes a seasonal trend, which captures repeating time-based variation. These three components were weighted and combined, with exponentially decreasing weight given to older data points.

The primary analysis was estimating the magnitude of difference between forecasted and actual data for March 2020 using 99% forecast prediction intervals calculated via the R forecast function as described by Hyndman and Khandakar.[11] The secondary analysis consisted of qualitative comparisons between forecasts and observed data by visual inspection of plots.

## Results

We focus on dispensed prescriptions for therapeutic subgroups and individual medications as percentage differences relative to forecast estimates in March 2020, as the first month of pandemic restrictions in Ireland. Through visual inspections, forecasts for the three months preceding March 2020 were found to match observed dispensings in all cases except for ferrous fumarate and other anti-anaemic preparations, and consequently results for these are not considered further as they are not a reliable estimate of the impact of the COVID-19 pandemic on dispensing.

We included 77 ATC second level pharmacological or therapeutic subgroups, and 80 common individual medications. Fifteen subgroups had more than one individual medication included, and so the remaining prescribing in these subgroups were also analysed alongside the individual medications. Total dispensing increased between 2016 and 2021, with 58.1 million dispensings in 2016 and 62 million in 2021. Overall dispensing was higher than forecast in March 2020, with an increase of 7.6% (99%CI 2.5% to 13.2%).

### Therapeutic subgroups

In total, 31 of 77 subgroups had dispensing significantly different from forecast in March 2020 (see Supplementary table 1 and Supplementary figure 1). The subgroup with the largest absolute as well as percentage difference relative to forecast data was Drugs for Obstructive Airway Diseases (ATC R03). For March 2020, dispensing was 26.2% (99%CI 19.5% to 33.6%) higher than forecasted, corresponding to approximately 77,000 additional dispensings (see Figure 1). Other groups with large absolute increases, as well as statistically significant relative differences, include minerals (A12, 11.3%, 99%CI 6.7% to 16.4%), analgesics (N02, 10.9% (99%CI 4.7% to 17.7%), thyroid therapy (H03, 9.6%, 99%CI 5.0% to 14.6%), serum lipid-reducing agents (C10, 9.2%, 99%CI 4.9% to 13.9%), and diuretics (C03, 9.0%, 99%CI 2.9% to 15.8%).

**Figure 1.**
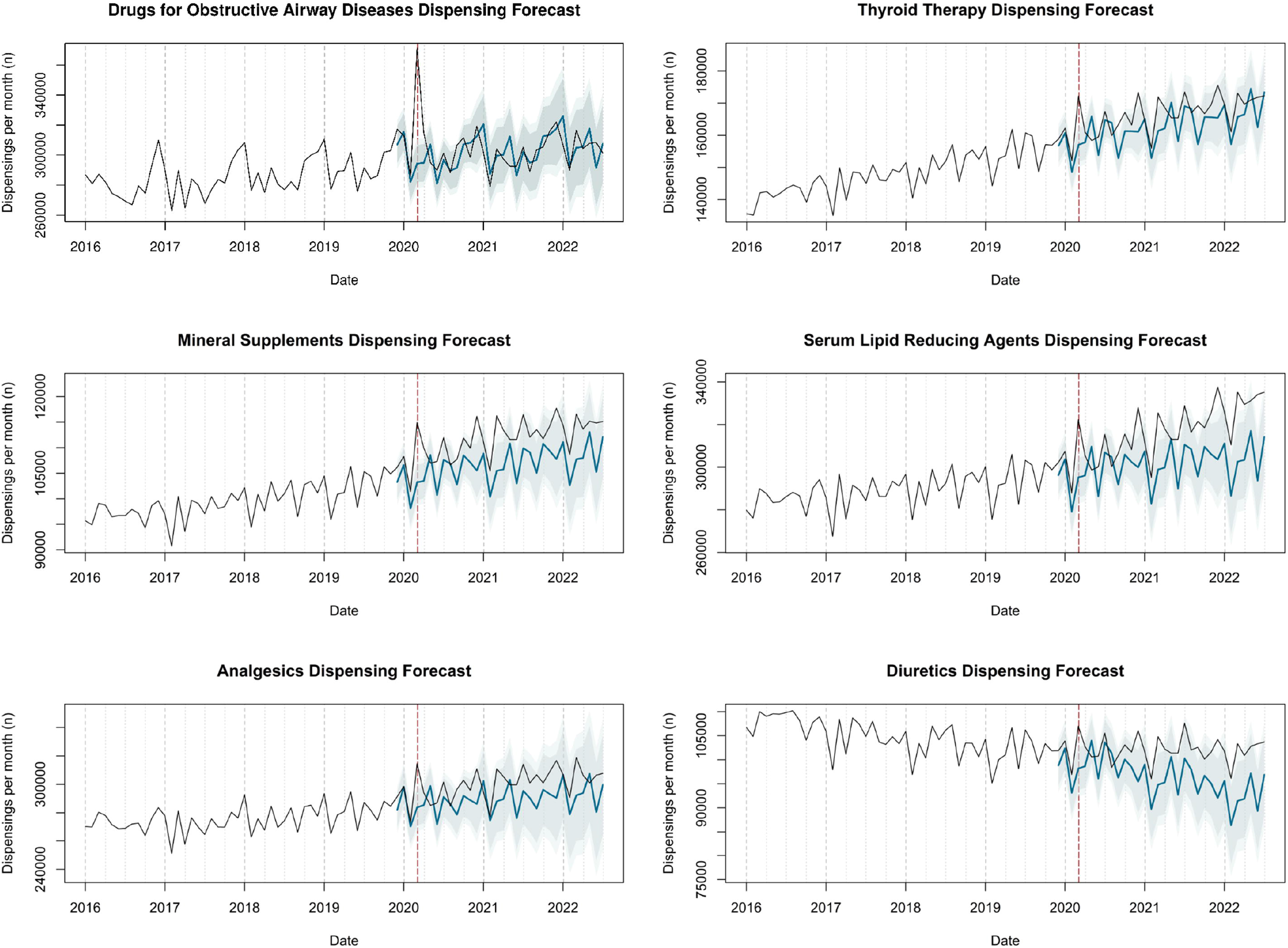
Actual dispensing (black) and forecasted dispensing (blue, with 95% and 99% prediction intervals) for therapeutics subgroups with the largest relative and absolute increases in dispensings compared to forecast in March 2020

Additionally, some therapeutic sub groups saw large relative differences, however in absolute terms the increase in dispensings was small (i.e. less than 1,000 items per month). These include several other non-therapeutic agents (ostomy requisites, urinary requisites and miscellaneous, ranging from 11.6% to 16.6% higher), and antimycobacterials (J04 15.3%, 99%CI 0.7% to 34.8%).

Two therapeutic subgroups saw significant decreases in prescribing (Figure 2); other gynaecologicals (G02, 17.7%, 99%CI 6.3% to 26.6%) and dressings (V07-04 11.6%, 99%CI 9.4% to 41.6%), however the absolute numbers of prescriptions were small (1,961 and 2,632 in March 2020 respectively). This divergence from forecasted dispensing persisted for April and May 2020.

**Figure 2.**
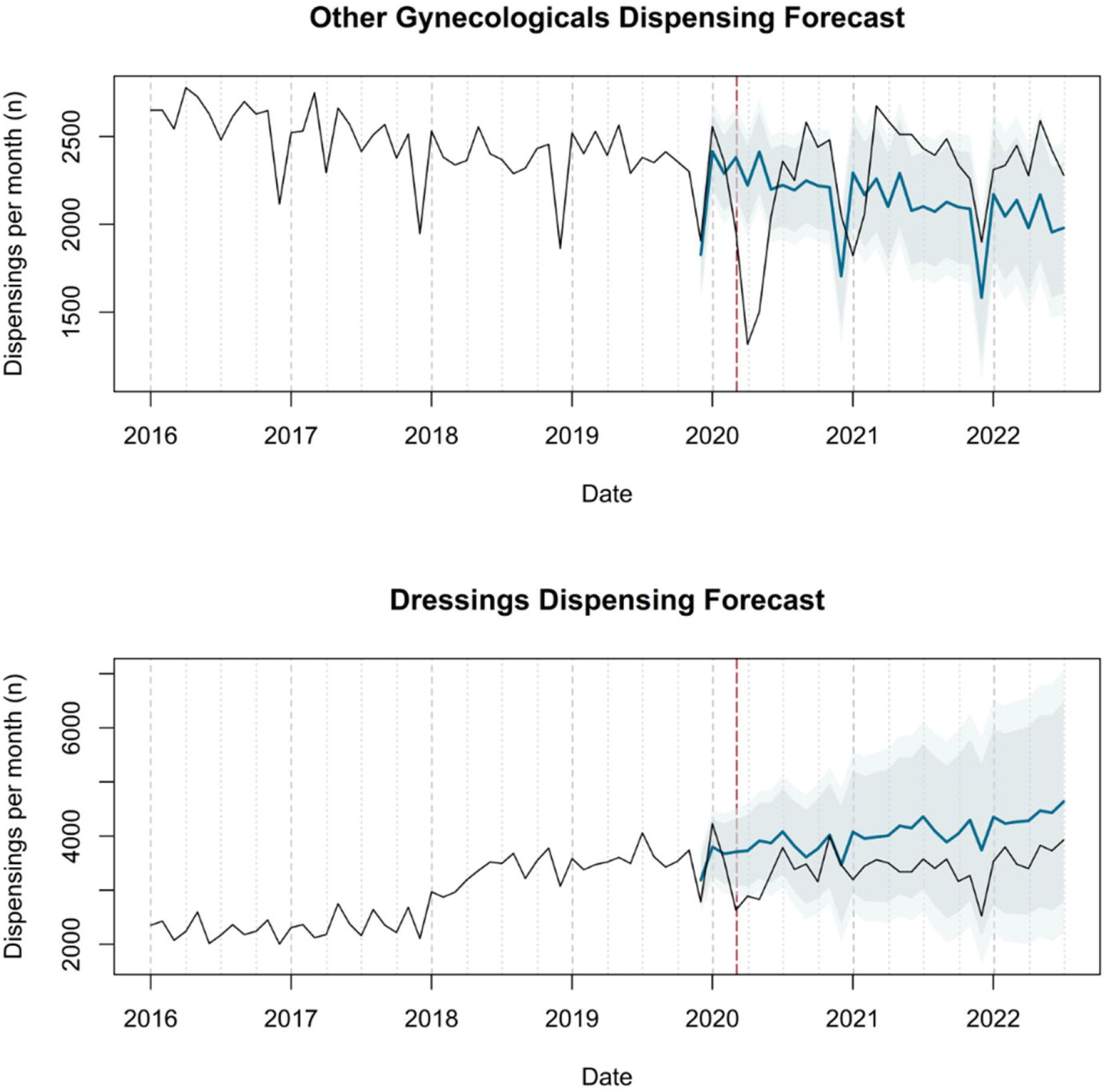
Actual dispensing (black) and forecasted dispensing (blue, with 95% and 99% prediction intervals) for therapeutic subgroups with dispensing lower than forecast in March 2020

### Common individual medications

#### Alimentary tract and metabolism

All common individual medications used for acid-related disorders had dispensing between 8 and 10% higher than forecasted, including alginic acid, esomeprazole, lansoprazole, omeprazole, and pantoprazole. Similar results were found for the laxative lactulose (6.9%, 99%CI 0.3% to 14.4%), the diabetes medication metformin (11.1%, 99%CI 1.0% to 23.6%), and the supplement calcium carbonate and colecalciferol (11.4%, 99%CI 6.7% to 16.4%). No significant difference was seen for colecalciferol prescribed on its own.

#### Blood and blood forming organs

In this group, dispensing was higher than forecast for acetylsalicylic acid (8.1%, 99%CI 3.0% to 13.7%), however levels remained unchanged for folic acid, and the anti-thrombotic agents, rivaroxaban, clopidogrel, and warfarin

#### Cardiovascular system

Among 16 individual medications affecting the cardiovascular system that were included, 14 had significantly higher than forecasted dispensing in March 2020. This included one diuretic, furosemide (10.7%, 99%CI 4.3% to 17.9%), three beta-blockers (atenolol, bisoprolol, and nebivolol with increases between 7.5 and 13.5%), two calcium channel blockers (amlodipine and lercanidipine increasing 6 to 7%), four angiotensin receptor blockers (losartan, olmesartan, perindopril, ramipril, increasing 6 to 9%), and four statins (atorvastatin, pravastatin, rosuvastatin, simvastatin, increasing 8.5 to 10%). Dispensing of two of 16 drugs did not differ significantly from the forecast; the antihypertensive doxazosin and the angiotensin receptor blocker valsartan.

#### Genitourinary system, sex hormones and systemic hormonal preparations

In these groups, three out of four medications saw significant increases, including the combined oral contraceptive levonorgestrel and ethinylestradiol (9.8%, 99%CI 3.0% to 17.6%), benign prostatic hyperplasia medication tamsulosin (8.0%, 99%CI 3.4% to 13.0%), and hypothyroid medication levothyroxine (9.6%, 99%CI 5.0% to 14.6%). Systemic prednisolone did not differ significantly from the forecasted dispensing in March 2020.

#### Musculoskeletal system

Antigout medication allopurinol (9.9%, 99%CI 5.0% to 15.3%) and alendronic acid for osteoporosis (9.0%, 99%CI 2.2% to 16.6%) were both higher than forecast in March 2020. Anti-inflammatories diclofenac and ibuprofen, as well as topical anti-inflammatories diclofenac and etofenamate did not differ significantly from the forecast. Naproxen and esomeprazole combination dispensing was 9.4% (99%CI 1.0% to 16.5%) lower than forecasted.

#### Nervous system

Among analgesics, paracetamol increased by 18.6% (99%CI 11.7% to 26.3%) and pregabalin by 6.3% (99%CI 1.1% to 12.1%), while tramadol and codeine combinations did not differ significantly. Psycholeptics olanzapine (8.3 %, 99%CI 2.9% to 14.4%) and zolpidem (6.0%, 99%CI 0.6% to 11.9%) increased, while zopiclone, alprazolam, diazepam, and quetiapine did not differ significantly. All common antidepressants included (amitriptyline, citalopram, duloxetine, escitalopram, fluoxetine, mirtazapine, sertraline, and venlafaxine) had significant increases between 6 and 10%. Vertigo medication betahistine was unchanged.

#### Respiratory system

All common individual medications from the drugs for obstructive airway diseases subgroup (R03) had significantly increased dispensing in March 2020 compared to forecast data, with the largest absolute and relative differences. This includes the inhaled bronchodilators salbutamol (33.9%, 99%CI 25.5% to 43.4%), and tiotropium bromide (14.3%, 99%CI 3.9% to 27.1%), inhaled corticosteroid beclomethasone (52.6%, 99%CI 40.4% to 67.1%), and combination inhalers containing formoterol with budesonide (27.2%, 99%CI 18.8% to 36.8%) and salmeterol with fluticasone (21.8%, 99%CI 11.6% to 34.0%). Prescribing of montelukast (16.6%, 99%CI 10.5% to 23.3%) also increased. Dispensing of other respiratory drugs, including carbocisteine, cetirizine, and nasal fluticasone furoate was unchanged.

#### Others

Of the remaining common individual medications, dispensing of ophthalmological preparations artificial tears and latanoprost increased significantly between 7.5 and 8.5%, as did ostomy requisites (11.9%, 99%CI 4.2% to 20.8%). All dermatological preparations, antibacterials for systemic use, and clinical nutritional products did not differ significantly from forecasted in March 2020. The “other” drugs included in the list of most commonly prescribed drugs and grouped into subgroups had similar patterns to the individual drugs. See Supplementary table 2 and Supplementary figure 2 for all included drugs.

### Changes after March 2020

While most medication dispensing returned to forecasted levels after March 2020, certain common individual medications saw persistent differences compared to forecasted dispensing, including antibacterials, systemic steroids, and respiratory medications. Prescribing of amoxicillin was significantly lower than forecasted most months between April 2020 and May 2021, with an average decrease of approximately 50% (Figure 3). Amoxicillin and beta-lactamase inhibitor in combination (i.e. co-amoxiclav) had significantly lower prescribing in December 2020 and January 2021 only. Other antibacterials had a similar pattern to amoxicillin and beta-lactamase inhibitor (see Supplementary figure 2). Systemic prednisolone saw consistently significantly lower prescribing between April 2020 and May 2021, with an average decrease of approximately 29%. There was also an attenuation of the seasonal peak in winter for prednisolone, and for the antibacterials. Conversely, dispensing of the combination inhaler containing formoterol and budesonide was significantly higher than forecast in March 2020, and dispensing remained significantly higher than forecast for much of the time between April 2020 and December 2021, on average 16% higher. For “Other drugs for obstructive airway disease” there was a March 2020 peak, however dispensing was at the lower end, or outside of the forecasted interval, from 2021 onwards (Supplementary figure 2).

**Figure 3.**
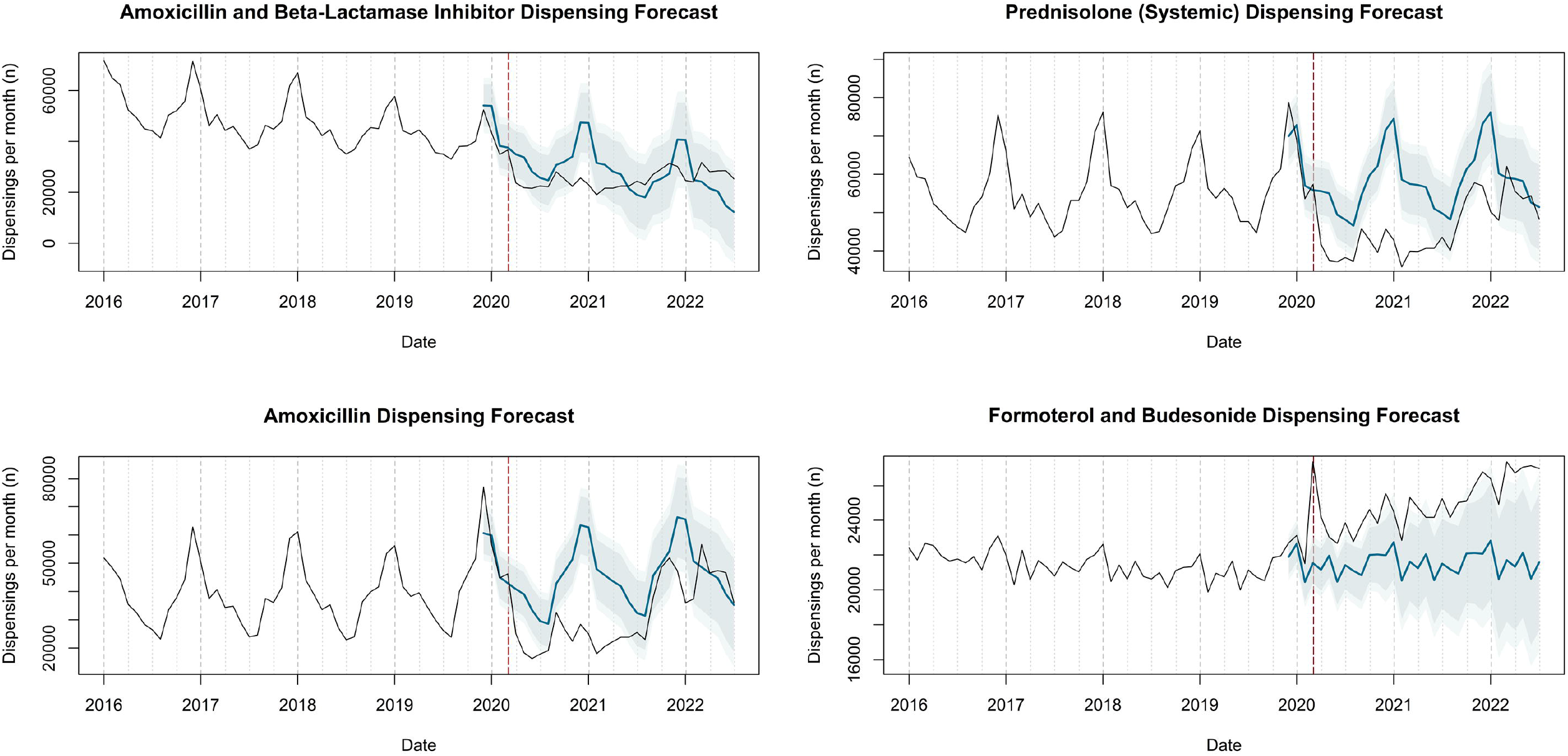
Actual dispensing (black) and forecasted dispensing (blue, with 95% and 99% prediction intervals) for individual medications with dispensing that diverged from forecast after March 2020

## Discussion

### Principal findings

Over the study period, there was an increase in annual number of dispensings on the GMS scheme in Ireland, in spite of a reduction in the eligible population numbers. Total dispensings in March 2020 were 7.6% higher than forecasted, and over one third of therapeutic subgroups had dispensings significantly above what was expected; drugs for obstructive airway disease had the greatest divergence (26.2%) from the forecast. Only two therapeutic subgroups had dispensings lower than forecasted for March 2020. Many individual medications also had higher dispensings in March 2020 than forecasted, inhaled beclomethasone having the greatest difference of 52.6%. Differences from the forecast after the onset of the pandemic were observed, with lower than expected dispensings of amoxicillin (alone and as co-amoxiclav) and oral prednisolone.

### Interpretation of findings

The forecast generally performed well, the exception being anti-anaemic preparations and ferrous fumarate specifically. This may be attributable to intermittent supply shortages affecting ferrous fumarate during 2019 (noted as resolved in December 2019) and 2020 (resolved in August 2020).[12, 13] Dispensing of medications for most common chronic conditions increased significantly, with generally similar increases across medication in most classes, and this was true of physical and mental health conditions, and applied across oral medications, topical products, and inhalers.[14-16] This likely reflected individuals getting additional supplies of their regular medication due to concerns with regards to supply and fear of impending restrictions. Potential evidence of stockpiling immediately before pandemic restrictions were implemented has been identified in studies across multiple countries.[15]

Respiratory medication for obstructive pulmonary disease as a subgroup, and individual medications, saw the largest peak in March 2020. The higher magnitude of peak for these medications may reflect greater precautions among people with existing respiratory conditions, given that COVID-19 is primarily a respiratory illness. Although formoterol/budesonide appeared to be increasing beyond forecasted levels from 2021 onwards, which could potentially be attributed to an increase in diagnosis and treatment of respiratory conditions, this was concurrent with lower than expected levels of other obstructive pulmonary disease drugs. The overall dispensing within this subgroup closely mirrored the forecast after March 2020, suggesting no detectable excess of new respiratory diagnoses after the onset of the pandemic in this study. A multi-country European study found a mixed pattern with some regions and countries with reduced volumes of obstructive pulmonary disease prescribing in the 12 months from March 2020 onward relative to the previous 12 months, and increased volume in others.[15] The authors of this study postulate that these reduced volumes could be due to lower exposure to factors likely to exacerbate respiratory conditions due to lockdowns.

The lower than expected dispensings of amoxicillin and co-amoxiclav may reflect lower rates of transmission of respiratory infections in the months after the onset of the pandemic, particularly during winter 2020/2021 with a loss of the seasonal peak in medication use, likely due to social distancing and increased mask-wearing.[17] International infection surveillance corroborates this, with significant reductions in respiratory infections 4-8 weeks after the introduction of COVID restrictions, and record lows in influenza cases.[18, 19] Another factor which may have contributed to reduced antimicrobial prescribing and enhanced antimicrobial choice, independent of the COVID-19 pandemic, was the introduction of the “Greed Red” antibiotic quality improvement initiative for community prescribers by the Health Services Executive in the Republic of Ireland in 2019.[20] Studies in the United States. Belgium and Scotland found similar reductions in antibiotic prescribing in the months following March 2020,[21-23] and a greatly diminished seasonal peak,[24] with reductions across all age groups, and suggestions that reductions were greatest for agents to treat respiratory infections. A study in the Netherlands also identified large relative reduction in prescribing for other types of infections, including gastrointestinal and skin infections.[25]

With regard to analgesia, we identified higher than expected paracetamol dispensing but not for codeine/paracetamol combinations or tramadol. Although a peak in paracetamol prescribing in England was potentially linked to shortages in non-prescription paracetamol due to stockpiling, it is unclear if similar shortages occurred in the Republic of Ireland.[4] This may also reflect paracetamol use as first-line regular treatment for arthritis, and so may largely be used as a regular medication for a chronic condition (and thus an increase in dispensing in line with other such medications), whereas opioids may be prescribed more often as episodic or as needed treatment, including as post-operative analgesia. Naproxen/esomeprazole was the only individual medication with dispensing significantly lower than forecast, which again may be due reduced need as a treatment post-operatively or due to injuries. International evidence suggests a non-operation rate of 15% in cancer surgery associated with periods of full lockdown restrictions,[26] while in Ireland, the pandemic was associated with reductions in emergency and elective surgeries of 17% and 30% respectively.[27]

We identified lower than expected dispensing of dressings. These are primarily supplied as stock orders for GP practices, and thus may be linked to reduced availability of in-person appointments with healthcare professionals immediately after the onset of the pandemic. The other subgroup with levels lower than forecasted in March 2020 was ‘other gynaecologicals’. Although this includes intravaginal contraceptives and uterotonics, the majority of dispensing in this group is accounted for by intrauterine contraceptive devices (1,386 of 2,355 items in February 2020, or 59%), which require a visit to a healthcare professional for insertion. This may suggest individuals deferring intrauterine device insertion due to reduced access to healthcare providers, and supported by an apparent increase in dispensing of the combined oral contraceptive levonorgestrel and ethinylestradiol. This mirrored patterns identified in England, where the most significant reductions in contraceptive prescribing from April to June 2020 were for those requiring a healthcare consultation for implantation or insertion.[4, 28]

### Strengths and limitations

This is the first study to examine changes in medication use following the onset of the COVID-19 pandemic in the Republic of Ireland, and one of few studies internationally to examine changes in prescribing across the full range of therapeutic areas. A limitation is that the data relates to the GMS scheme, which covers only one third of the population, over-representing older adults and those of lower socioeconomic status. It is possible that the pandemic may have had a differential impact on prescribing in this group, for example widening or narrowing existing differences in prescribing associated with deprivation.[29] Although model forecasts generally performed well against several months of pre-pandemic data, it did not match observed data in one case where a supply shortage occurred towards the end of the “forecast” data time period. There may have been other concurrent events or factor that could have affected dispensing of some classes, however the broad consistency in observed pattern across a large proportion of therapeutic subgroups and medications is supportive of a pandemic-related impact.

## Conclusions

This study found widespread peaks in medication dispensing during March 2020 on the eve of the COVID-19 pandemic in Ireland. It also identified lower than expected dispensing for items related to in-person consultations, as well as for some treatments for respiratory infections which persisted through to 2021. Although the acute phase of the pandemic has passed, this study enhances the evidence of how health system delivery continued in Ireland during 2020 and beyond. This also provides evidence for planning responses to future health system threats to medication supply chain and averting shortages, an increasingly common challenge for health systems outside of the pandemic setting.

## Data Availability

The data that support the findings of this study are available in Zenodo at https://doi.org/10.5281/zenodo.7999791. These data were derived from the Health Service Executive Primary Care Reimbursement Service's Reporting and Open Data platform available in the public domain: https://www.sspcrs.ie/portal/annual-reporting/

## Acknowledgments

MM is supported by a grant from the Health Research Board under their Secondary Data Analysis Projects scheme (CDRx project, SDAP-2019-023, PI FM); JAH contributed to this research while on placement and was supported by funds from the School of Pharmacy and Biomolecular Sciences, RCSI University of Medicine and Health Sciences.

## Supplementary files

**Supplementary table 1**. Actual and forecasted dispensing in March 2020 and differences with 99% prediction intervals for therapeutic subgroups

**Supplementary table 2**. Actual and forecasted dispensing in March 2020 and differences with 99% prediction intervals for individual medications

**Supplementary figure 1**. Actual dispensing (black) and forecasted dispensing (blue, with 95% and 99% prediction intervals) for all therapeutic subgroups

**Supplementary table 2**. Actual dispensing (black) and forecasted dispensing (blue, with 95% and 99% prediction intervals) for all individual medications

## References

1. Wabe N, Thomas J, Sezgin G, Sheikh MK, Gault E, Georgiou A. Medication prescribing in face-to-face versus telehealth consultations during the COVID-19 pandemic in Australian general practice: a retrospective observational study. BJGP Open. 2022;6:pBJGPO.2021.0132.

2. Gleeson LL, Ludlow A, Wallace E, Argent R, Collins C, Clyne B, et al. Changes to primary care delivery during the COVID-19 pandemic and perceived impact on medication safety: A survey study. Explor Res Clin Soc Pharm. 2022;6:100143.

3. Gleeson LL, Ludlow A, Clyne B, Ryan B, Argent R, Barlow J, et al. Pharmacist and patient experiences of primary care during the COVID-19 pandemic: An interview study. Explor Res Clin Soc Pharm. 2022;8:100193.

4. Frazer JS, Frazer GR. Analysis of primary care prescription trends in England during the COVID-19 pandemic compared against a predictive model. Fam Med Community Health. 2021;9.

5. Clement J, Jacobi M, Greenwood BN. Patient access to chronic medications during the Covid-19 pandemic: Evidence from a comprehensive dataset of US insurance claims. PLoS One. 2021;16:e0249453.

6. Campitelli MA, Bronskill SE, Maclagan LC, Harris DA, Cotton CA, Tadrous M, et al. Comparison of Medication Prescribing Before and After the COVID-19 Pandemic Among Nursing Home Residents in Ontario, Canada. JAMA Network Open. 2021;4:e2118441–e.

7. Selke Krulichová I, Selke GW, Bennie M, Hajiebrahimi M, Nyberg F, Fürst J, et al. Comparison of drug prescribing before and during the COVID-19 pandemic: A cross-national European study. Pharmacoepidemiol Drug Saf. 2022;31:1046–55.

8. Mattsson M, Hong JA, Frazer JS, Frazer GR, Moriarty F. Trends in medication use after the onset of the COVID-19 pandemic in the Republic of Ireland. Open Science Framework.

9. Mattsson M, Flood M, Wallace E, Boland F, Moriarty F. Eligibility rates and representativeness of the General Medical Services scheme population in Ireland 2017-2021: A methodological report [version 1; peer review: awaiting peer review]. HRB Open Research. 2022;5.

10. Sinnott SJ, Bennett K, Cahir C. Pharmacoepidemiology resources in Ireland-an introduction to pharmacy claims data. Eur J Clin Pharmacol. 2017;73:1449–55.

11. Hyndman R. forecast: Forecasting functions for time series. 2008.

12. HPRA. Resolved shortages 6/4/2020. Health Products Regulatory Alliance; 2020.

13. HPRA. Resolved shortages 11/10/2020. Health Products Regulatory Authority 2020.

14. Reppas-Rindlisbacher C, Rochon PA, Stall NM. The COVID-19 Pandemic and Drug Prescribing in Ontario Nursing Homes-From Confinement Syndrome to Unconfined Prescribing. JAMA Netw Open. 2021;4:e2119028.

15. Selke Krulichová I, Selke GW, Bennie M, Hajiebrahimi M, Nyberg F, Fürst J, et al. Comparison of drug prescribing before and during the COVID-19 pandemic: A cross-national European study. Pharmacoepidemiology and Drug Safety. 2022;31:1046–55.

16. Campitelli MA, Bronskill SE, Maclagan LC, Harris DA, Cotton CA, Tadrous M, et al. Comparison of Medication Prescribing Before and After the COVID-19 Pandemic Among Nursing Home Residents in Ontario, Canada. JAMA Netw Open. 2021;4:e2118441.

17. Zhang T, Shen X, Liu R, Zhao L, Wang D, Lambert H, et al. The impact of COVID-19 on primary health care and antibiotic prescribing in rural China: qualitative study. BMC Health Services Research. 2021;21:1048.

18. Brueggemann AB, Jansen van Rensburg MJ, Shaw D, McCarthy ND, Jolley KA, Maiden MCJ, et al. Changes in the incidence of invasive disease due to Streptococcus pneumoniae, Haemophilus influenzae, and Neisseria meningitidis during the COVID-19 pandemic in 26 countries and territories in the Invasive Respiratory Infection Surveillance Initiative: a prospective analysis of surveillance data. Lancet Digit Health. 2021;3:e360–e70.

19. Adlhoch C, Mook P, Lamb F, Ferland L, Melidou A, Amato-Gauci AJ, et al. Very little influenza in the WHO European Region during the 2020/21 season, weeks140 2020 to 8 2021. Eurosurveillance. 2021;26:2100221.

20. HSE. Green Red antibiotic QI initiative for community prescribers. Health Service Executive; 2022 [updated 2022; cited]; Available from: https://www.hse.ie/eng/services/list/2/gp/antibiotic-prescribing/antibicrobial-stewardship-audit-tools/campaign-materials/.

21. Malcolm W, Seaton RA, Haddock G, Baxter L, Thirlwell S, Russell P, et al. Impact of the COVID-19 pandemic on community antibiotic prescribing in Scotland. JAC-Antimicrobial Resistance. 2020;2:dlaa105.

22. Colliers A, De Man J, Adriaenssens N, Verhoeven V, Anthierens S, De Loof H, et al. Antibiotic Prescribing Trends in Belgian Out-of-Hours Primary Care during the COVID-19 Pandemic: Observational Study Using Routinely Collected Health Data. Antibiotics (Basel). 2021;10.

23. King LM, Lovegrove MC, Shehab N, Tsay S, Budnitz DS, Geller AI, et al. Trends in US Outpatient Antibiotic Prescriptions During the Coronavirus Disease 2019 Pandemic. Clinical Infectious Diseases. 2021;73:e652–e60.

24. Buehrle DJ, Wagener MM, Nguyen MH, Clancy CJ. Trends in Outpatient Antibiotic Prescriptions in the United States During the COVID-19 Pandemic in 2020. JAMA Network Open. 2021;4:e2126114–e.

25. van de Pol AC, Boeijen JA, Venekamp RP, Platteel T, Damoiseaux R, Kortekaas MF, et al. Impact of the COVID-19 Pandemic on Antibiotic Prescribing for Common Infections in The Netherlands: A Primary Care-Based Observational Cohort Study. Antibiotics (Basel). 2021;10.

26. COVIDSurg Collaborative. Effect of COVID-19 pandemic lockdowns on planned cancer surgery for 15 tumour types in 61 countries: an international, prospective, cohort study. The Lancet Oncology. 2021;22:1507–17.

27. NCPS. The impact of the Coronavirus Pandemic on Surgery in Ireland. National Clinical Programme in Surgery; 2021.

28. Walker SH. Effect of the COVID-19 pandemic on contraceptive prescribing in general practice: a retrospective analysis of English prescribing data between 2019 and 2020. Contraception and Reproductive Medicine. 2022;7:3.

29. Prendergast C, Flood M, Murry L, Clyne B, Fahey T, Moriarty F. Prescribing differences among older adults with differing health cover and socioeconomic status: a cohort study. medRxiv. 2023:2023.03.30.23287967.

