## Supplementary figure 1 for "Trends in medication use after the onset of the COVID-19 pandemic in the Republic of Ireland: an interrupted time series study"

**Supplementary figure 1.** Actual dispensing (black) and forecasted dispensing (blue, with 95% and 99% prediction intervals) for all therapeutic subgroups

*Therapeutic subgroups appear in alphabetical order by subgroup name*

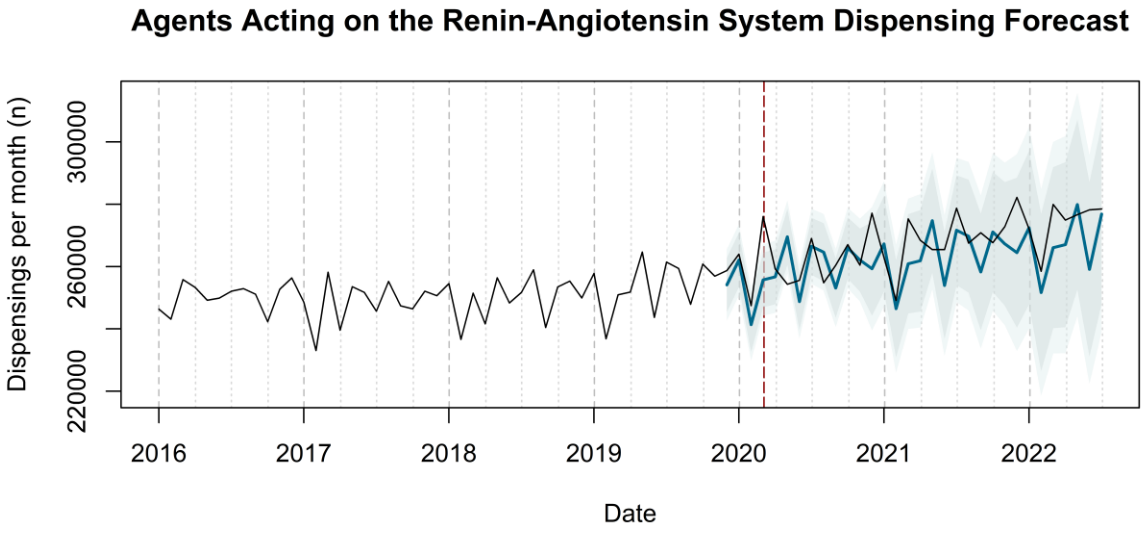


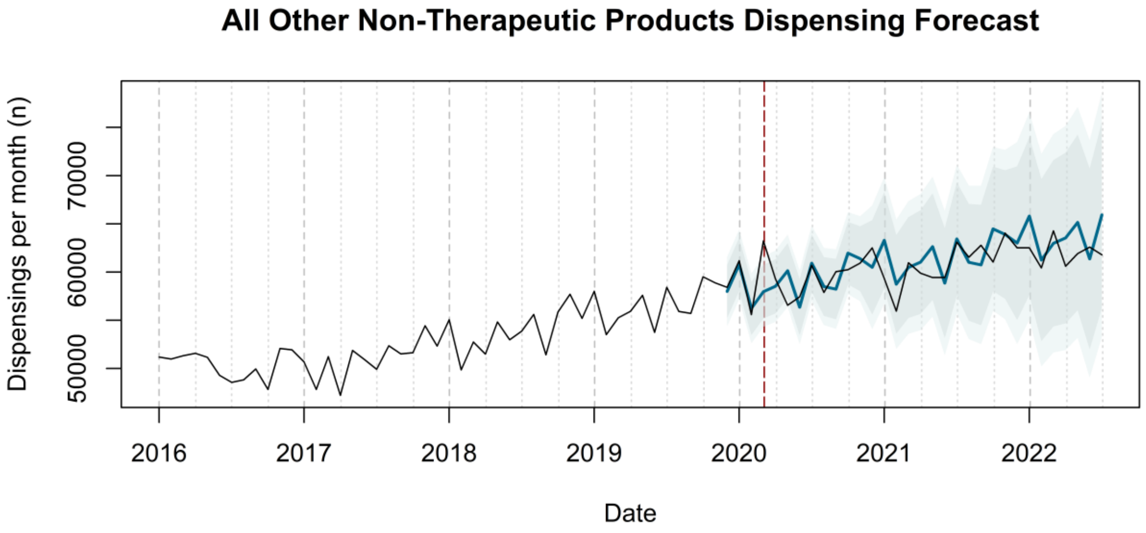

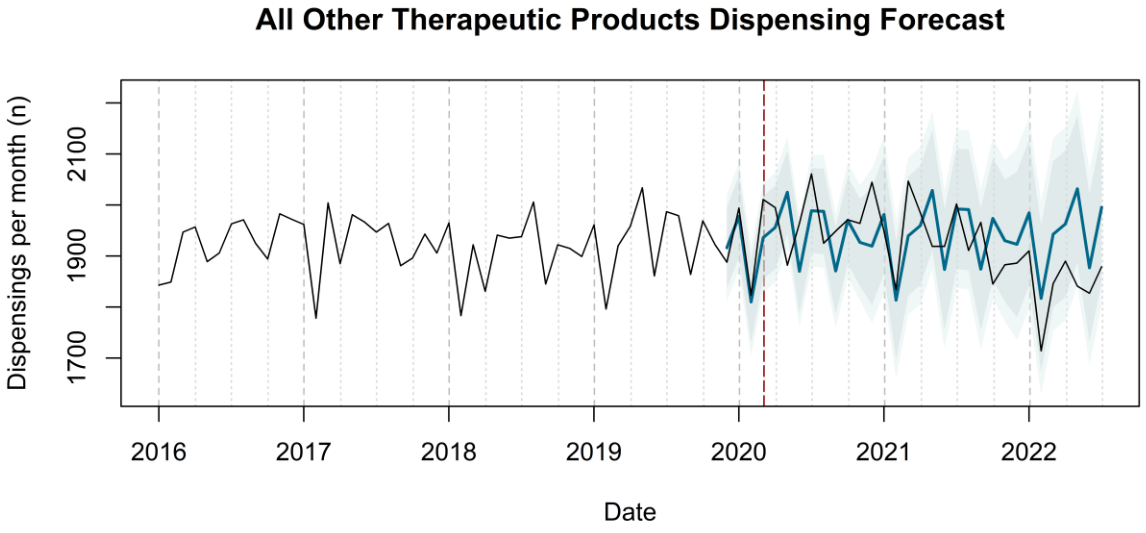

All Other Therapeutic Products Dispensing Forecast
Dispensings per month (n)
1700
1900
2100
2016
2017
2018
2019
2020
2021
2022
Date


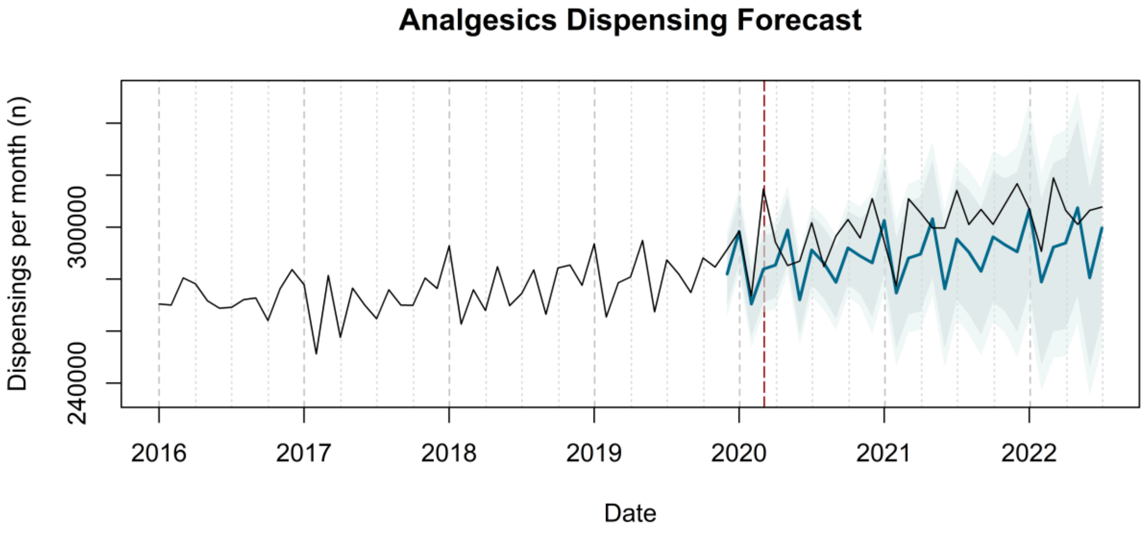

Analgesics Dispensing Forecast
Dispensings per month (n)
240000
300000
2016
2017
2018
2019
2020
2021
2022
Date


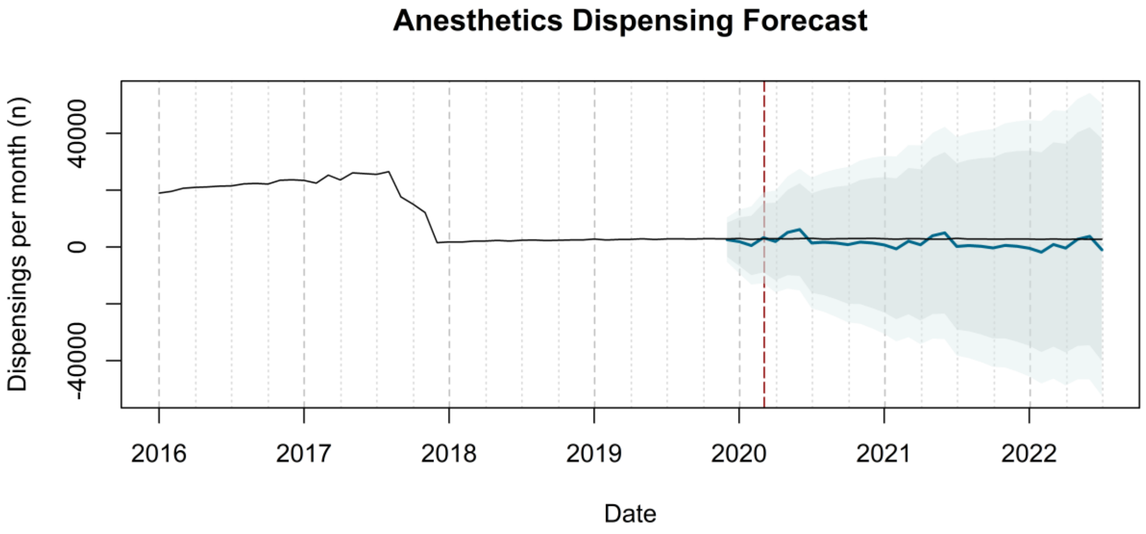

Anesthetics Dispensing Forecast
Dispensings per month (n)
-40000
0
40000
2016
2017
2018
2019
2020
2021
2022
Date

### Anthelmintics Dispensing Forecast

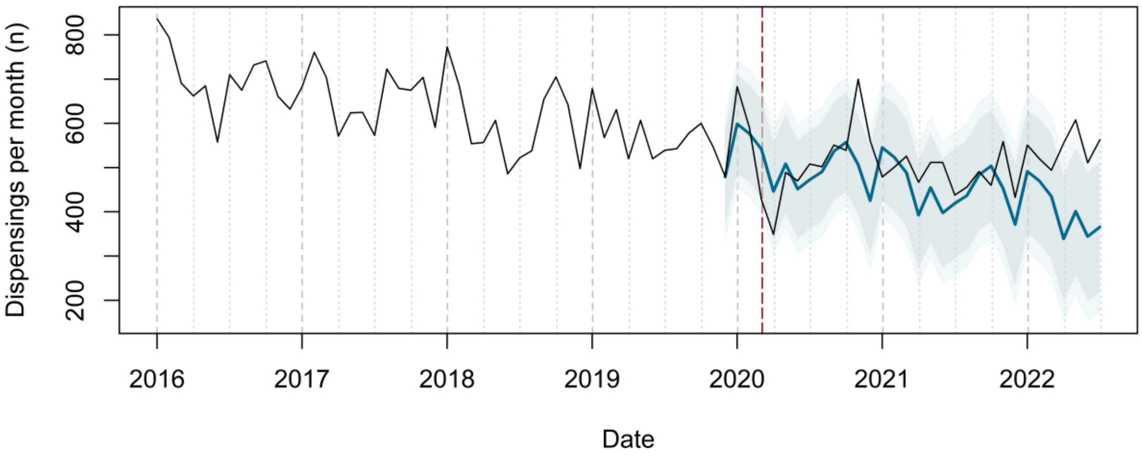


### Anti-Acne Preparations Dispensing Forecast

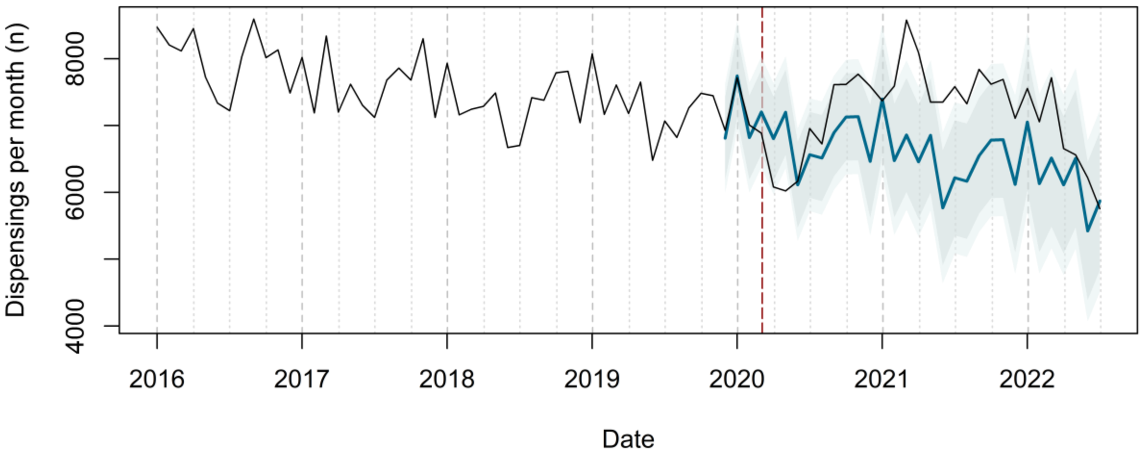


### Antianemic Preparations Dispensing Forecast

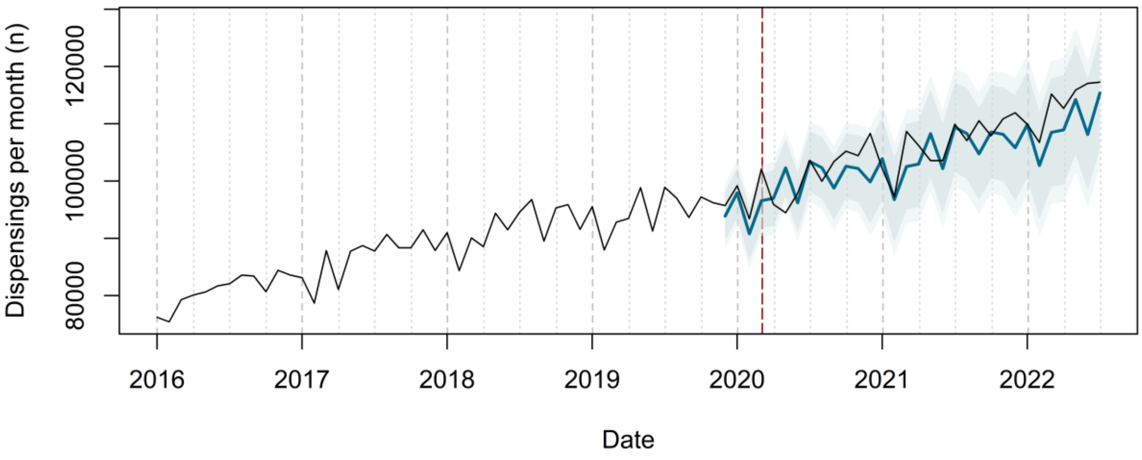

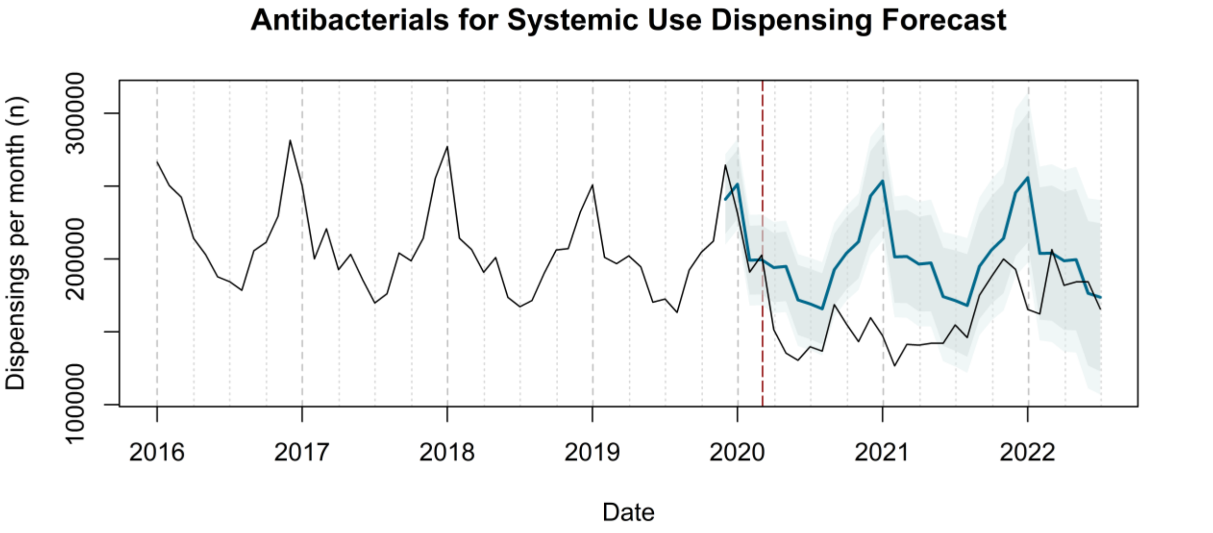

Antibacterials for Systemic Use Dispensing Forecast
Dispensings per month (n)
100000
200000
300000
2016
2017
2018
2019
2020
2021
2022
Date


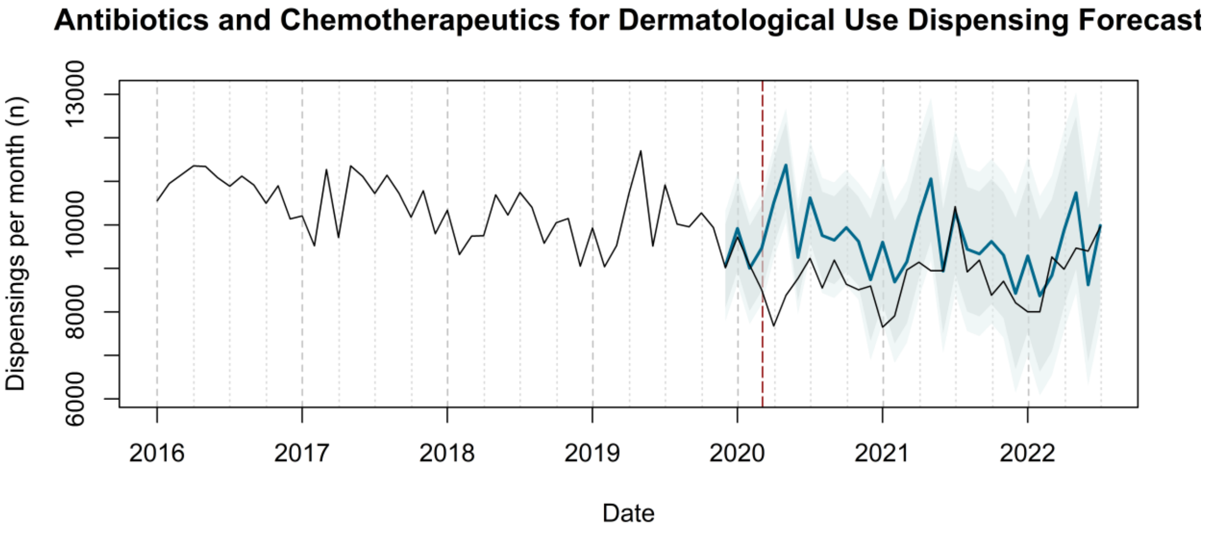

Antibiotics and Chemotherapeutics for Dermatological Use Dispensing Forecast
Dispensings per month (n)
6000
8000
10000
13000
2016
2017
2018
2019
2020
2021
2022
Date


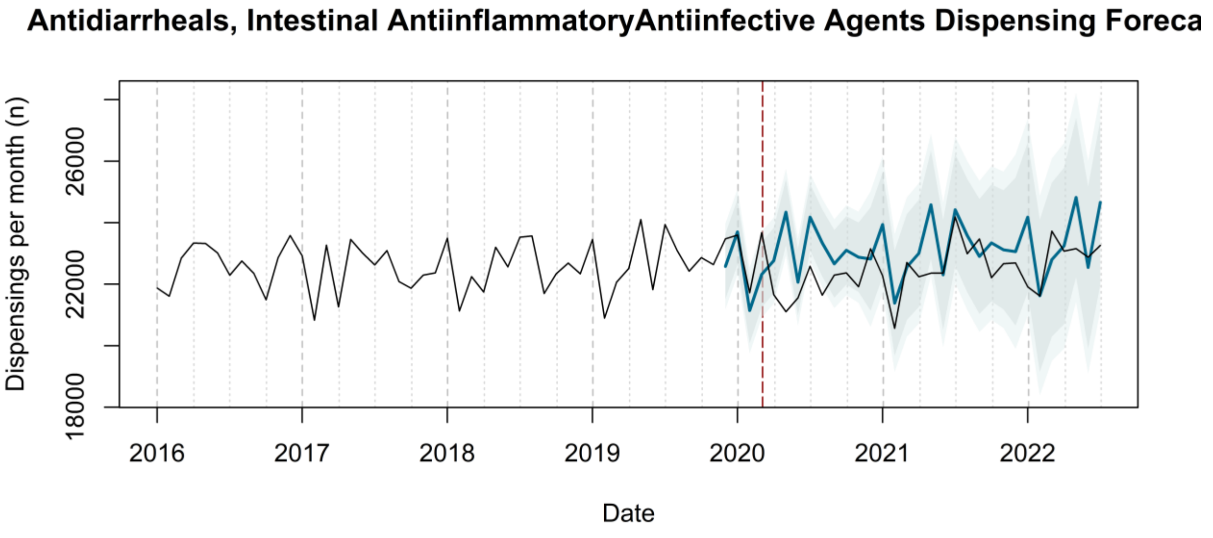

Antidiarrheals, Intestinal AntiinflammatoryAntiinfective Agents Dispensing Foreca
Dispensings per month (n)
18000
22000
26000
2016
2017
2018
2019
2020
2021
2022
Date

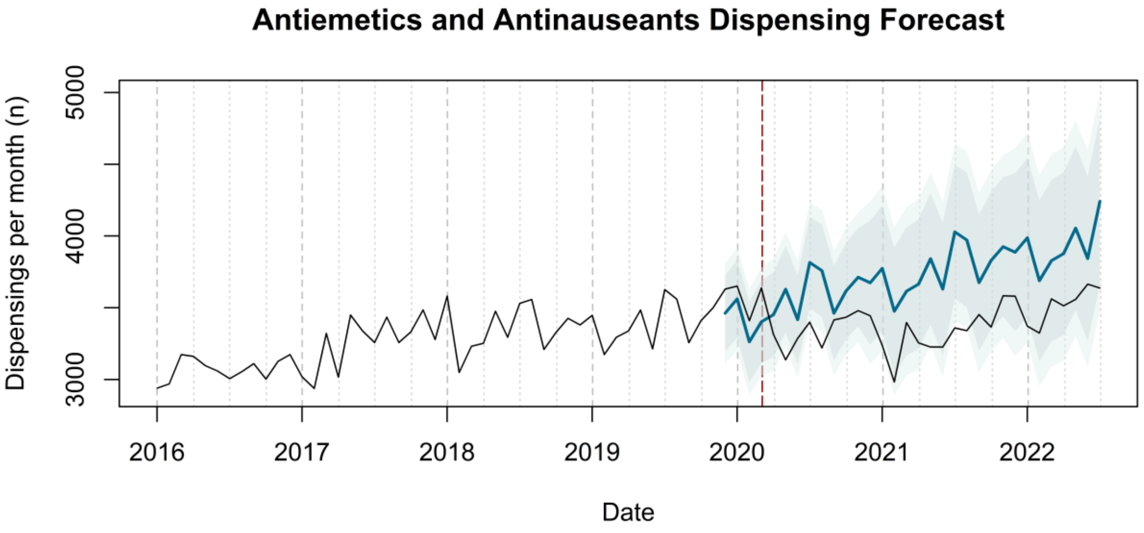

Antiemetics and Antinauseants Dispensing Forecast
Dispensings per month (n)
3000
4000
5000
2016
2017
2018
2019
2020
2021
2022
Date


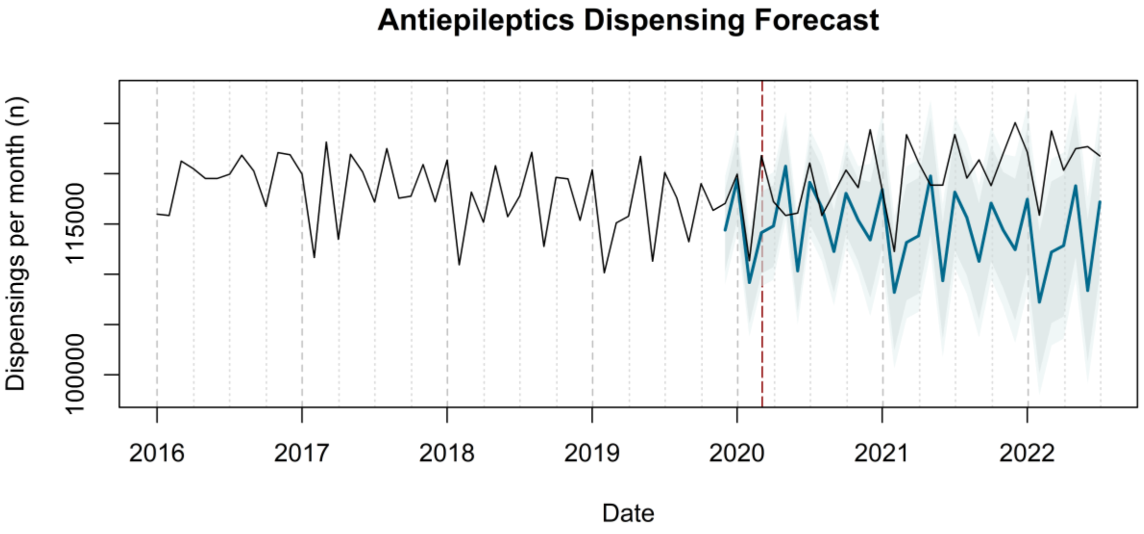

Antiepileptics Dispensing Forecast
Dispensings per month (n)
100000
115000
2016
2017
2018
2019
2020
2021
2022
Date


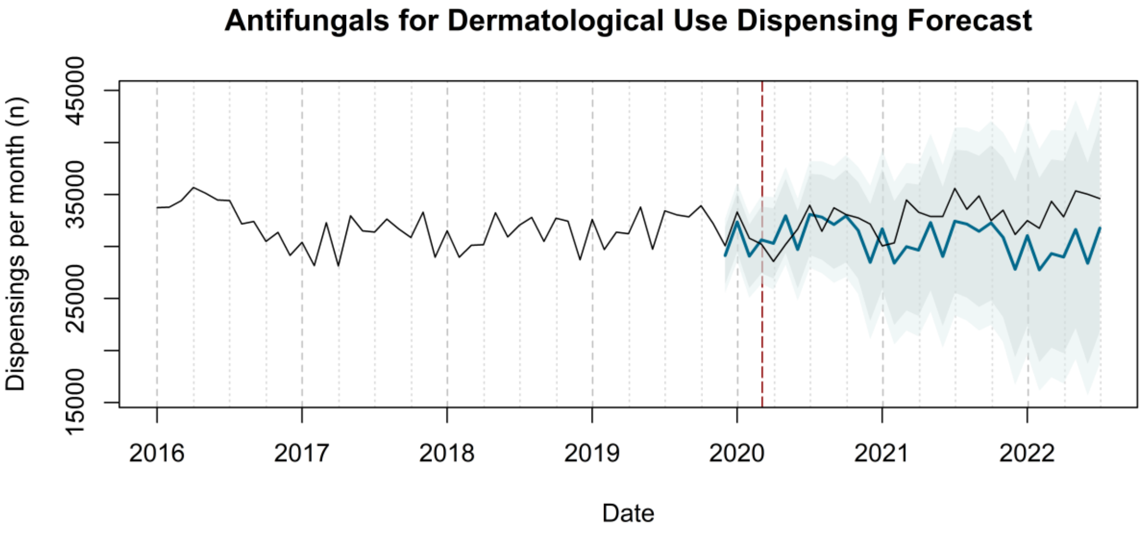

Antifungals for Dermatological Use Dispensing Forecast
Dispensings per month (n)
15000
25000
35000
45000
2016
2017
2018
2019
2020
2021
2022
Date

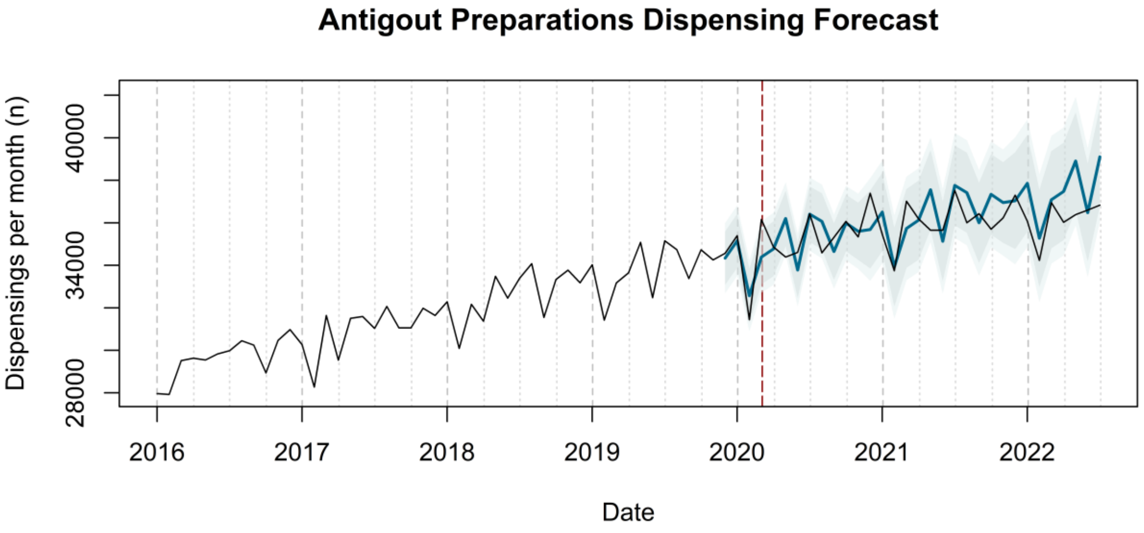
Antigout Preparations Dispensing Forecast
Dispensings per month (n)
28000
34000
40000
2016
2017
2018
2019
2020
2021
2022
Date

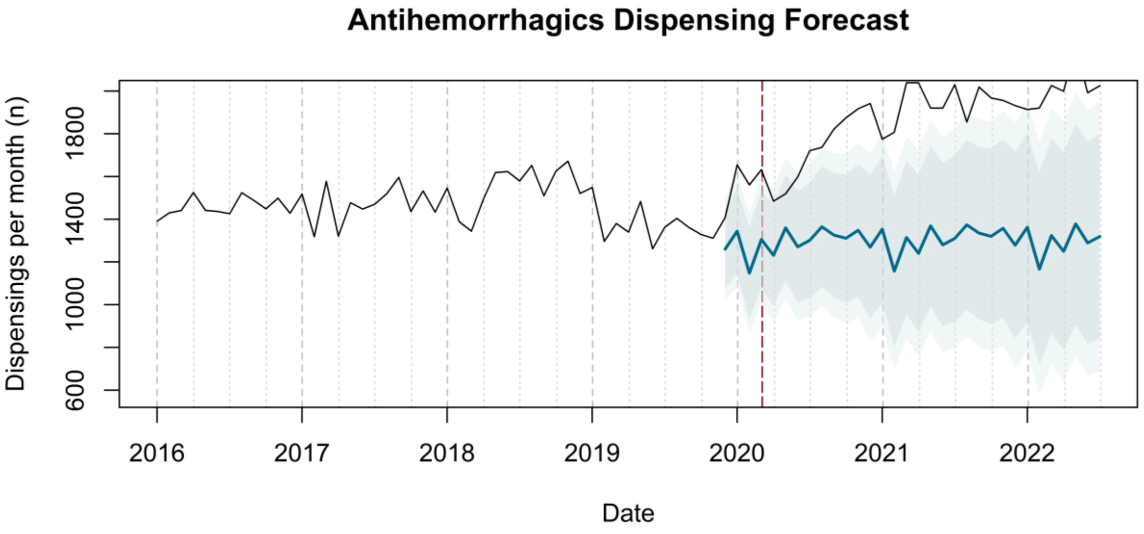
Antihemorrhagics Dispensing Forecast
Dispensings per month (n)
600
1000
1400
1800
2016
2017
2018
2019
2020
2021
2022
Date

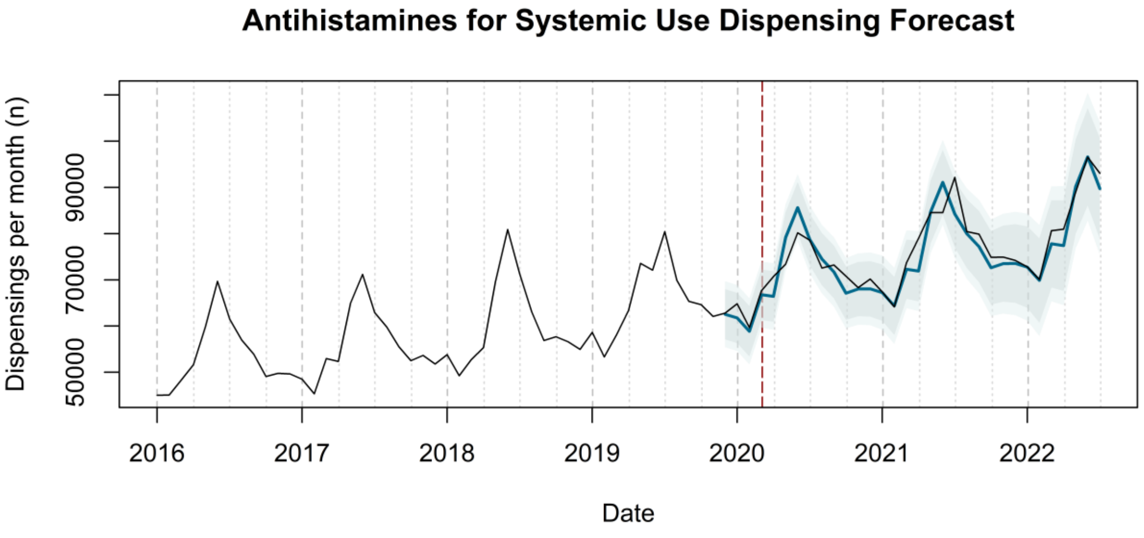
Antihistamines for Systemic Use Dispensing Forecast
Dispensings per month (n)
50000
70000
90000
2016
2017
2018
2019
2020
2021
2022
Date

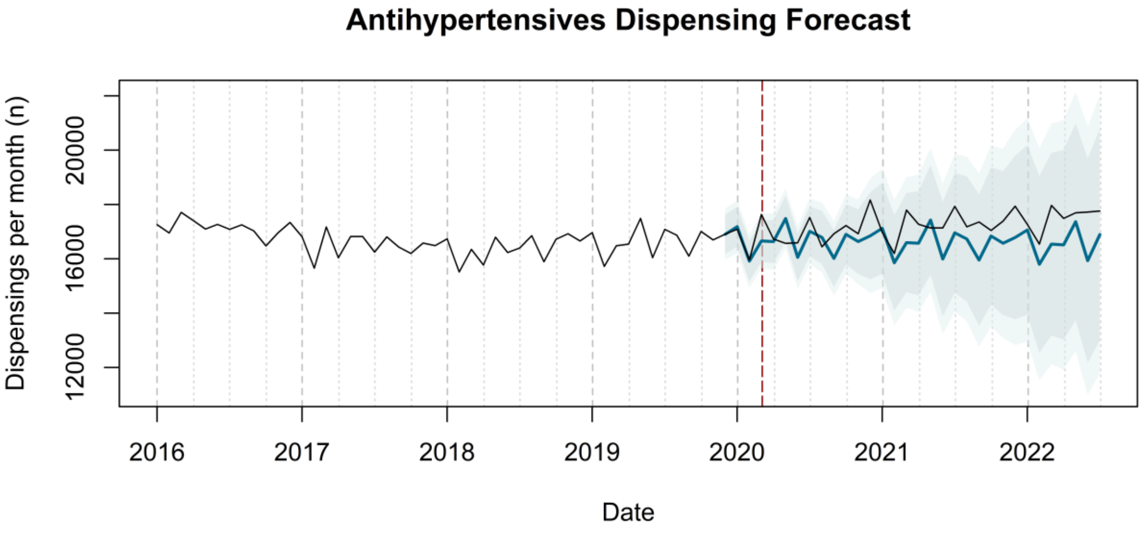
Antihypertensives Dispensing Forecast
Dispensings per month (n)
12000
16000
20000
2016
2017
2018
2019
2020
2021
2022
Date

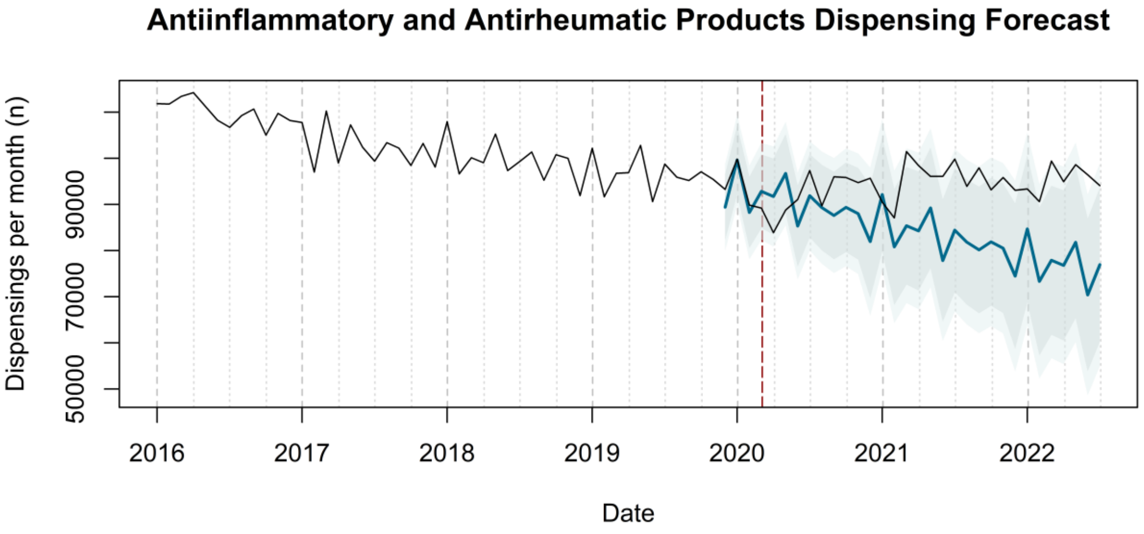
Antiinflammatory and Antirheumatic Products Dispensing Forecast
Dispensings per month (n)
50000
70000
90000
2016
2017
2018
2019
2020
2021
2022
Date

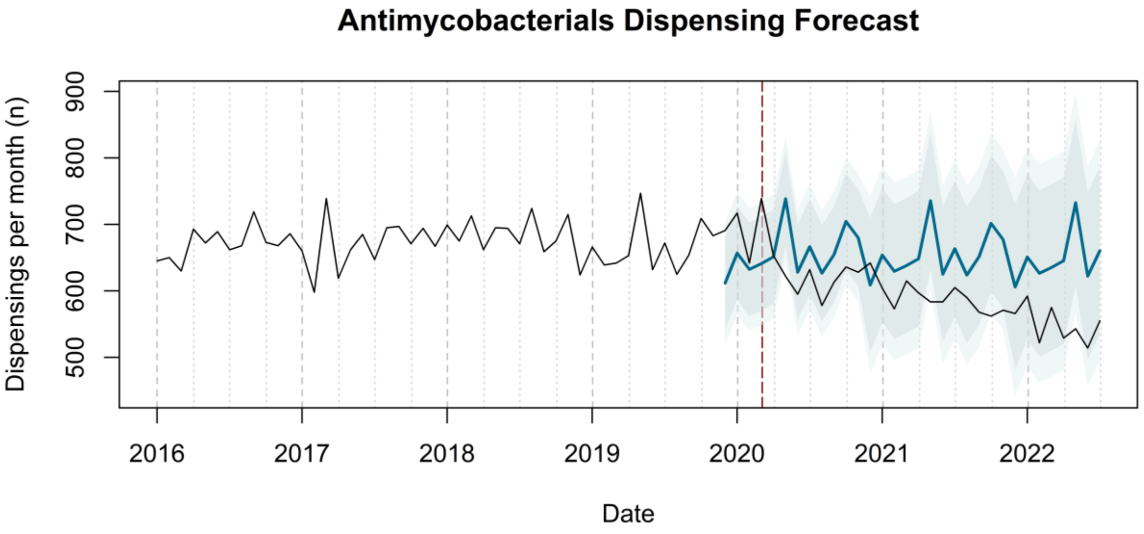
Antimycobacterials Dispensing Forecast
Dispensings per month (n)
500
600
700
800
900
2016
2017
2018
2019
2020
2021
2022
Date

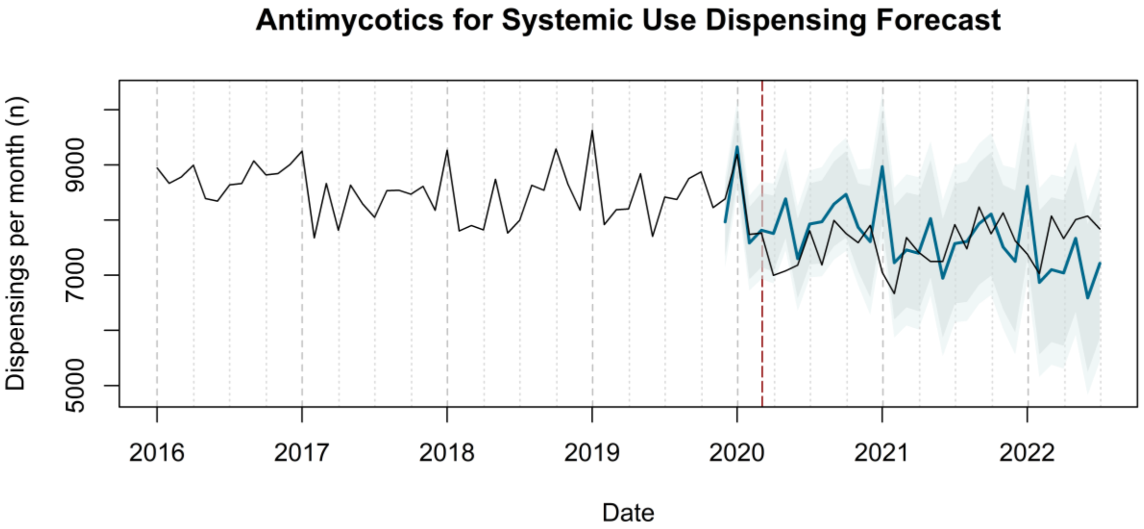

Antimycotics for Systemic Use Dispensing Forecast
Dispensings per month (n)
5000
7000
9000
2016
2017
2018
2019
2020
2021
2022
Date


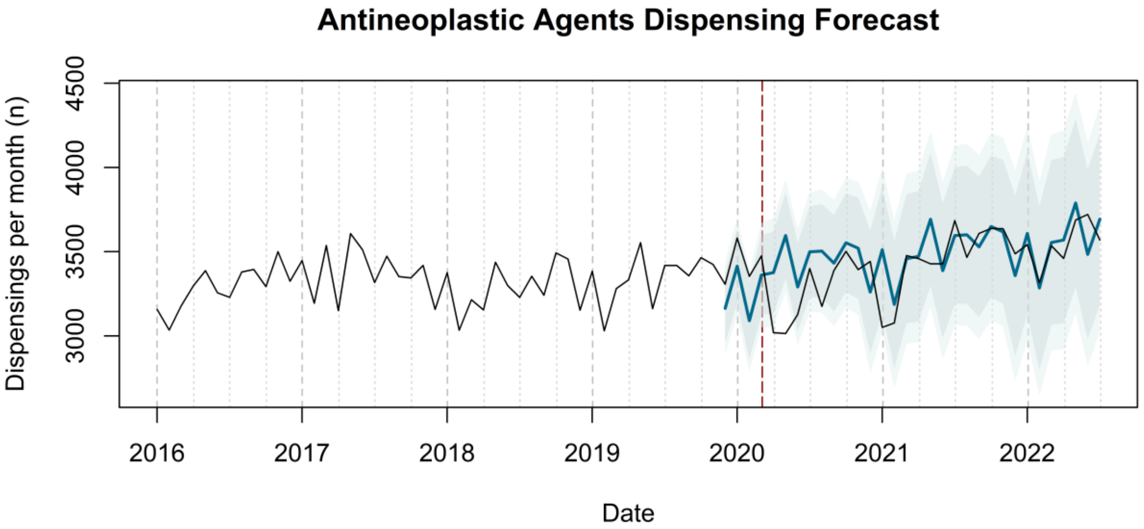

Antineoplastic Agents Dispensing Forecast
Dispensings per month (n)
3000
3500
4000
4500
2016
2017
2018
2019
2020
2021
2022
Date


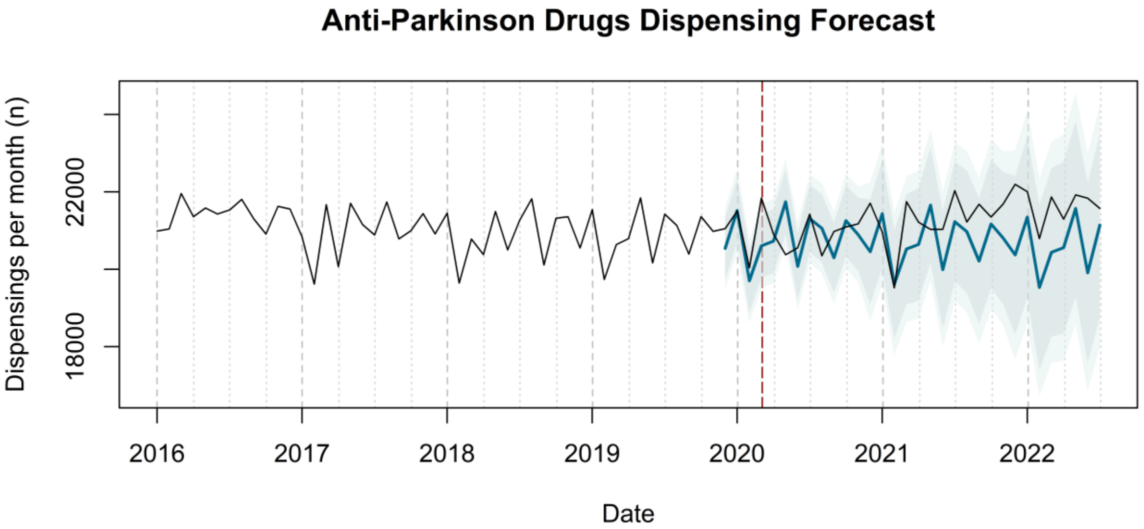

Anti-Parkinson Drugs Dispensing Forecast
Dispensings per month (n)
18000
22000
2016
2017
2018
2019
2020
2021
2022
Date

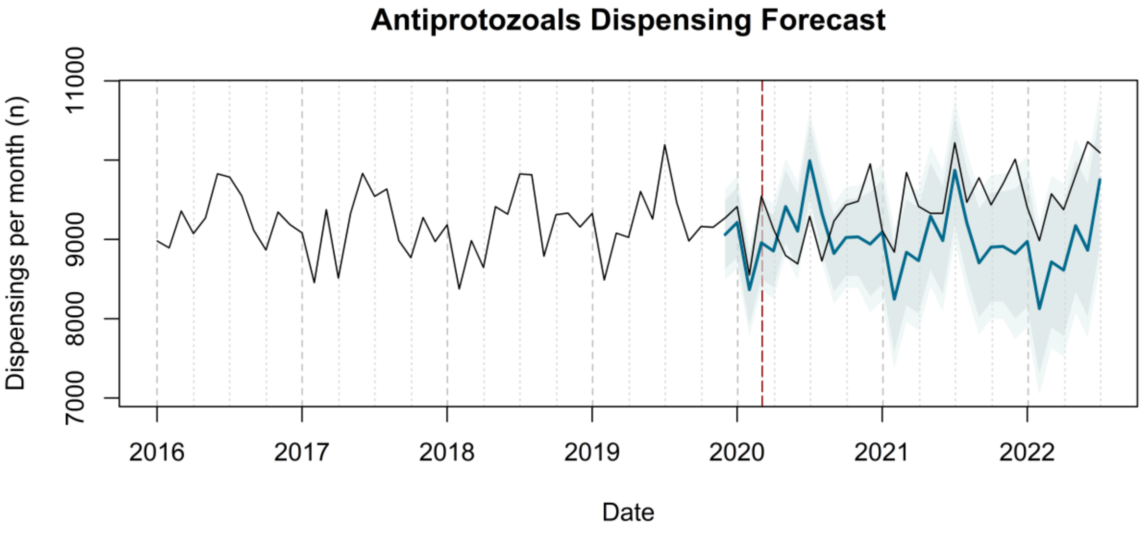
Antiprotozoals Dispensing Forecast
Dispensings per month (n)
7000
8000
9000
11000
2016
2017
2018
2019
2020
2021
2022
Date

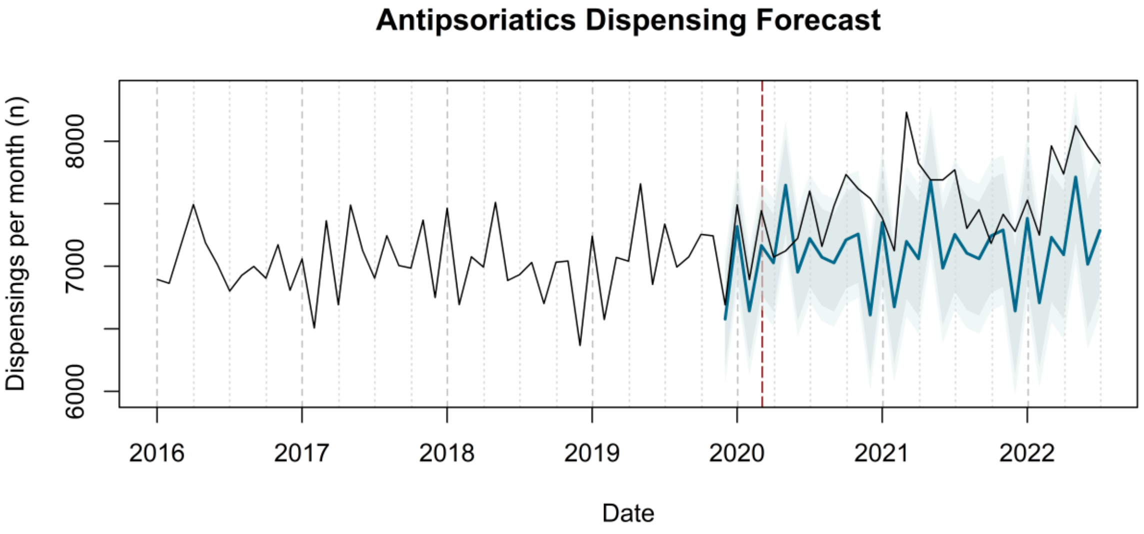
Antipsoriatics Dispensing Forecast
Dispensings per month (n)
6000
7000
8000
2016
2017
2018
2019
2020
2021
2022
Date

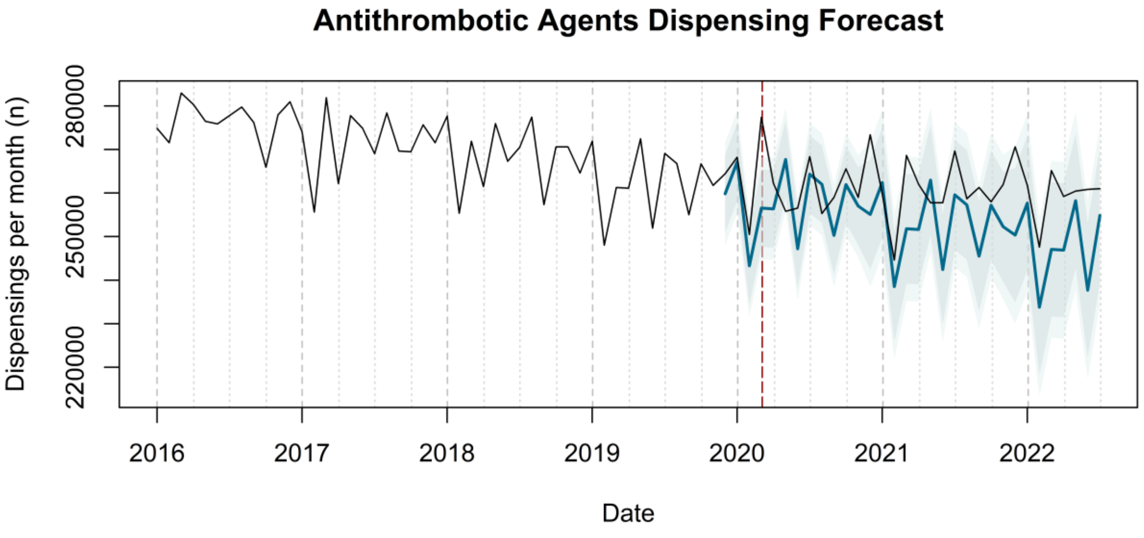
Antithrombotic Agents Dispensing Forecast
Dispensings per month (n)
220000
250000
280000
2016
2017
2018
2019
2020
2021
2022
Date

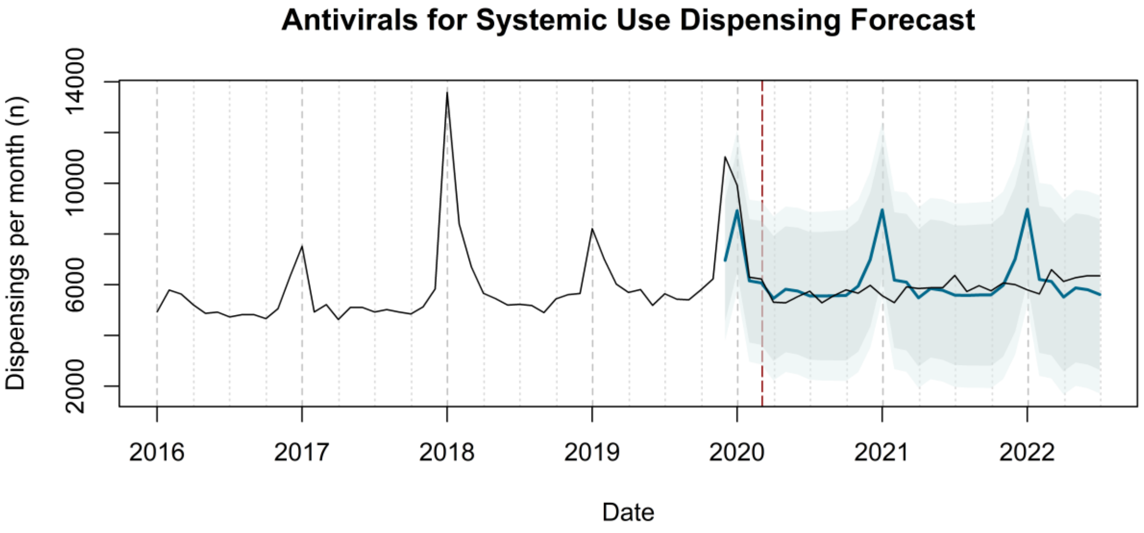

Antivirals for Systemic Use Dispensing Forecast
Dispensings per month (n)
2000
6000
10000
14000
2016
2017
2018
2019
2020
2021
2022
Date


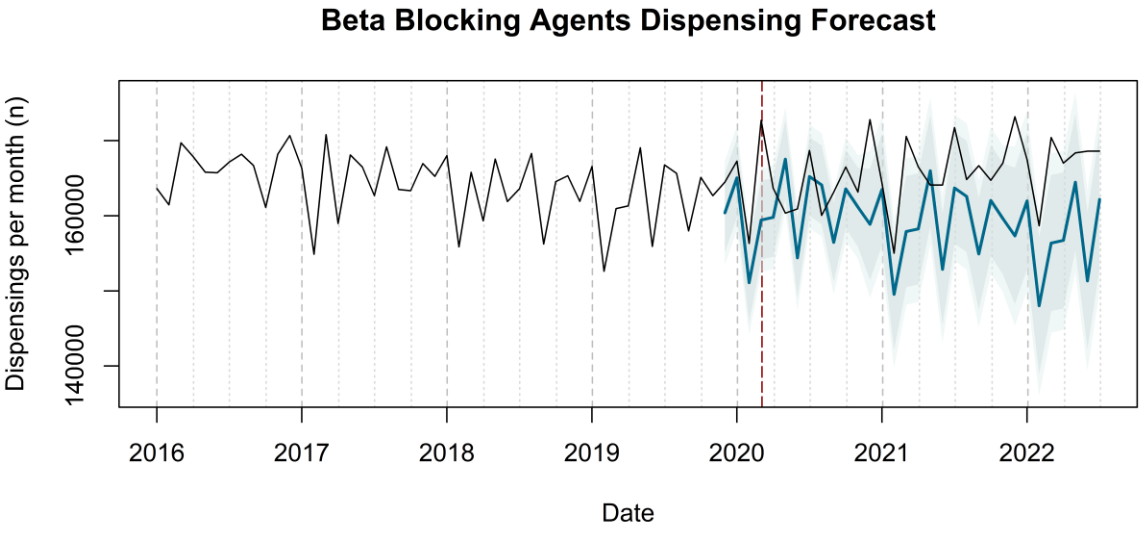

Beta Blocking Agents Dispensing Forecast
Dispensings per month (n)
140000
160000
2016
2017
2018
2019
2020
2021
2022
Date


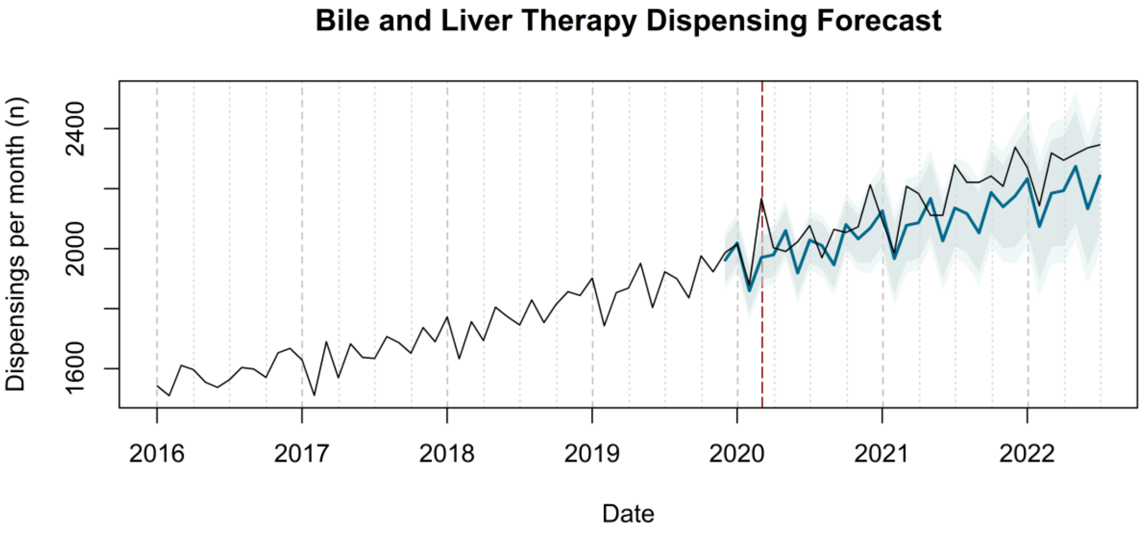

Bile and Liver Therapy Dispensing Forecast
Dispensings per month (n)
1600
2000
2400
2016
2017
2018
2019
2020
2021
2022
Date

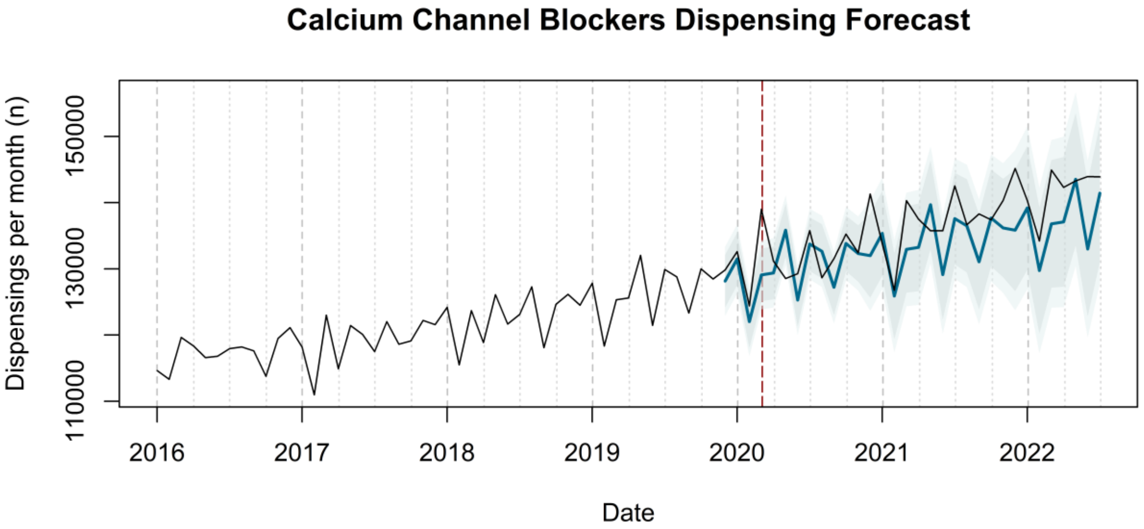

Calcium Channel Blockers Dispensing Forecast
Dispensings per month (n)
110000
130000
150000
2016
2017
2018
2019
2020
2021
2022
Date


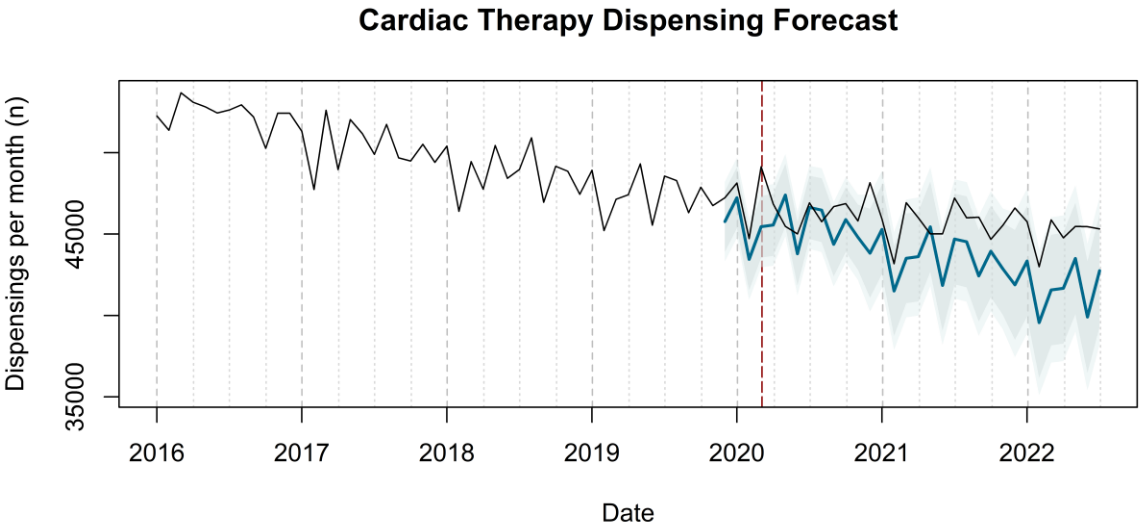

Cardiac Therapy Dispensing Forecast
Dispensings per month (n)
35000
45000
2016
2017
2018
2019
2020
2021
2022
Date

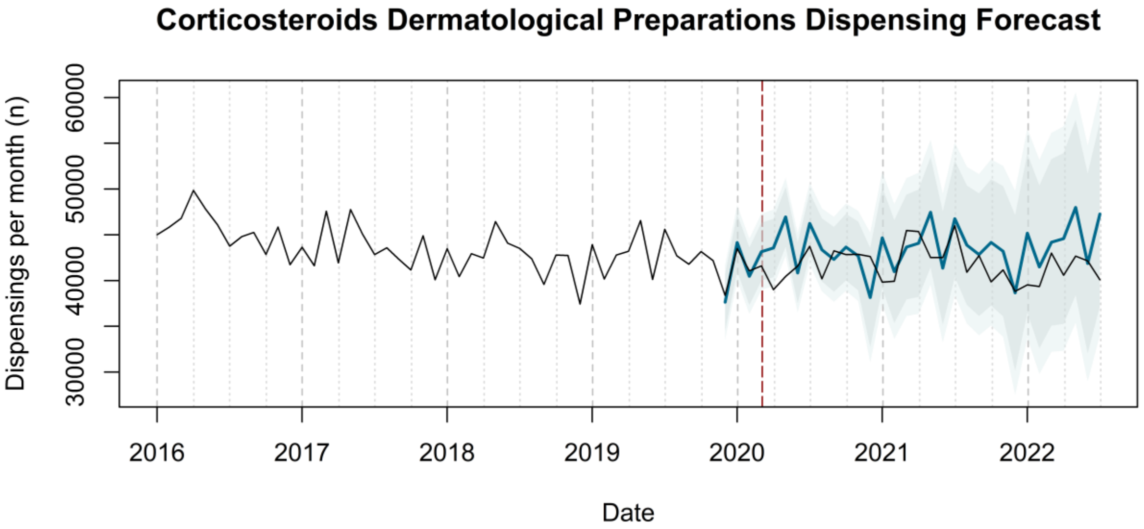
Corticosteroids Dermatological Preparations Dispensing Forecast
Dispensings per month (n)
30000
40000
50000
60000
2016
2017
2018
2019
2020
2021
2022
Date

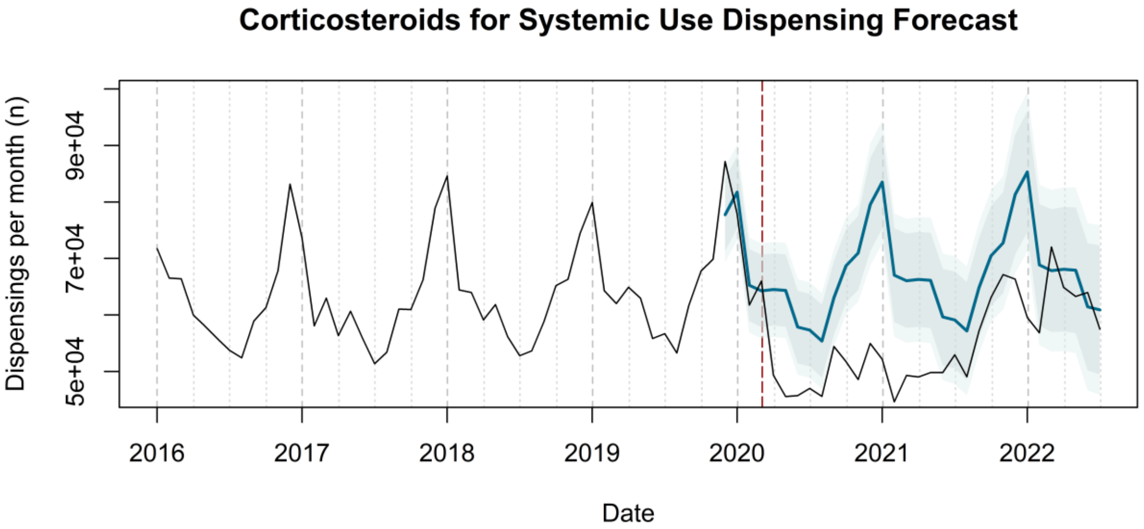
Corticosteroids for Systemic Use Dispensing Forecast
Dispensings per month (n)
5e+04
7e+04
9e+04
2016
2017
2018
2019
2020
2021
2022
Date

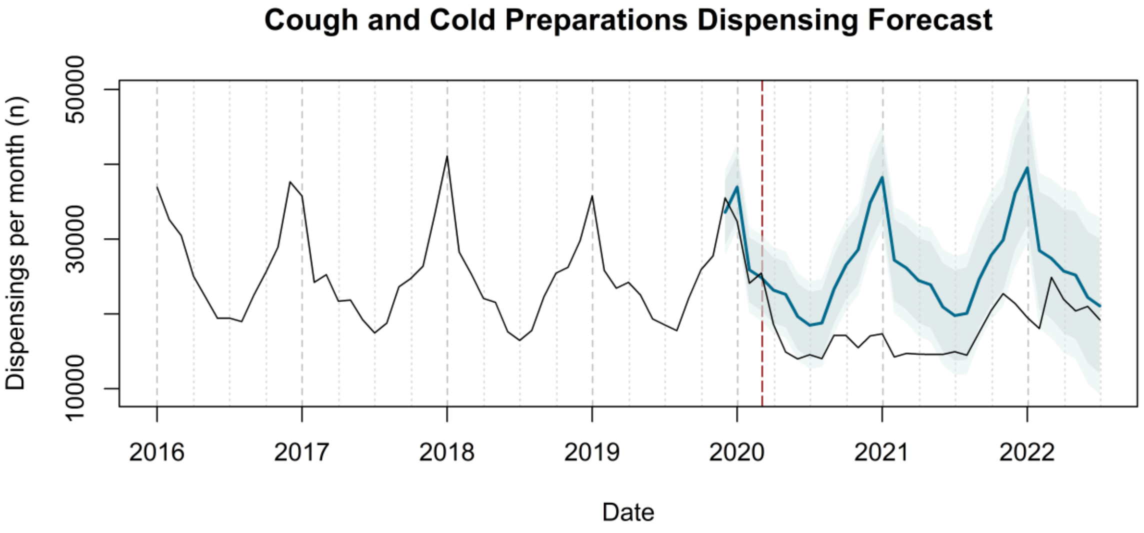

Cough and Cold Preparations Dispensing Forecast
Dispensings per month (n)
10000
30000
50000
2016
2017
2018
2019
2020
2021
2022
Date


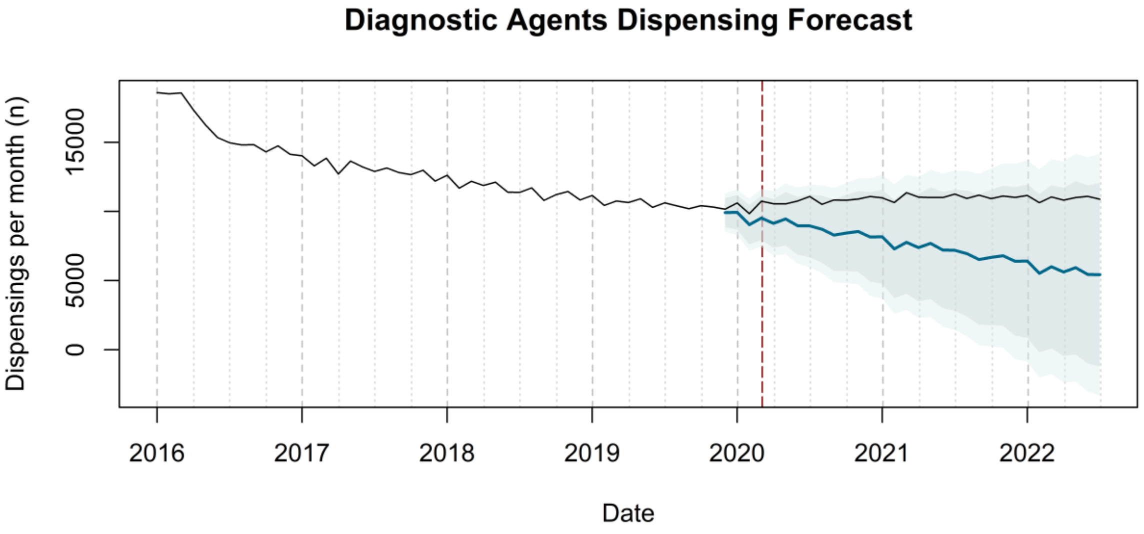

Diagnostic Agents Dispensing Forecast
Dispensings per month (n)
0
5000
15000
2016
2017
2018
2019
2020
2021
2022
Date


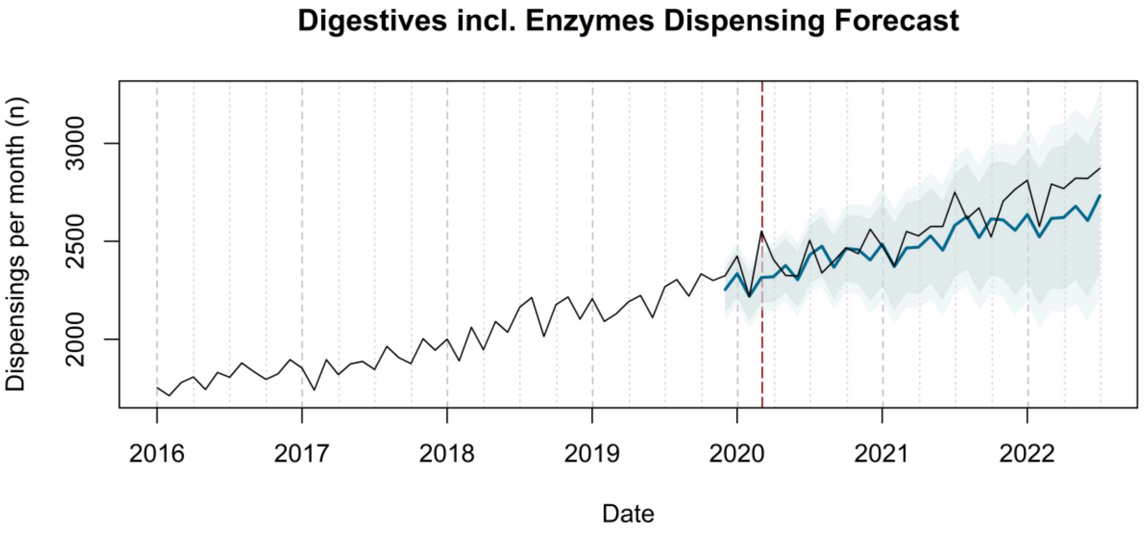

Digestives incl. Enzymes Dispensing Forecast
Dispensings per month (n)
2000
2500
3000
2016
2017
2018
2019
2020
2021
2022
Date

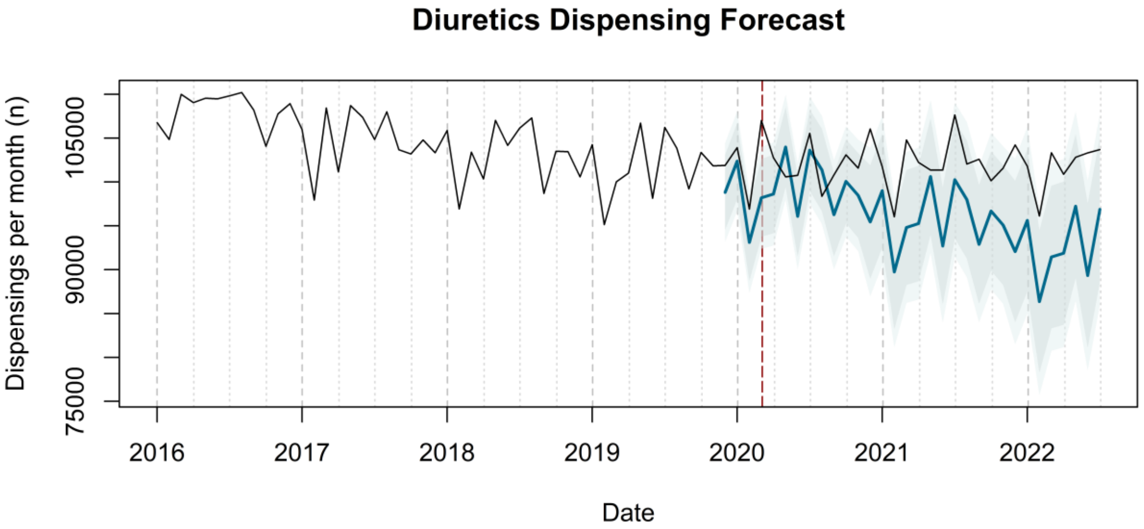

Diuretics Dispensing Forecast
Dispensings per month (n)
75000
90000
105000
2016
2017
2018
2019
2020
2021
2022
Date


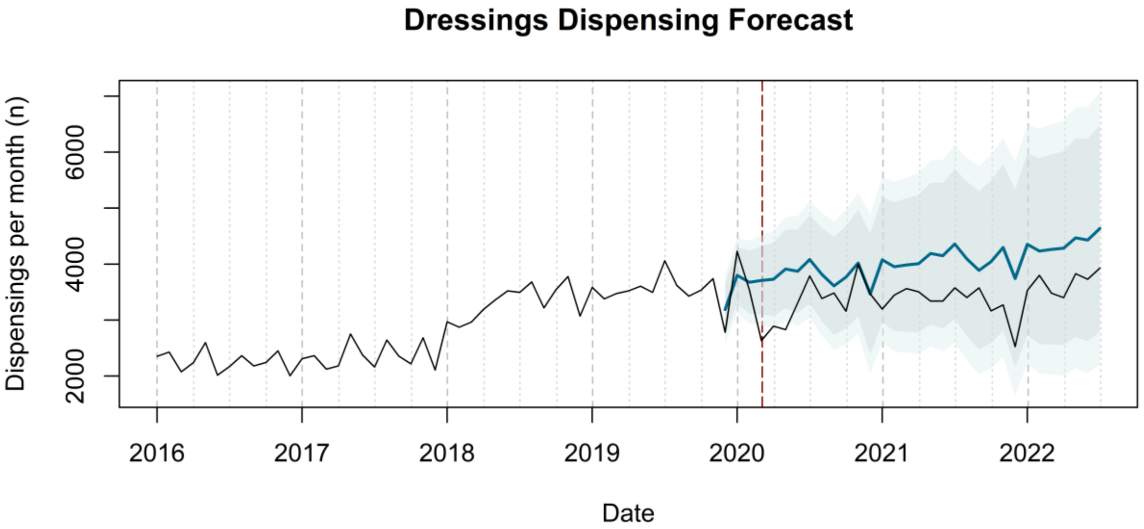

Dressings Dispensing Forecast
Dispensings per month (n)
2000
4000
6000
2016
2017
2018
2019
2020
2021
2022
Date


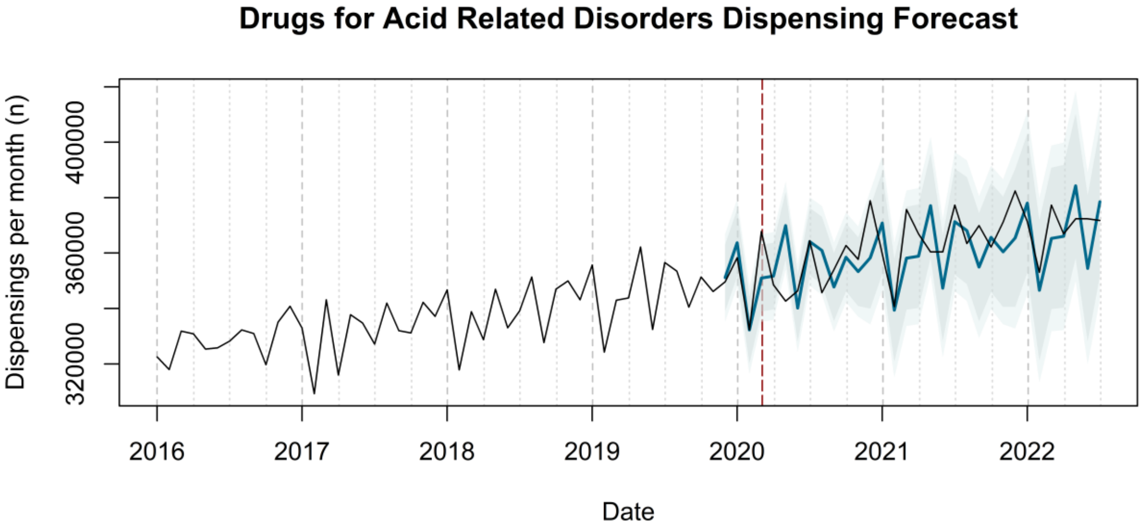

Drugs for Acid Related Disorders Dispensing Forecast
Dispensings per month (n)
320000
360000
400000
2016
2017
2018
2019
2020
2021
2022
Date

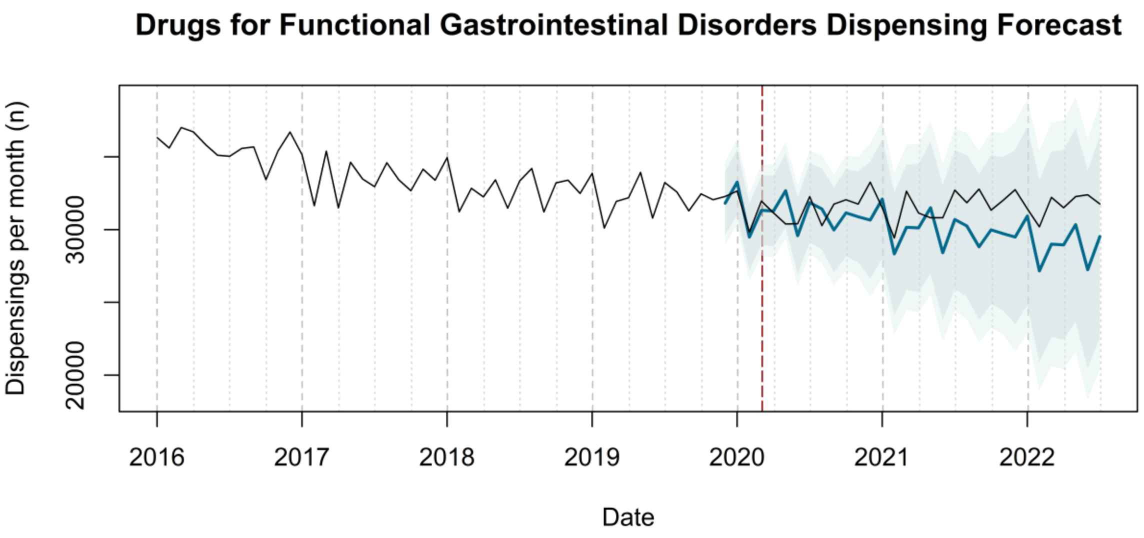
Drugs for Functional Gastrointestinal Disorders Dispensing Forecast
Dispensings per month (n)
20000
30000
2016
2017
2018
2019
2020
2021
2022
Date

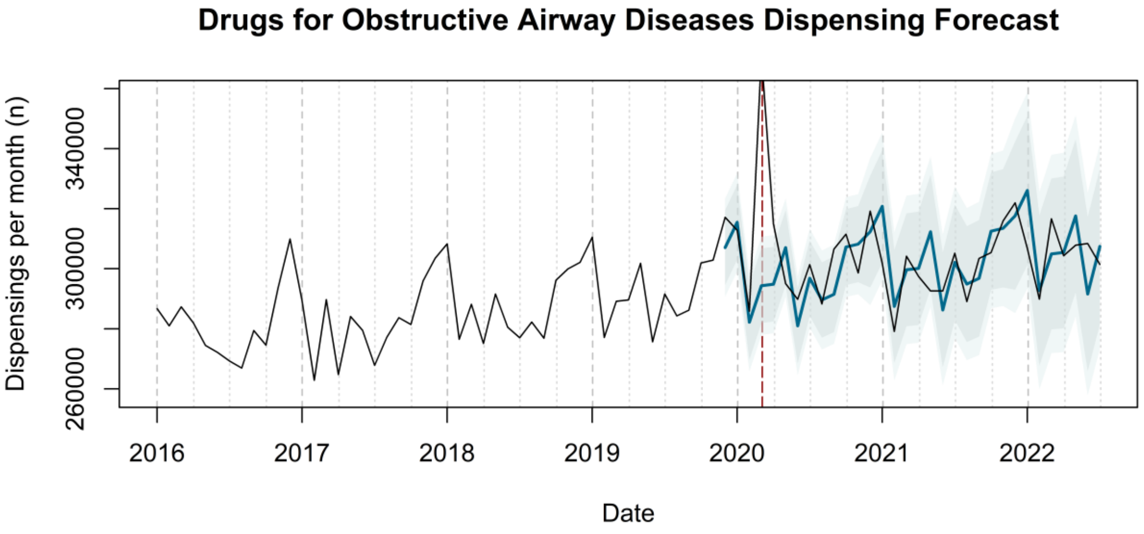
Drugs for Obstructive Airway Diseases Dispensing Forecast
Dispensings per month (n)
260000
300000
340000
2016
2017
2018
2019
2020
2021
2022
Date

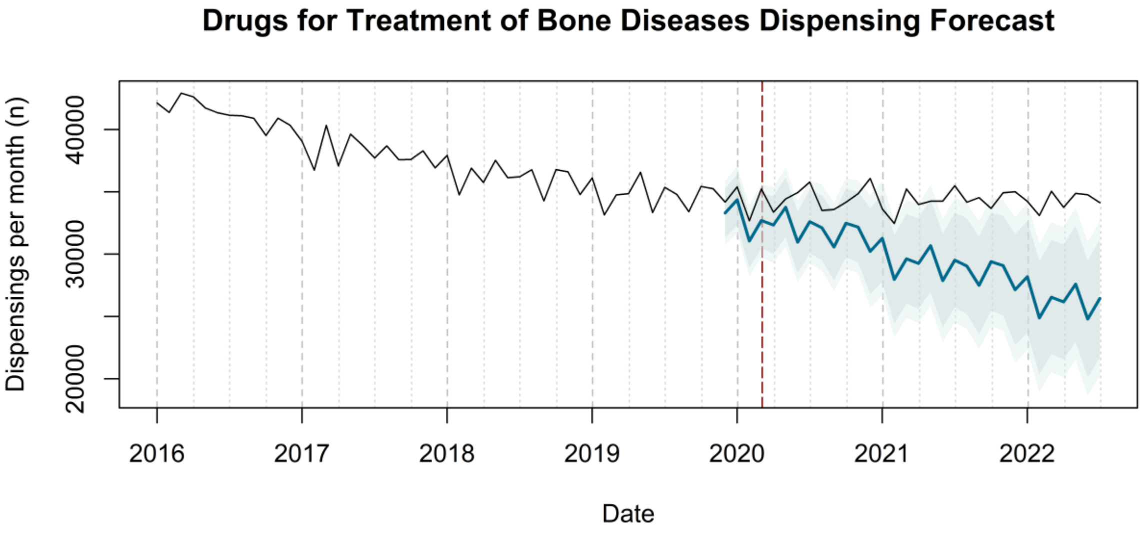
Drugs for Treatment of Bone Diseases Dispensing Forecast
Dispensings per month (n)
20000
30000
40000
2016
2017
2018
2019
2020
2021
2022
Date

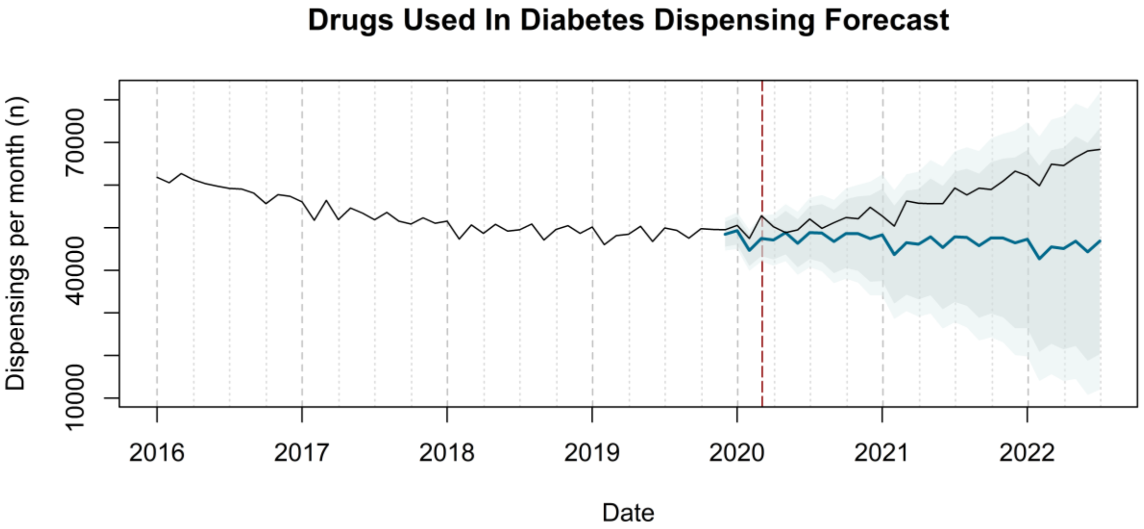
Drugs Used In Diabetes Dispensing Forecast
Dispensings per month (n)
10000
40000
70000
2016
2017
2018
2019
2020
2021
2022
Date

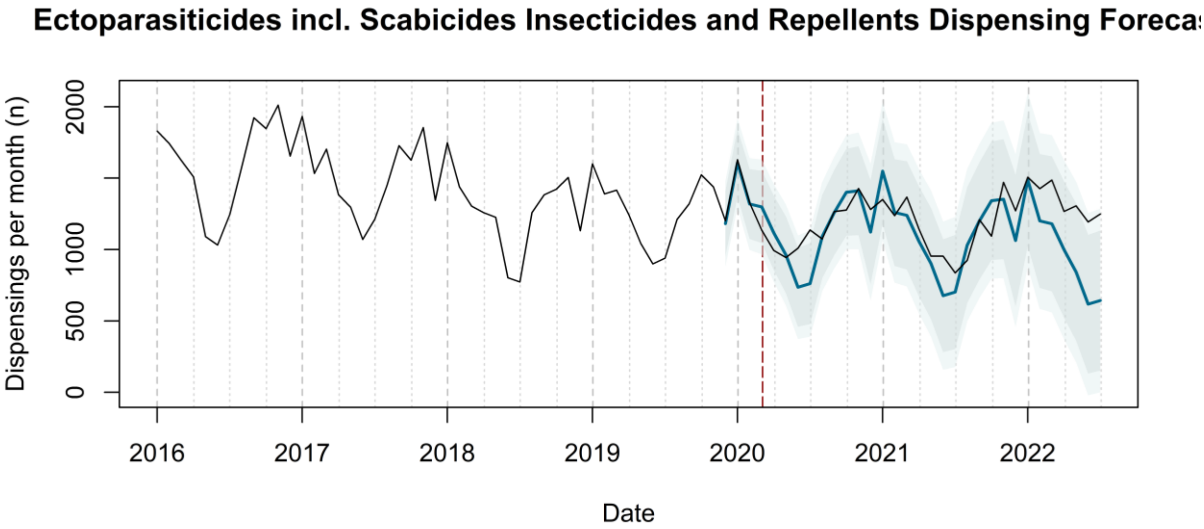
Ectoparasiticides incl. Scabicides Insecticides and Repellents Dispensing Foreca
Dispensings per month (n)
0
500
1000
2000
2016
2017
2018
2019
2020
2021
2022
Date

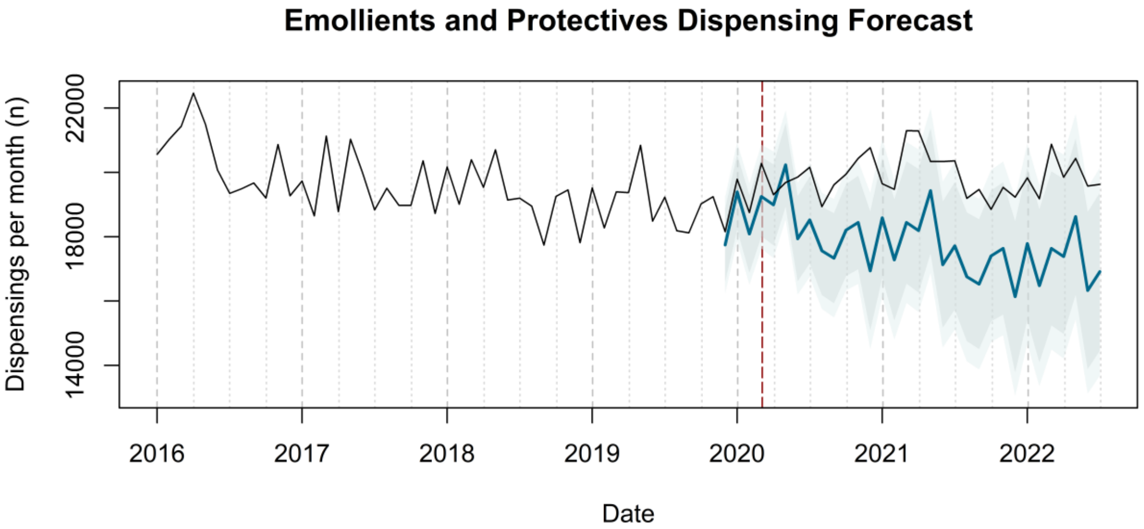
Emollients and Protectives Dispensing Forecast
Dispensings per month (n)
14000
18000
22000
2016
2017
2018
2019
2020
2021
2022
Date

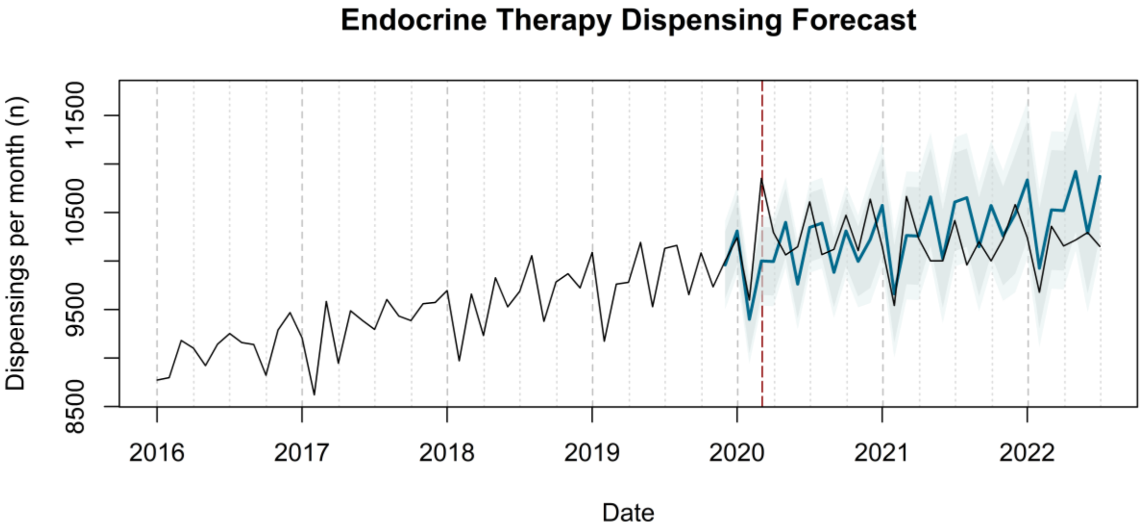

Endocrine Therapy Dispensing Forecast
Dispensings per month (n)
8500
9500
10500
11500
2016
2017
2018
2019
2020
2021
2022
Date


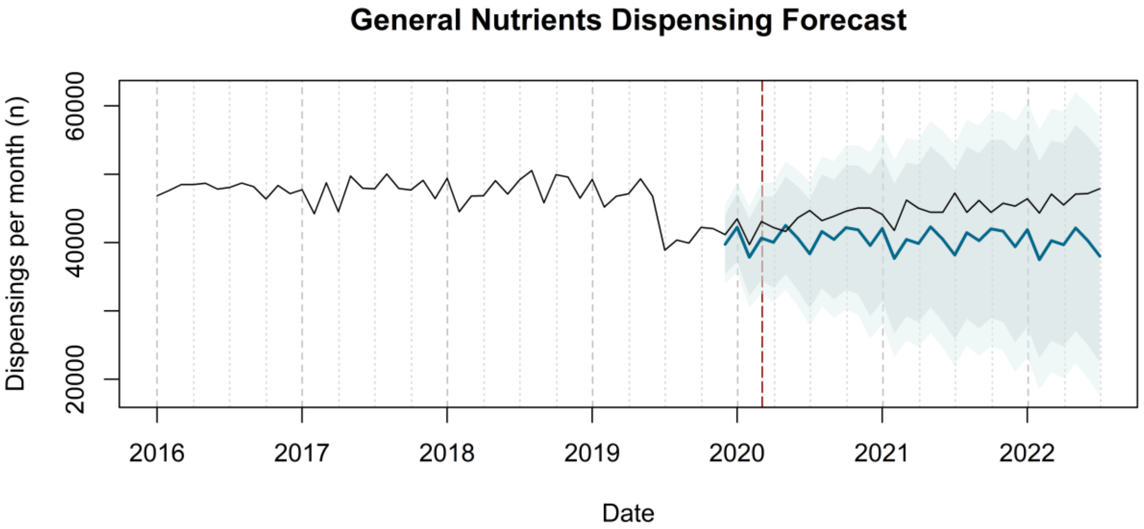

General Nutrients Dispensing Forecast
Dispensings per month (n)
20000
40000
60000
2016
2017
2018
2019
2020
2021
2022
Date


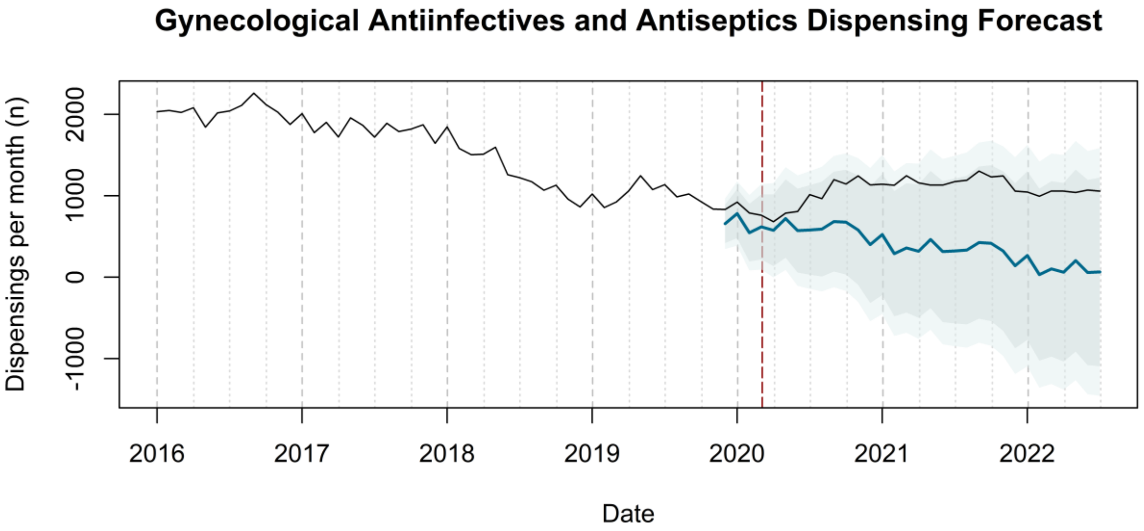

Gynecological Antiinfectives and Antiseptics Dispensing Forecast
Dispensings per month (n)
-1000
0
1000
2000
2016
2017
2018
2019
2020
2021
2022
Date

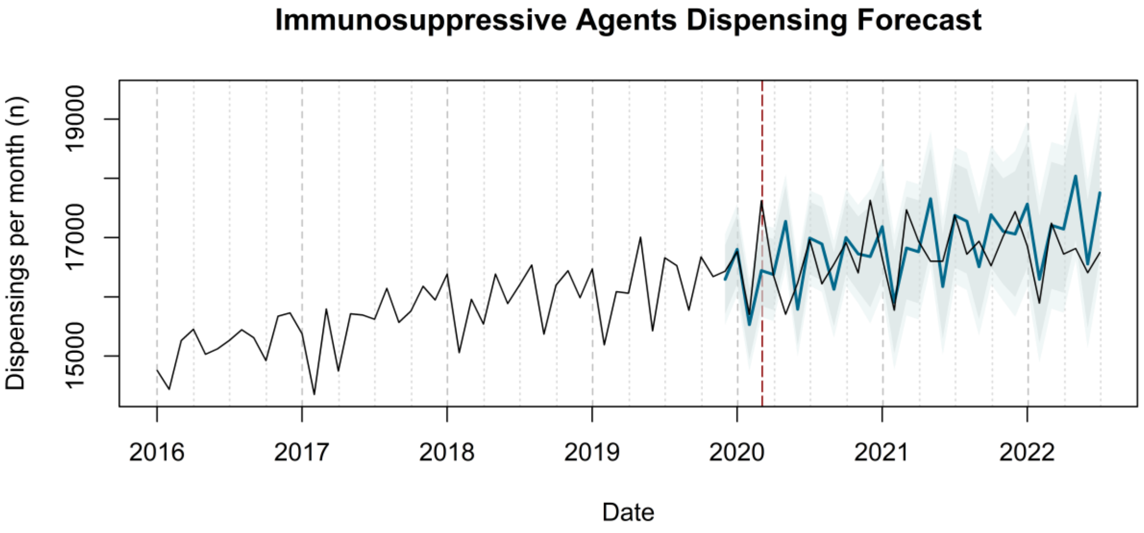
Immunosuppressive Agents Dispensing Forecast
Dispensings per month (n)
15000
17000
19000
2016
2017
2018
2019
2020
2021
2022
Date

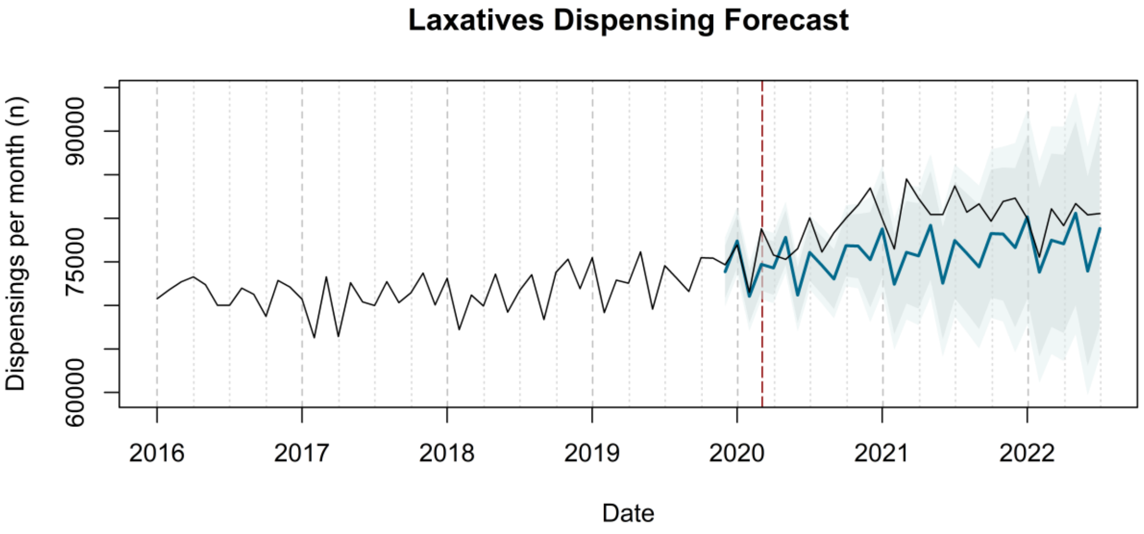
Laxatives Dispensing Forecast
Dispensings per month (n)
60000
75000
90000
2016
2017
2018
2019
2020
2021
2022
Date

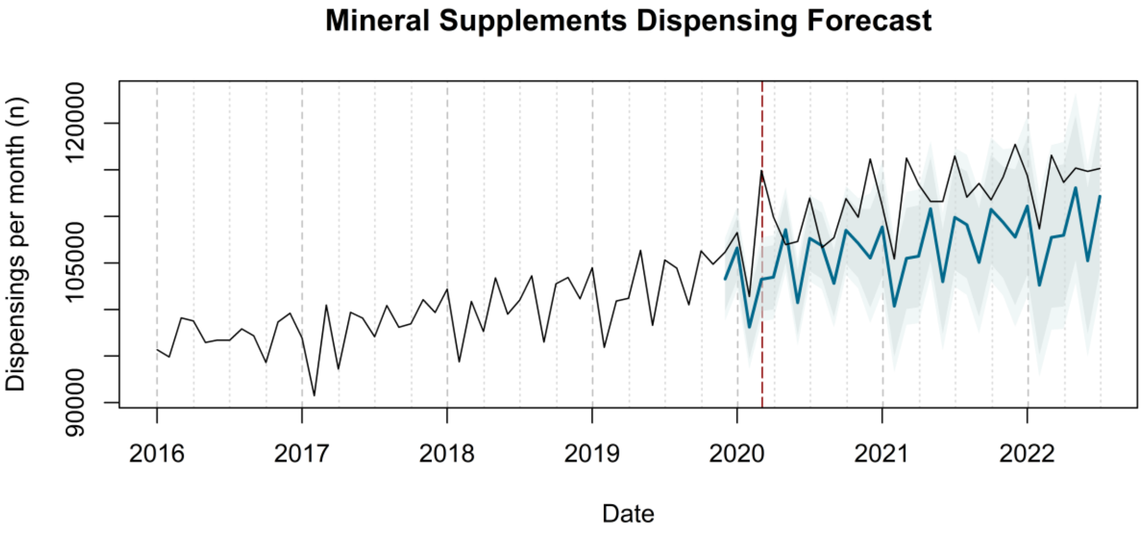
Mineral Supplements Dispensing Forecast
Dispensings per month (n)
90000
105000
120000
2016
2017
2018
2019
2020
2021
2022
Date

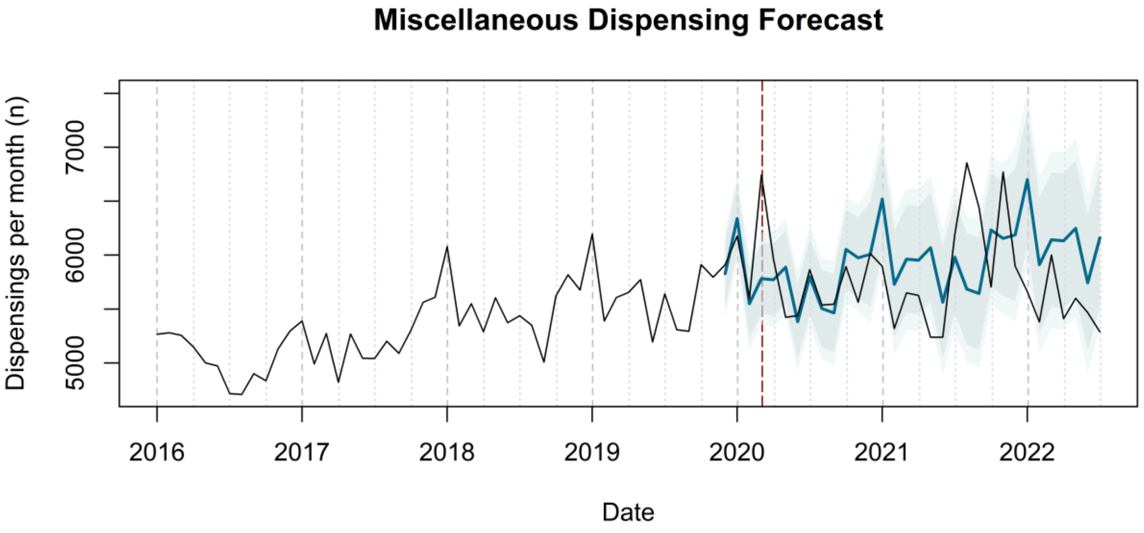
Miscellaneous Dispensing Forecast
Dispensings per month (n)
5000
6000
7000
2016
2017
2018
2019
2020
2021
2022
Date

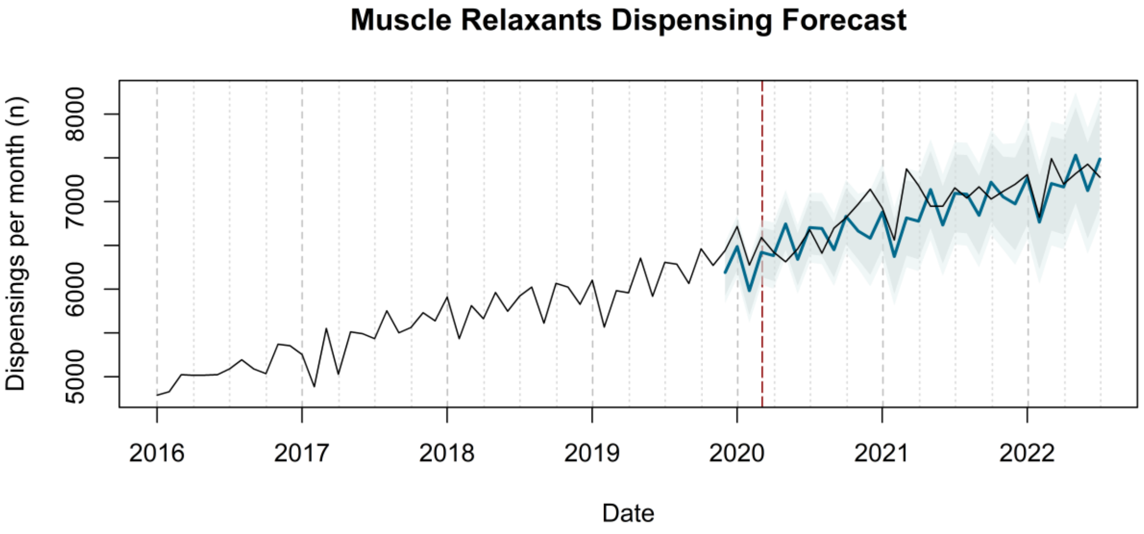
Muscle Relaxants Dispensing Forecast
Dispensings per month (n)
5000
6000
7000
8000
2016
2017
2018
2019
2020
2021
2022
Date

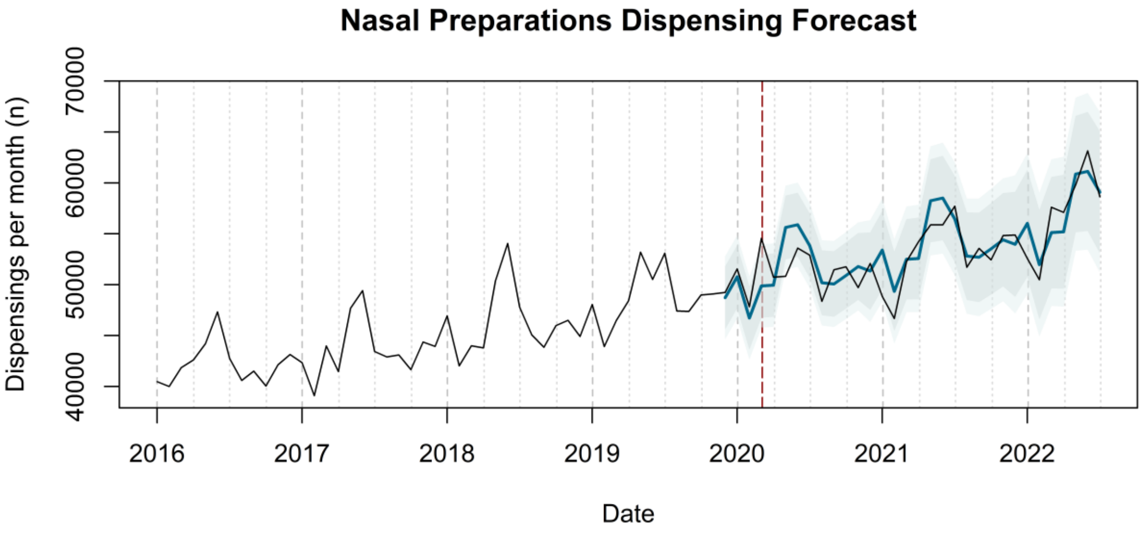
Nasal Preparations Dispensing Forecast
Dispensings per month (n)
40000
50000
60000
70000
2016
2017
2018
2019
2020
2021
2022
Date

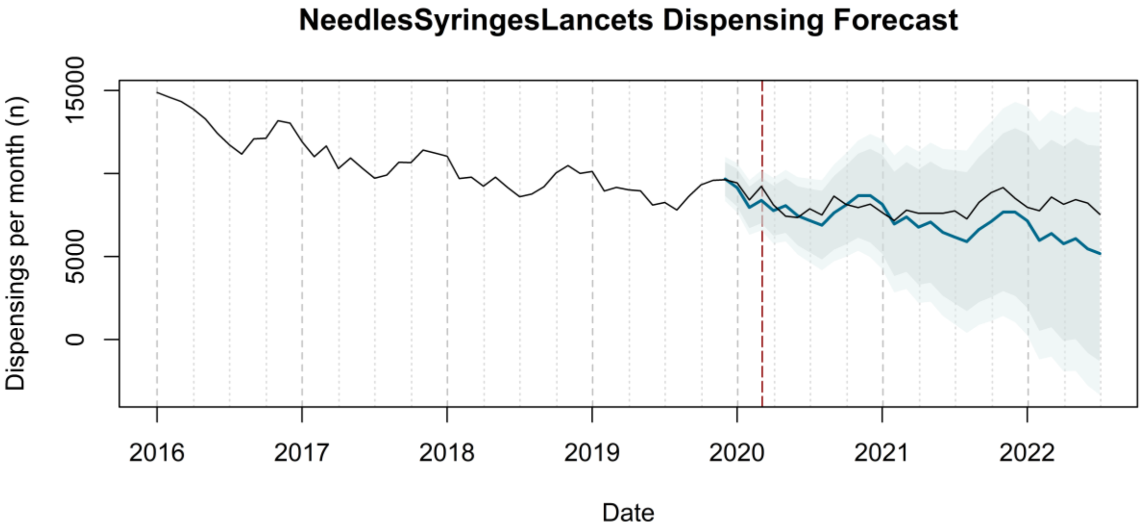
NeedlesSyringesLancets Dispensing Forecast
Dispensings per month (n)
15000
5000
0
2016
2017
2018
2019
2020
2021
2022
Date

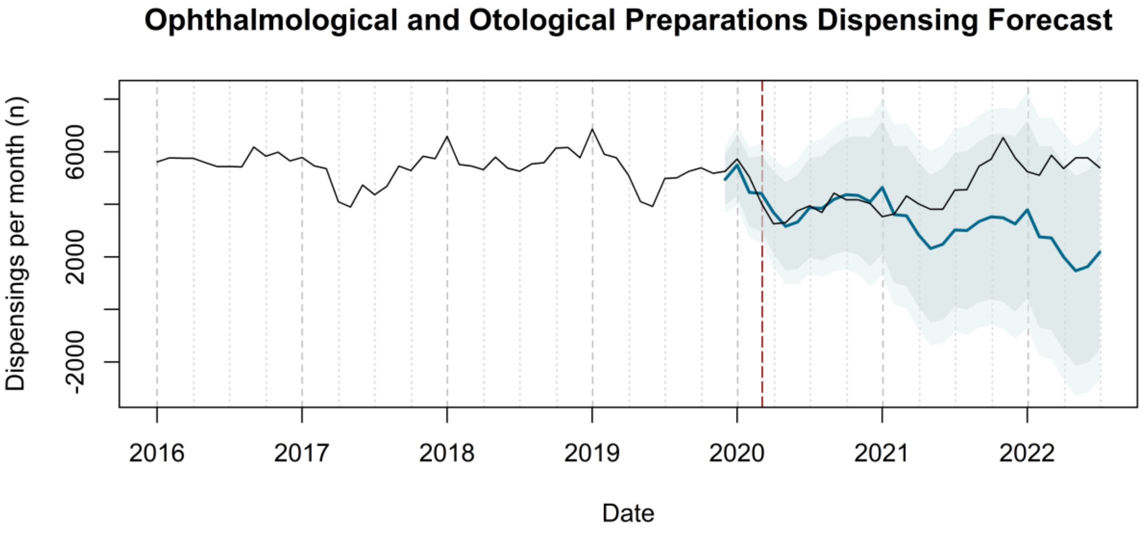
Ophthalmological and Otological Preparations Dispensing Forecast
Dispensings per month (n)
6000
2000
-2000
2016
2017
2018
2019
2020
2021
2022
Date

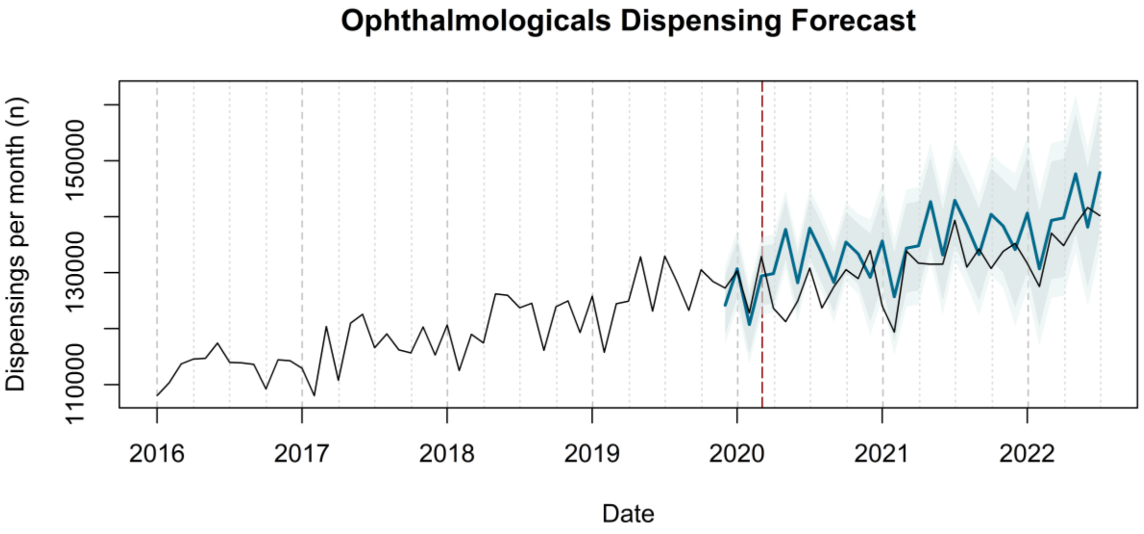
Ophthalmologicals Dispensing Forecast
Dispensings per month (n)
150000
130000
110000
2016
2017
2018
2019
2020
2021
2022
Date

Other Dermatological Preparations Dispensing Forecast

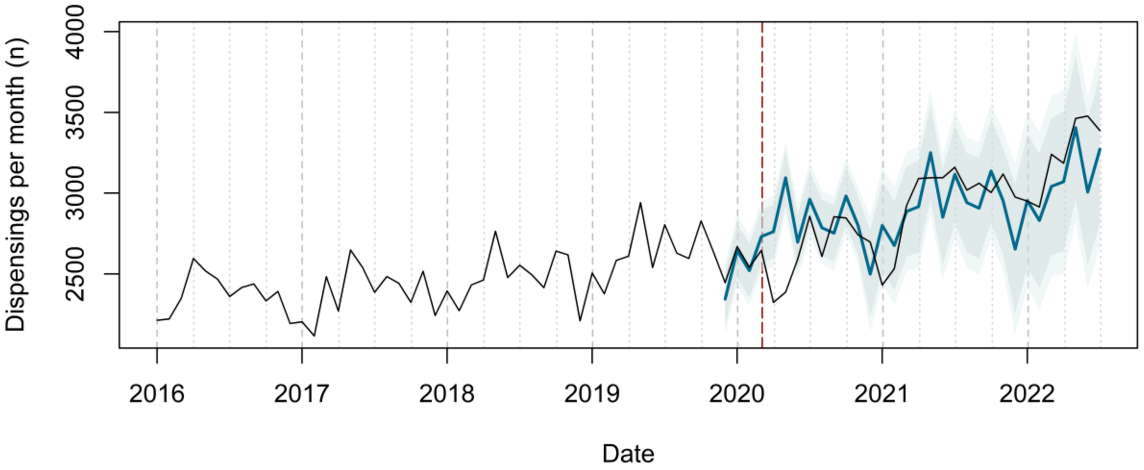


Other Drugs for Disorders of the Musculo-Skeletal System Dispensing Forecast

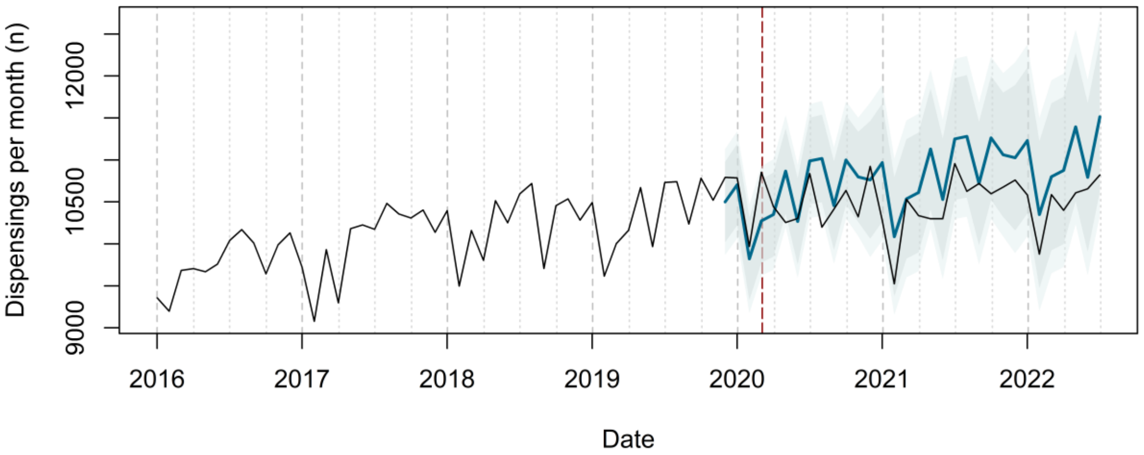


Other Gynecologicals Dispensing Forecast

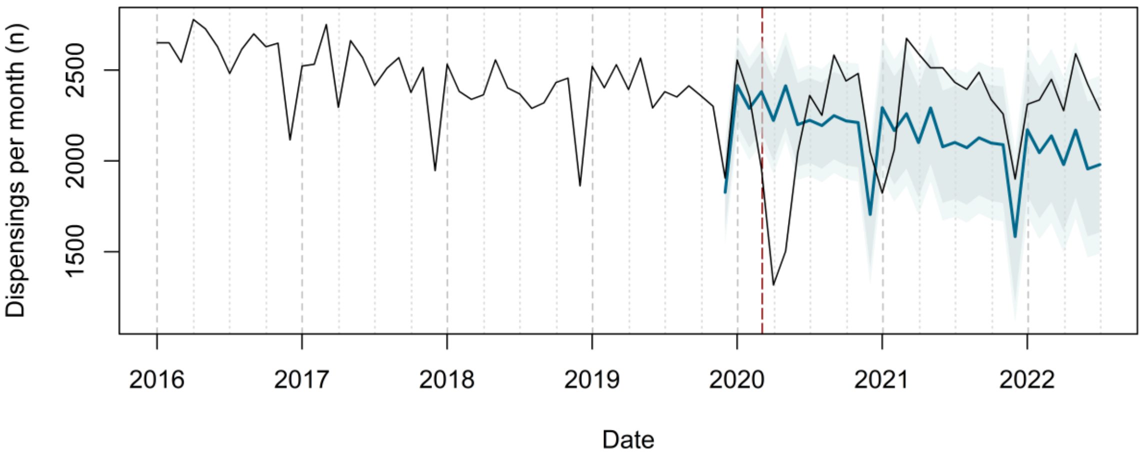

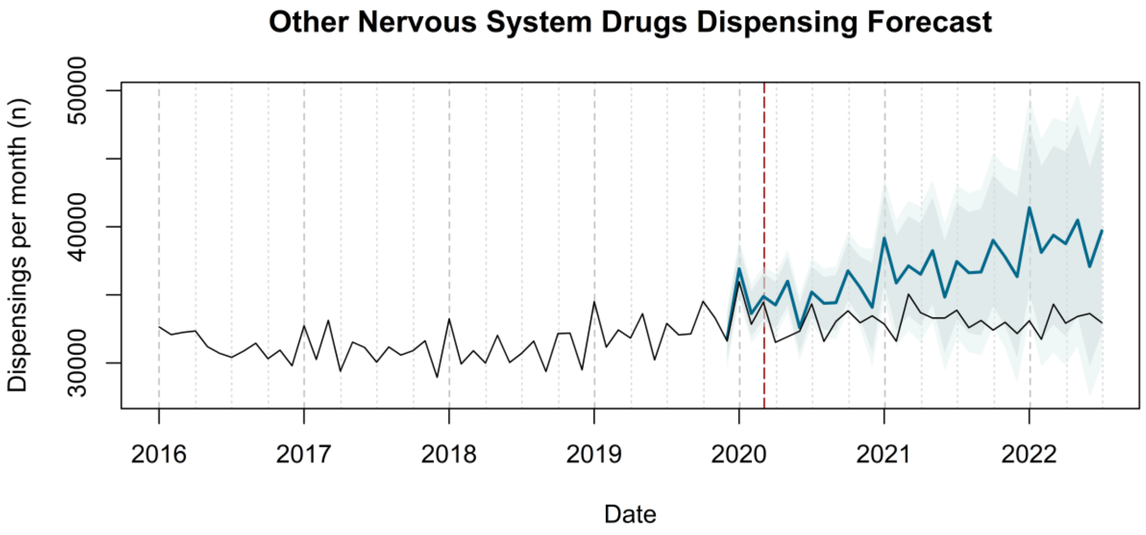
Other Nervous System Drugs Dispensing Forecast
Dispensings per month (n)
30000
40000
50000
2016
2017
2018
2019
2020
2021
2022
Date

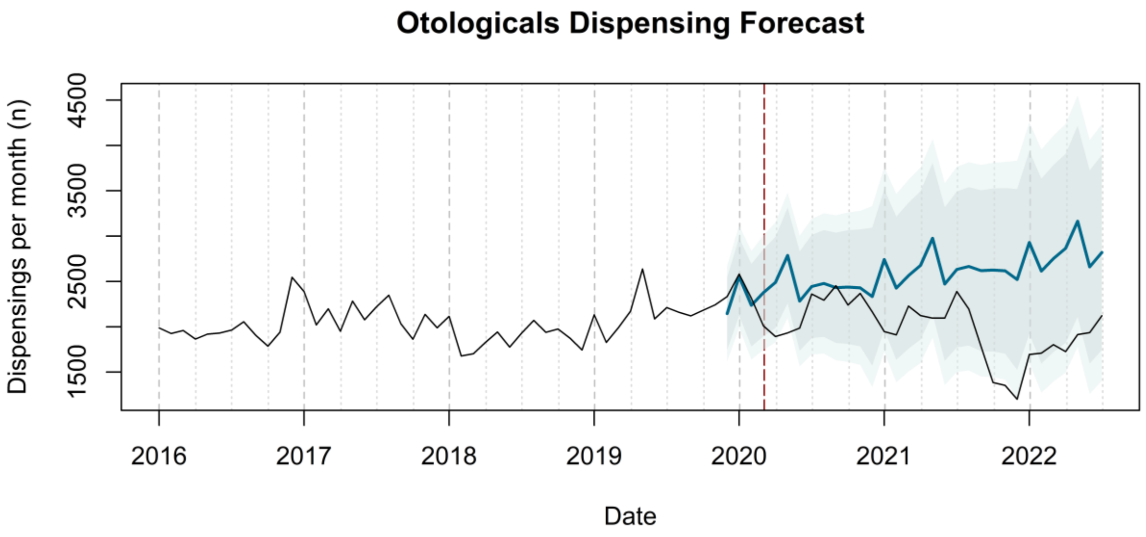
Otologicals Dispensing Forecast
Dispensings per month (n)
1500
2500
3500
4500
2016
2017
2018
2019
2020
2021
2022
Date

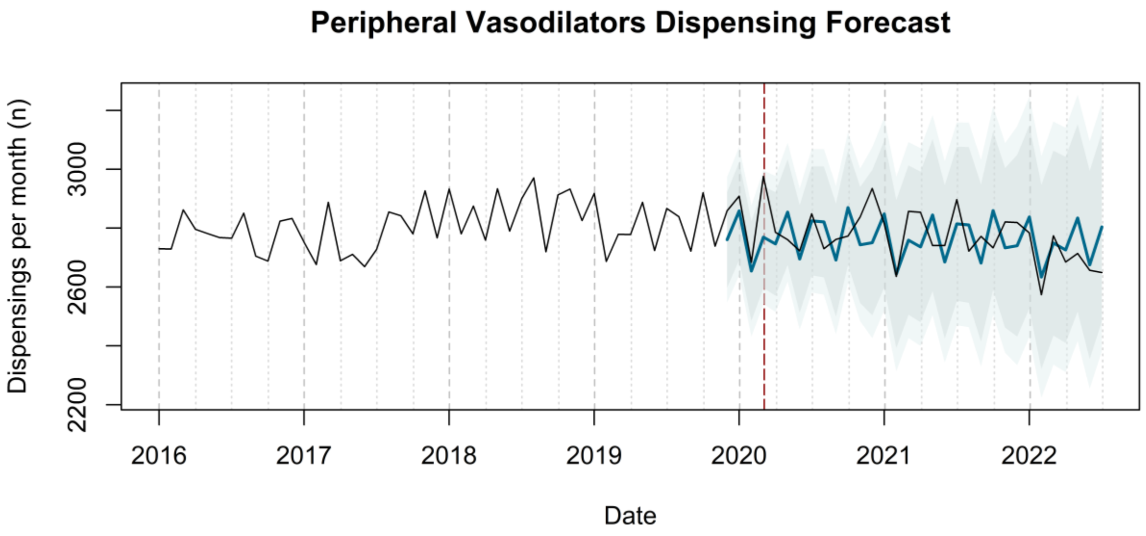
Peripheral Vasodilators Dispensing Forecast
Dispensings per month (n)
2200
2600
3000
2016
2017
2018
2019
2020
2021
2022
Date

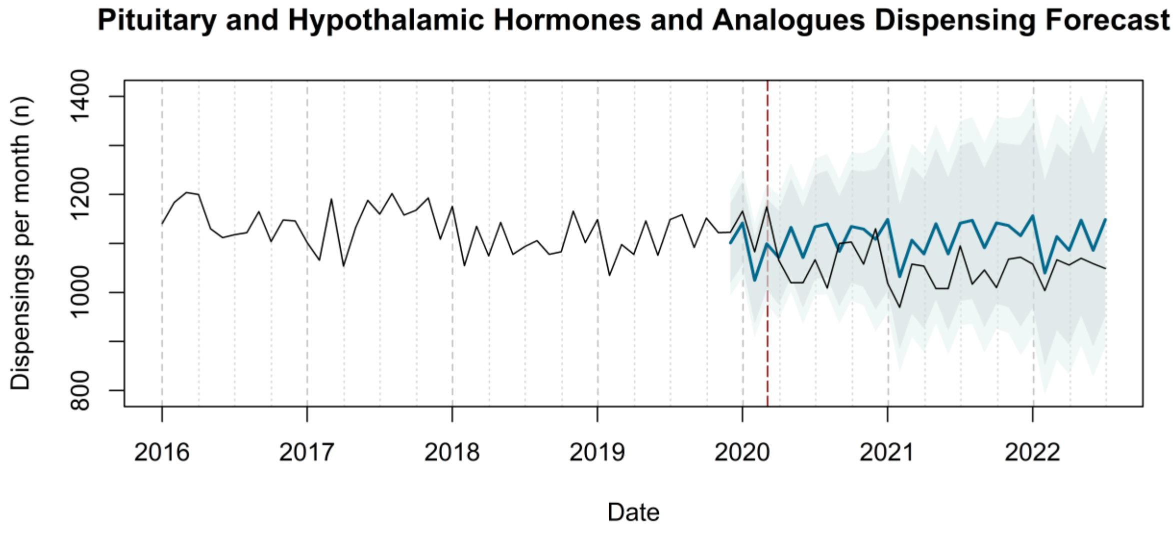

Pituitary and Hypothalamic Hormones and Analogues Dispensing Forecast
Dispensings per month (n)
800
1000
1200
1400
2016
2017
2018
2019
2020
2021
2022
Date


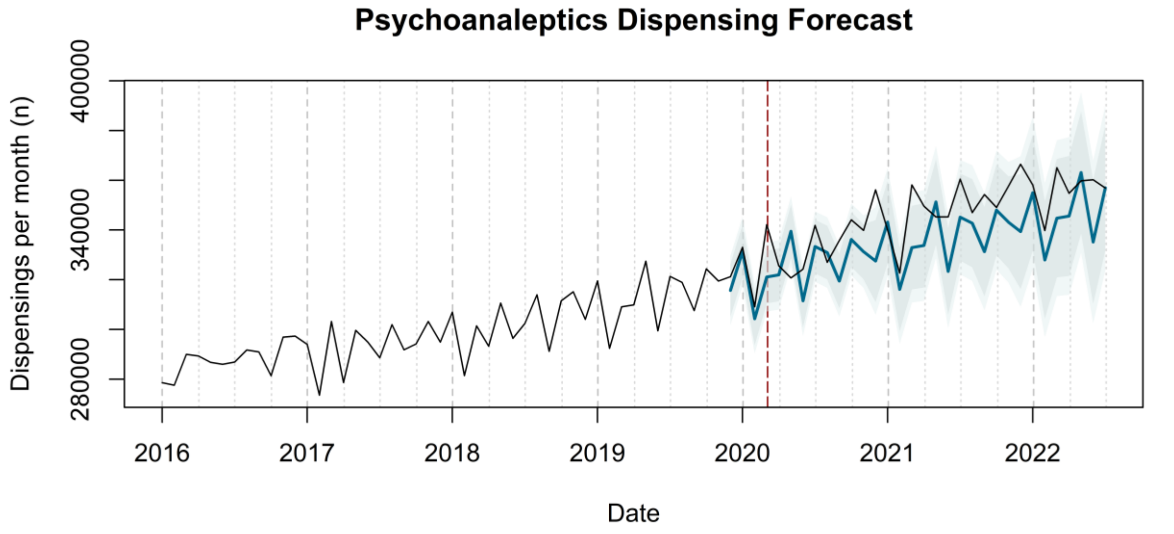

Psychoanaleptics Dispensing Forecast
Dispensings per month (n)
280000
340000
400000
2016
2017
2018
2019
2020
2021
2022
Date


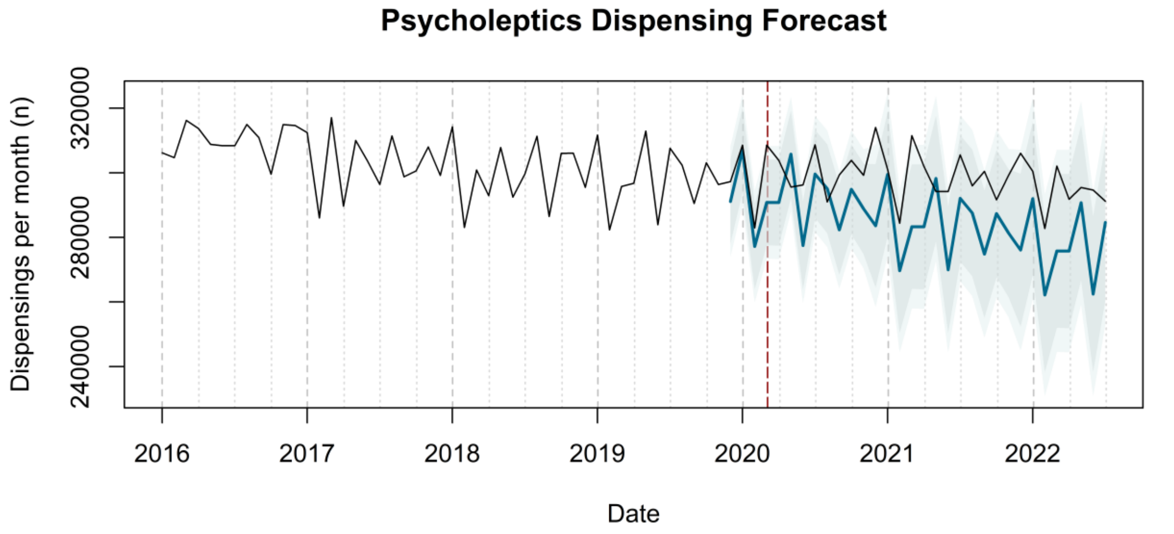

Psycholeptics Dispensing Forecast
Dispensings per month (n)
240000
280000
320000
2016
2017
2018
2019
2020
2021
2022
Date

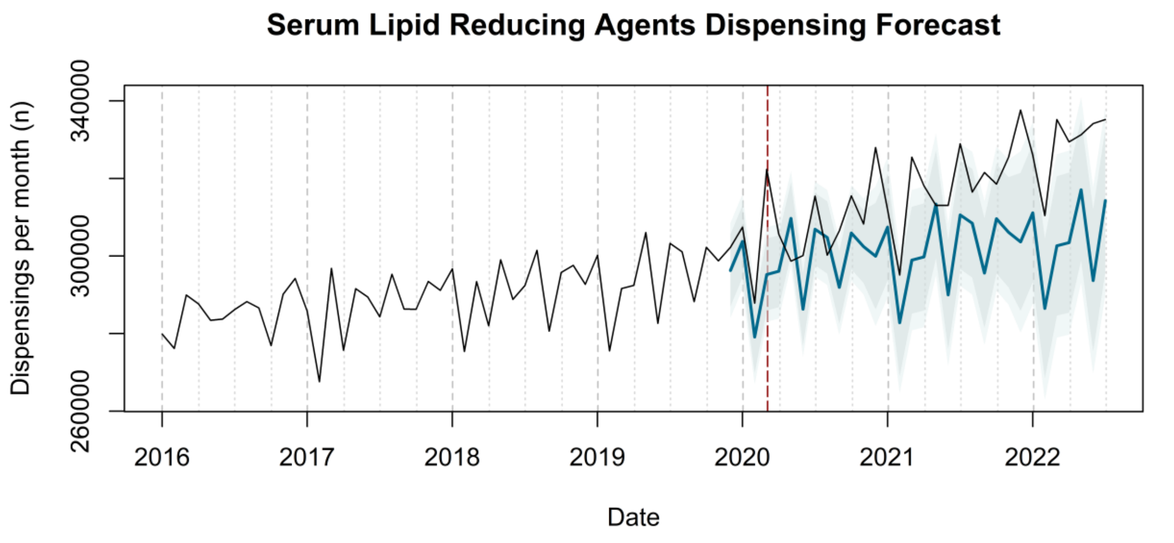

Serum Lipid Reducing Agents Dispensing Forecast
Dispensings per month (n)
260000
300000
340000
2016
2017
2018
2019
2020
2021
2022
Date


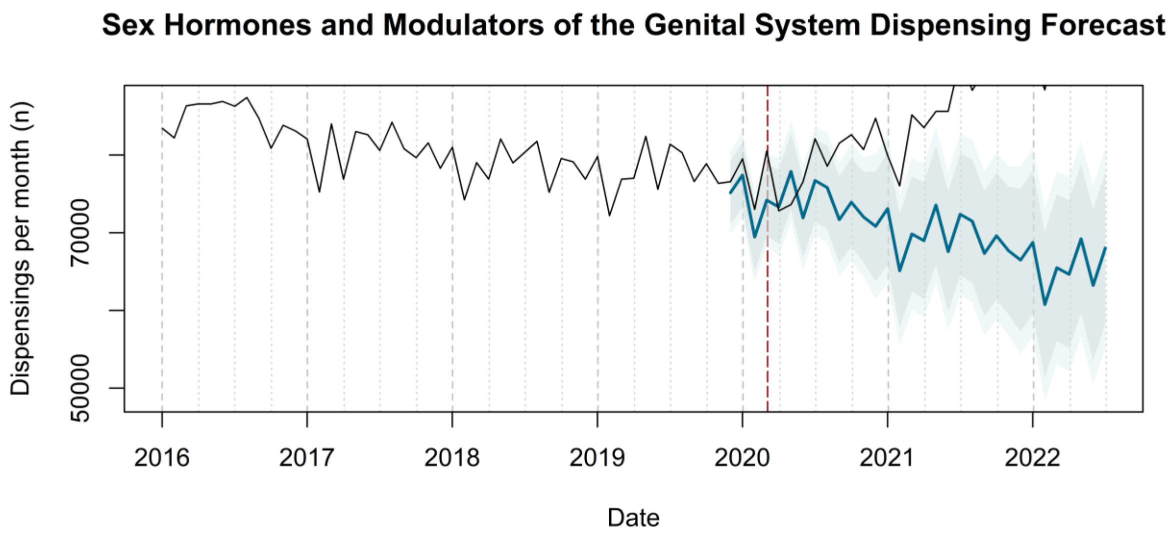

Sex Hormones and Modulators of the Genital System Dispensing Forecast
Dispensings per month (n)
50000
70000
2016
2017
2018
2019
2020
2021
2022
Date


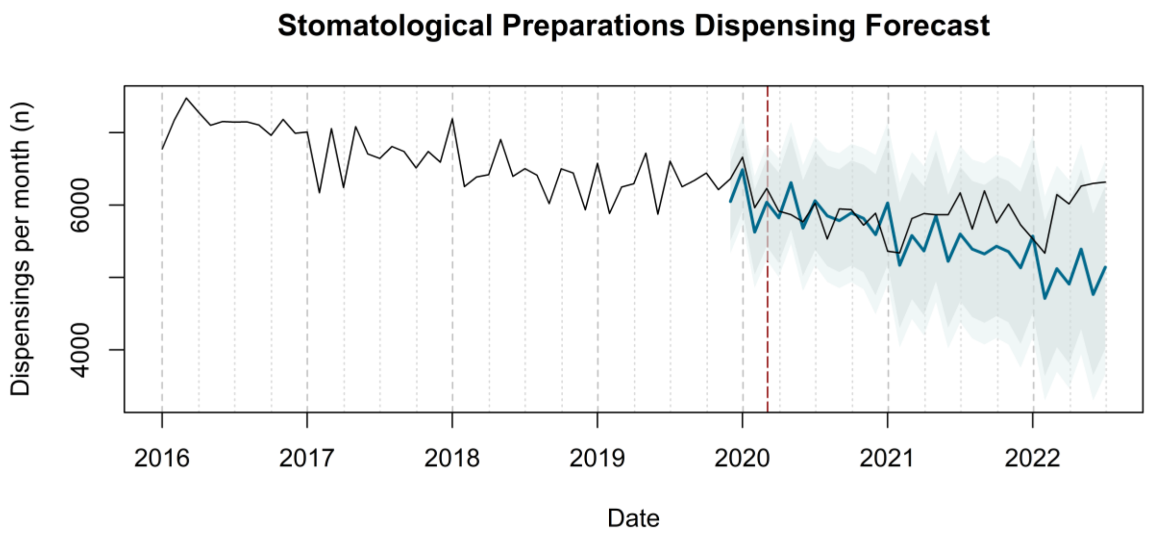

Stomatological Preparations Dispensing Forecast
Dispensings per month (n)
4000
6000
2016
2017
2018
2019
2020
2021
2022
Date

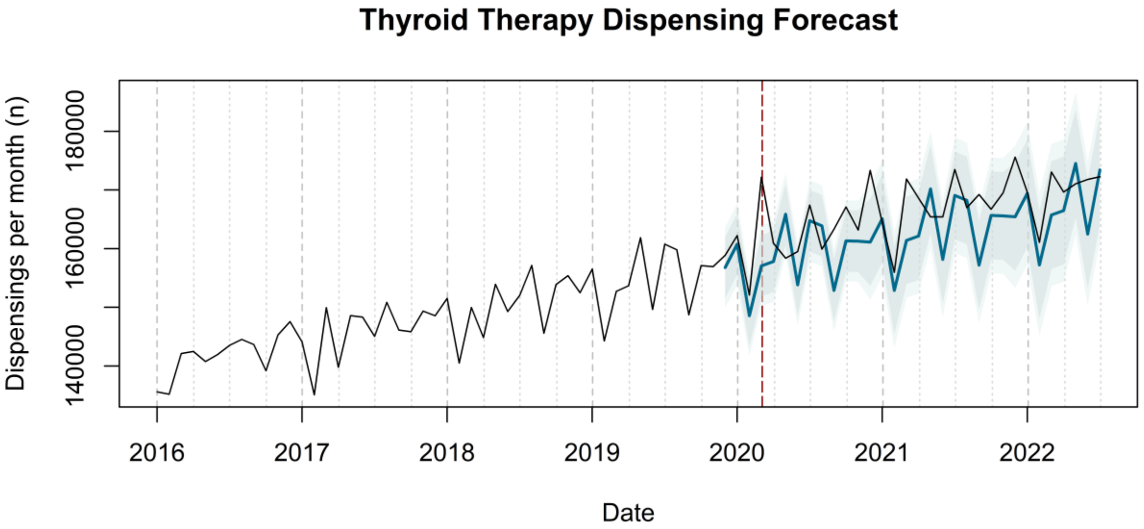
Thyroid Therapy Dispensing Forecast
Dispensings per month (n)
140000
160000
180000
2016
2017
2018
2019
2020
2021
2022
Date

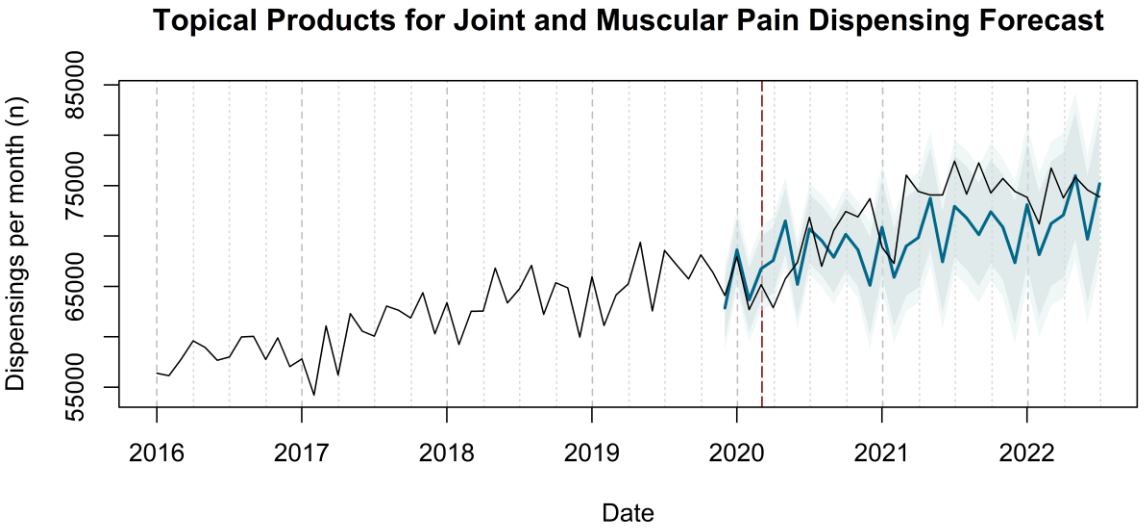
Topical Products for Joint and Muscular Pain Dispensing Forecast
Dispensings per month (n)
55000
65000
75000
85000
2016
2017
2018
2019
2020
2021
2022
Date

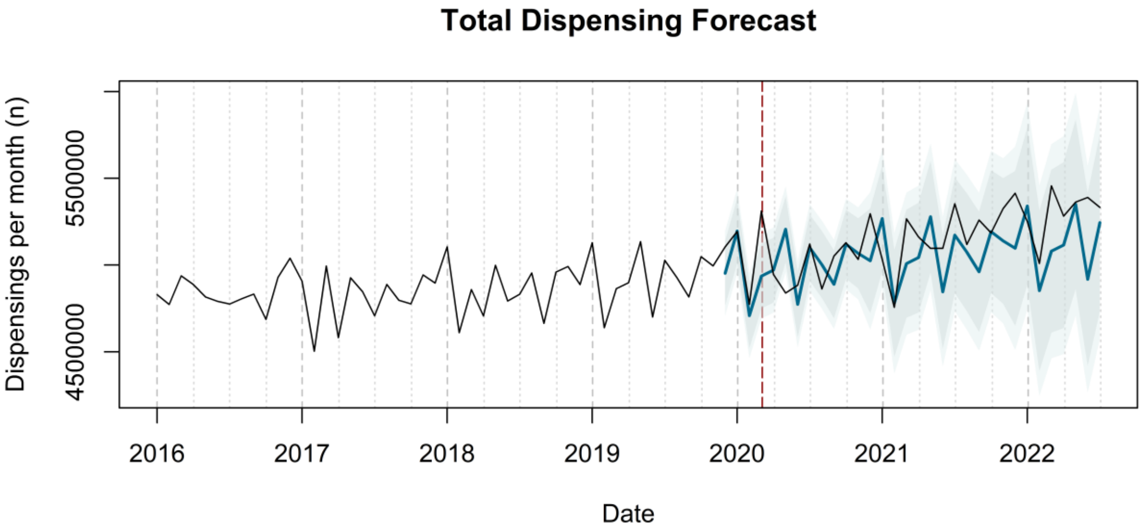
Total Dispensing Forecast
Dispensings per month (n)
4500000
5500000
2016
2017
2018
2019
2020
2021
2022
Date

**Urinary Requisites Dispensing Forecast**

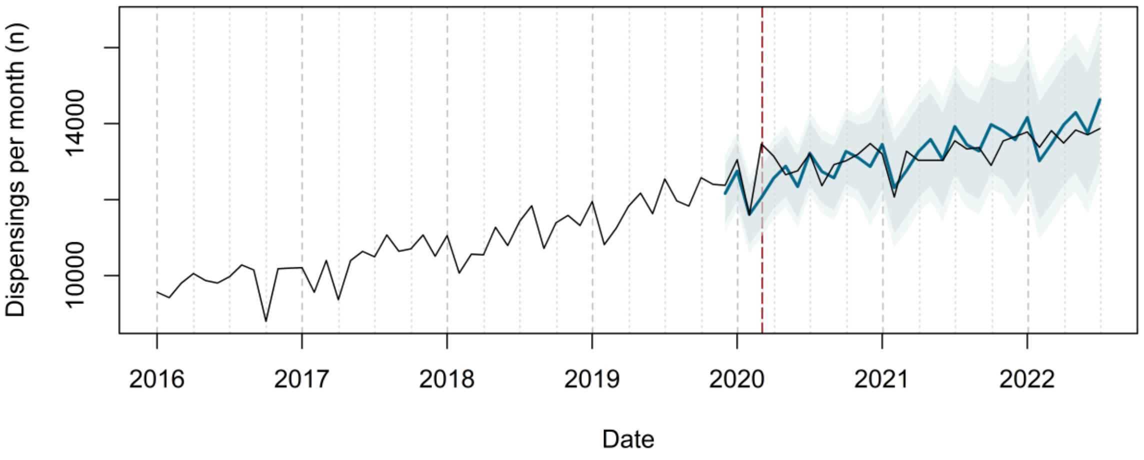


**Urologicals Dispensing Forecast**

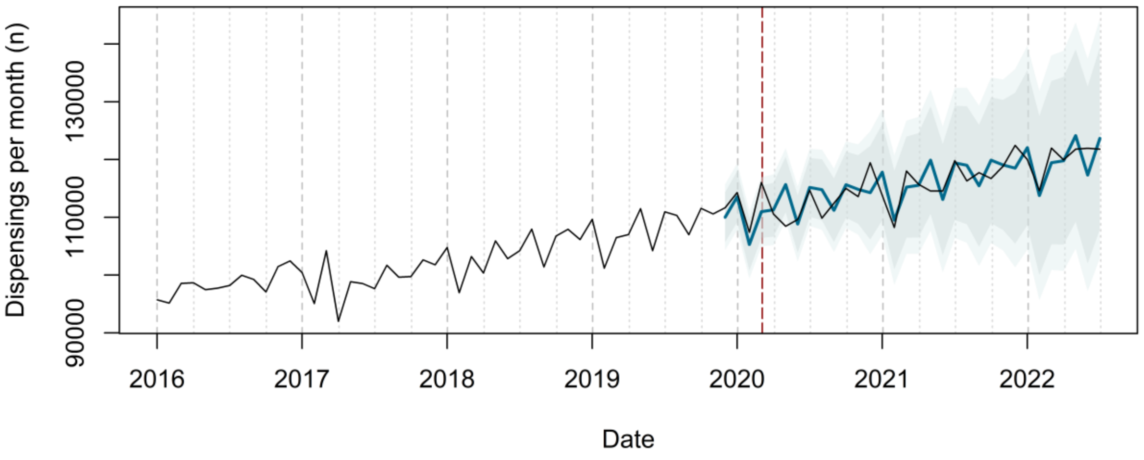

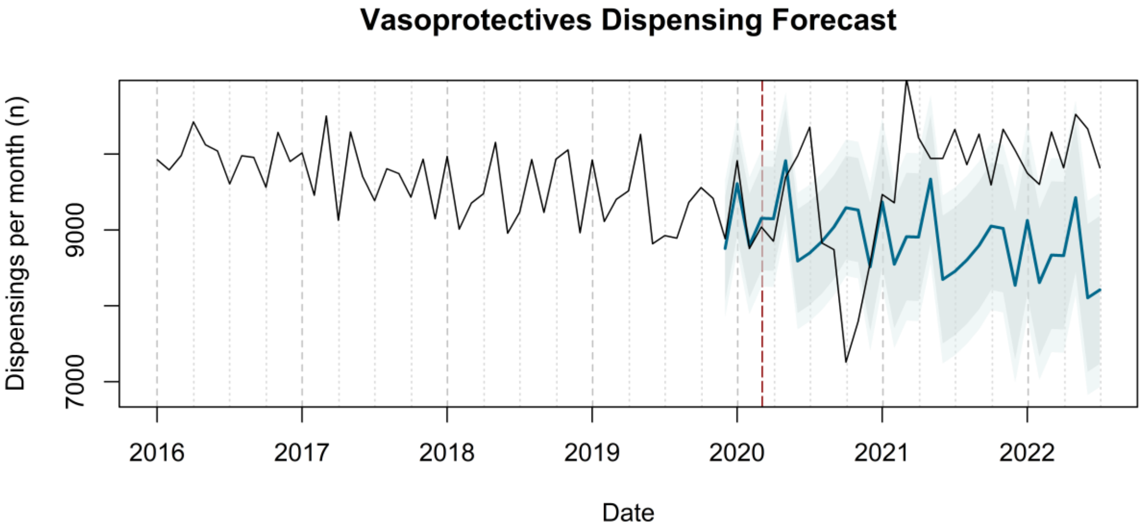

Vasoprotectives Dispensing Forecast
Dispensings per month (n)
7000
9000
2016
2017
2018
2019
2020
2021
2022
Date


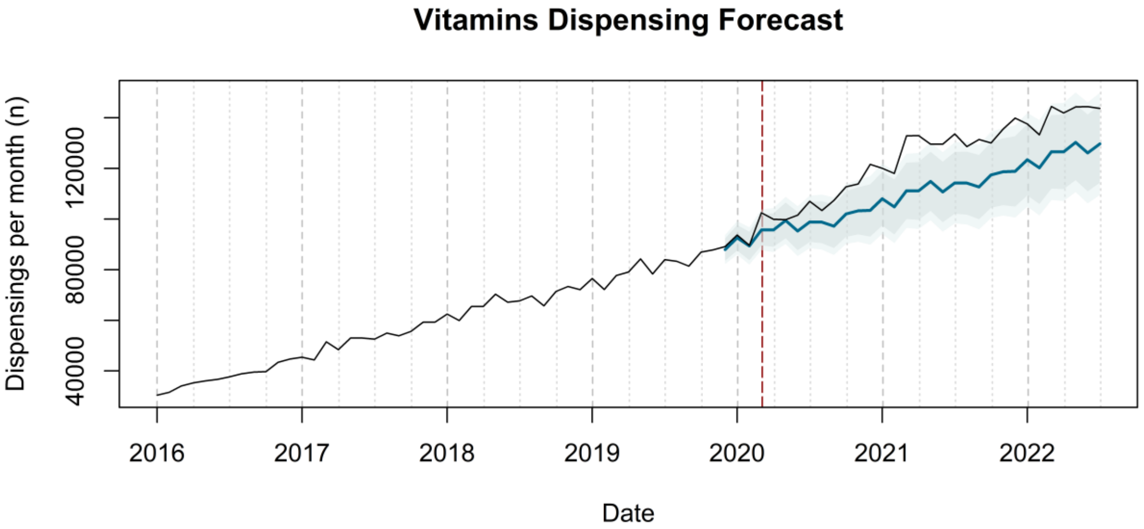

Vitamins Dispensing Forecast
Dispensings per month (n)
40000
80000
120000
2016
2017
2018
2019
2020
2021
2022
Date
