## Supplementary figure 2 for "Trends in medication use after the onset of the COVID-19 pandemic in the Republic of Ireland: an interrupted time series study"

**Supplementary figure 1.** Actual dispensing (black) and forecasted dispensing (blue, with 95% and 99% prediction intervals) for all therapeutic subgroups

*Therapeutic subgroups appear in alphabetical order by subgroup name*

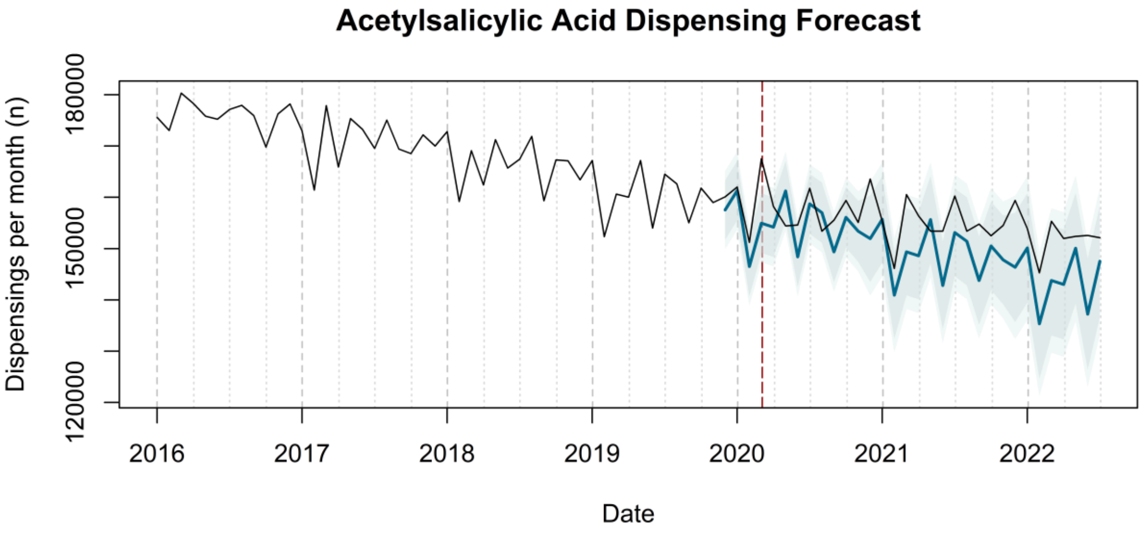


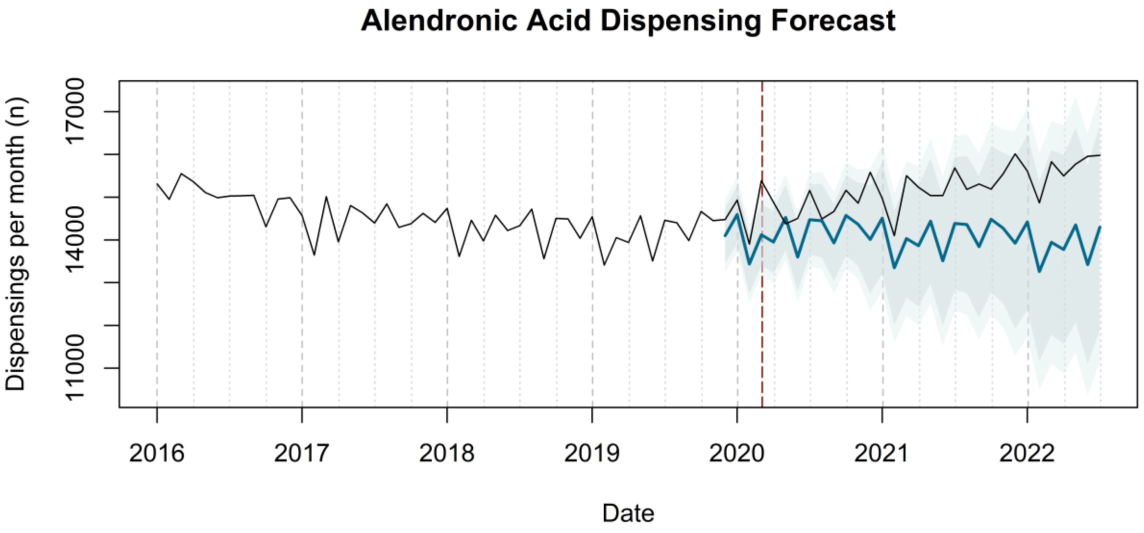

**Alginic Acid Dispensing Forecast**

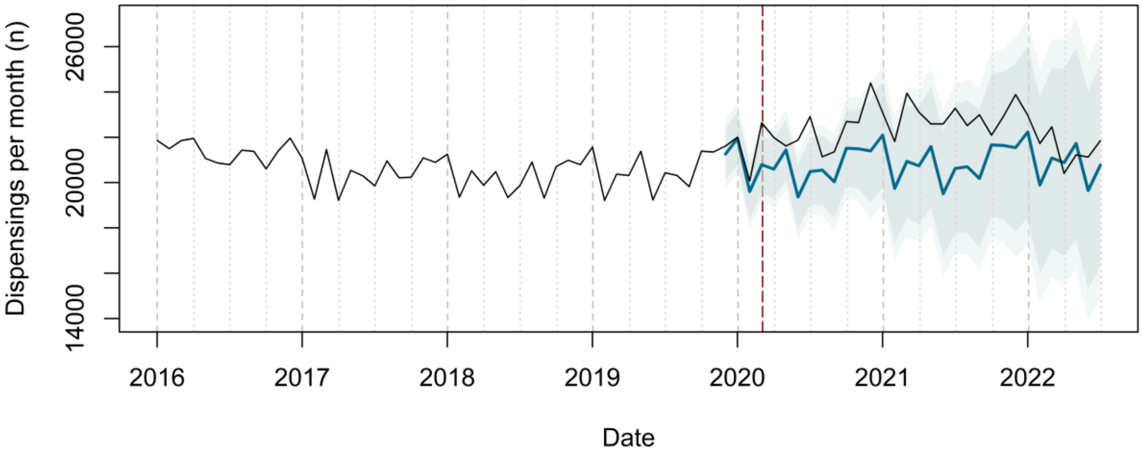


**Allopurinol Dispensing Forecast**

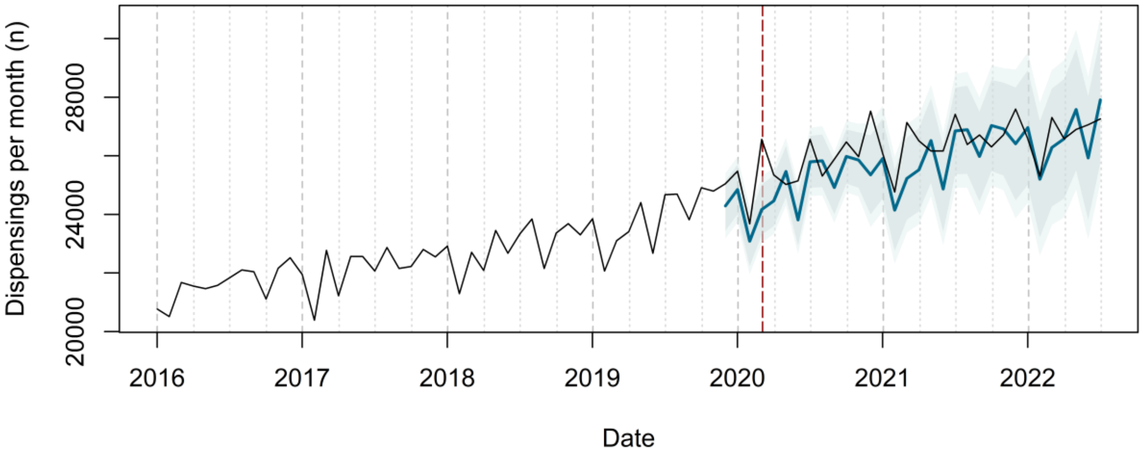


**Alprazolam Dispensing Forecast**

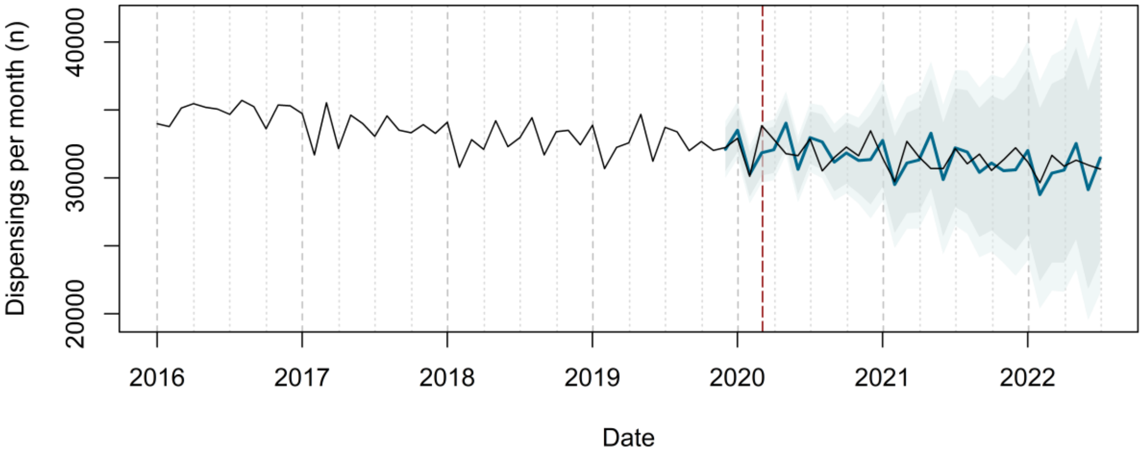

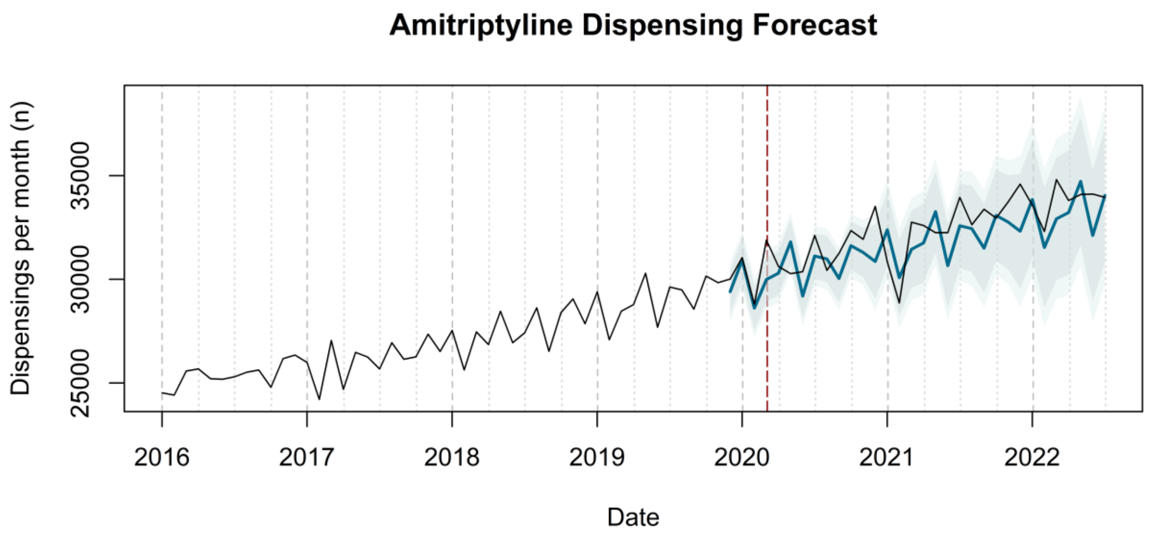

Amitriptyline Dispensing Forecast
Dispensings per month (n)
25000
30000
35000
2016
2017
2018
2019
2020
2021
2022
Date


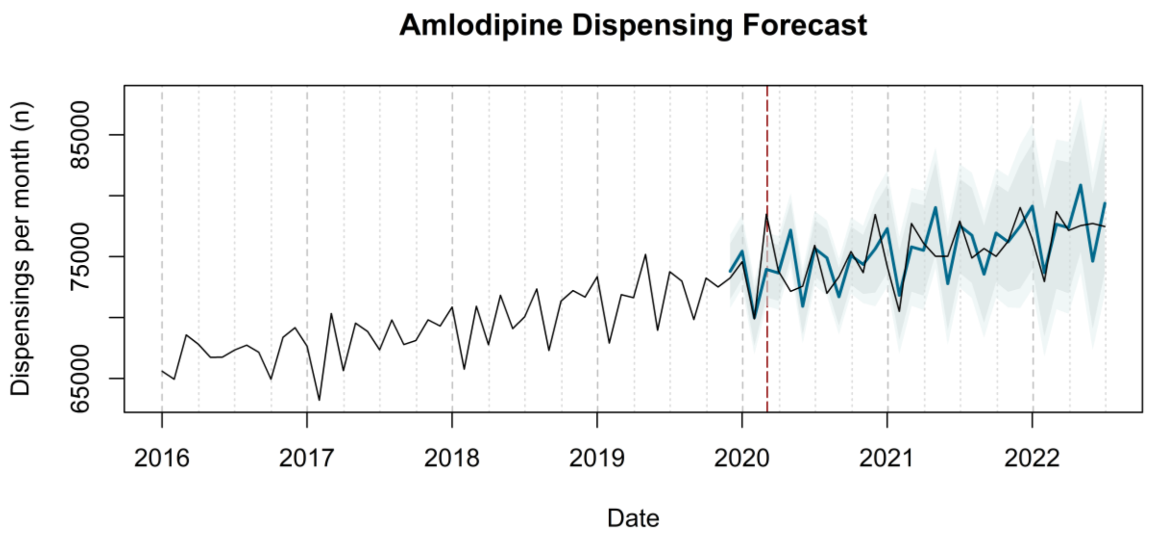

Amlodipine Dispensing Forecast
Dispensings per month (n)
65000
75000
85000
2016
2017
2018
2019
2020
2021
2022
Date


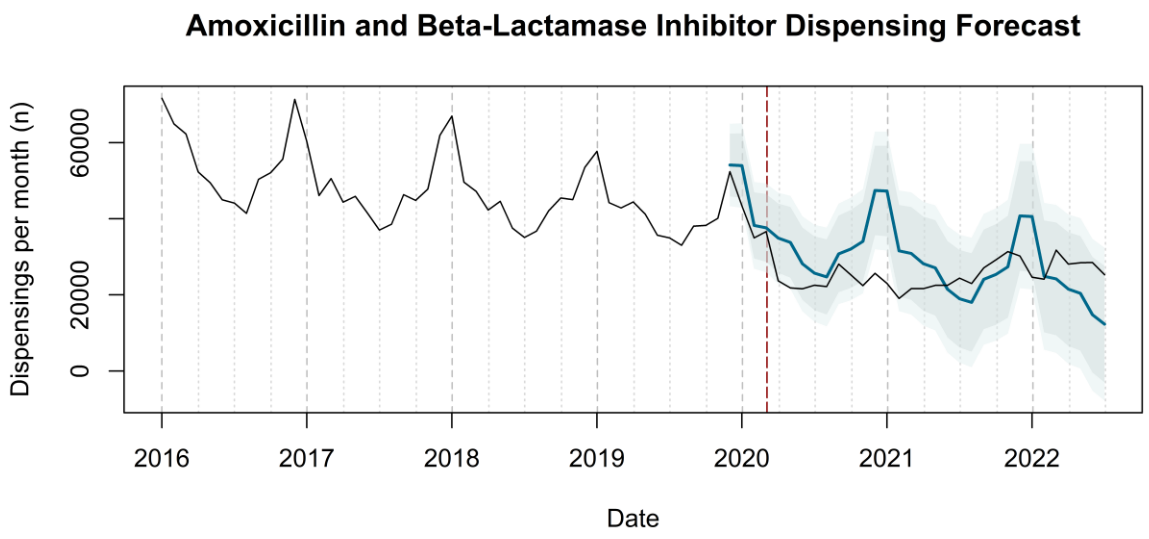

Amoxicillin and Beta-Lactamase Inhibitor Dispensing Forecast
Dispensings per month (n)
0
20000
60000
2016
2017
2018
2019
2020
2021
2022
Date

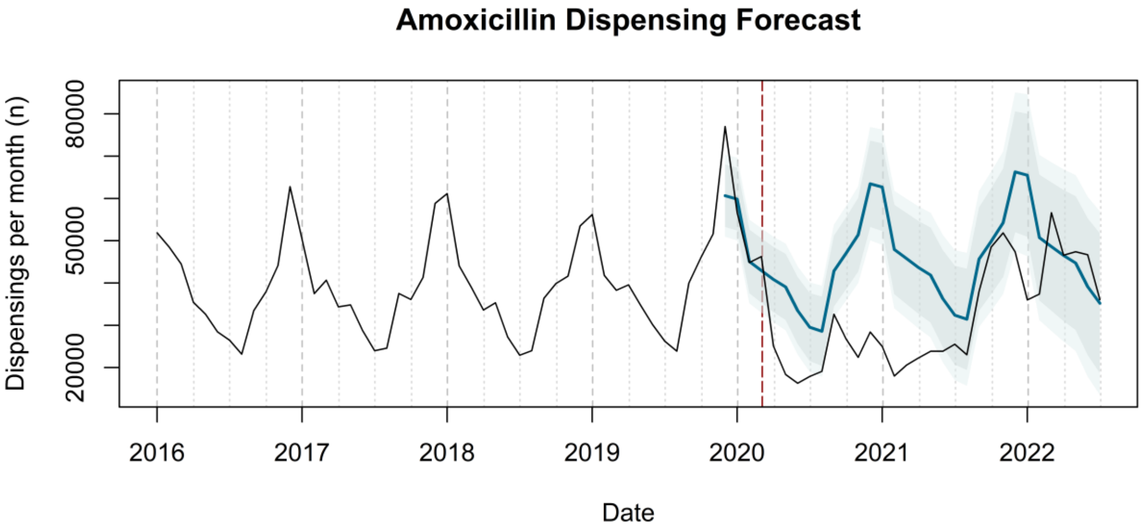
Amoxicillin Dispensing Forecast
Dispensings per month (n)
20000
50000
80000
2016
2017
2018
2019
2020
2021
2022
Date

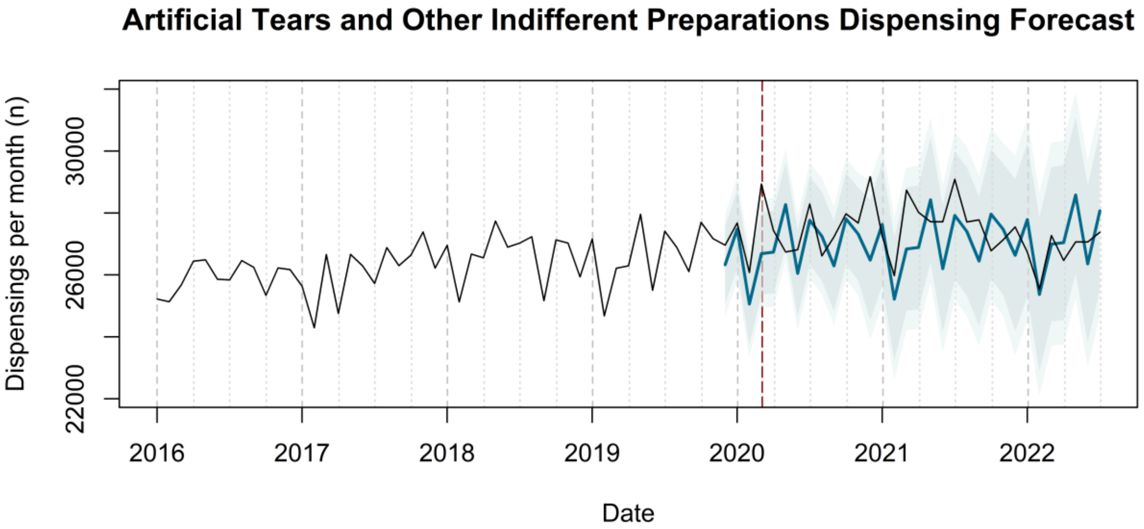
Artificial Tears and Other Indifferent Preparations Dispensing Forecast
Dispensings per month (n)
22000
26000
30000
2016
2017
2018
2019
2020
2021
2022
Date

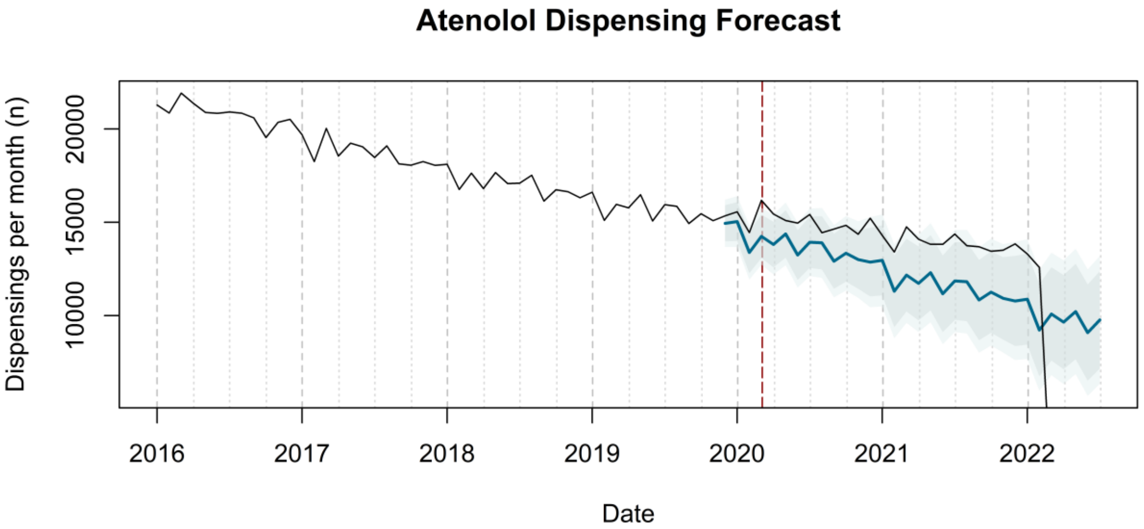
Atenolol Dispensing Forecast
Dispensings per month (n)
10000
15000
20000
2016
2017
2018
2019
2020
2021
2022
Date

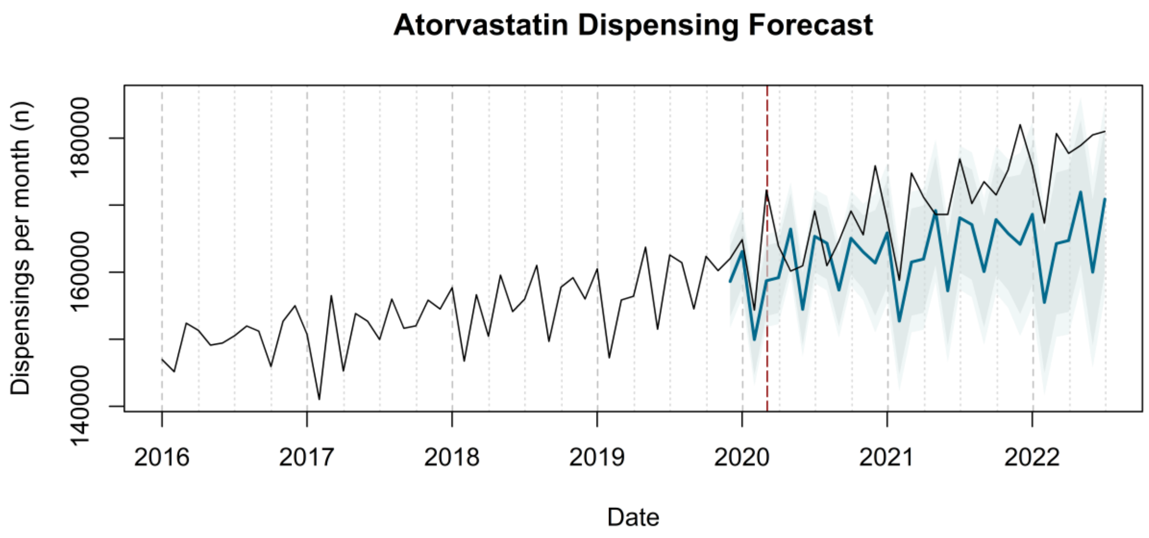

Atorvastatin Dispensing Forecast
Dispensings per month (n)
140000
160000
180000
2016
2017
2018
2019
2020
2021
2022
Date


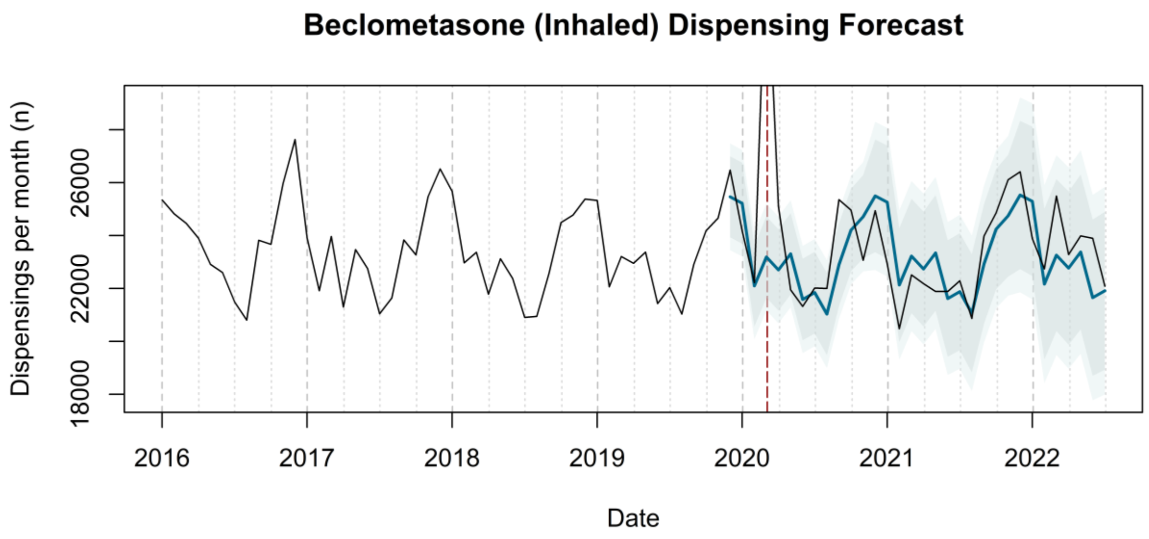

Beclometasone (Inhaled) Dispensing Forecast
Dispensings per month (n)
18000
22000
26000
2016
2017
2018
2019
2020
2021
2022
Date


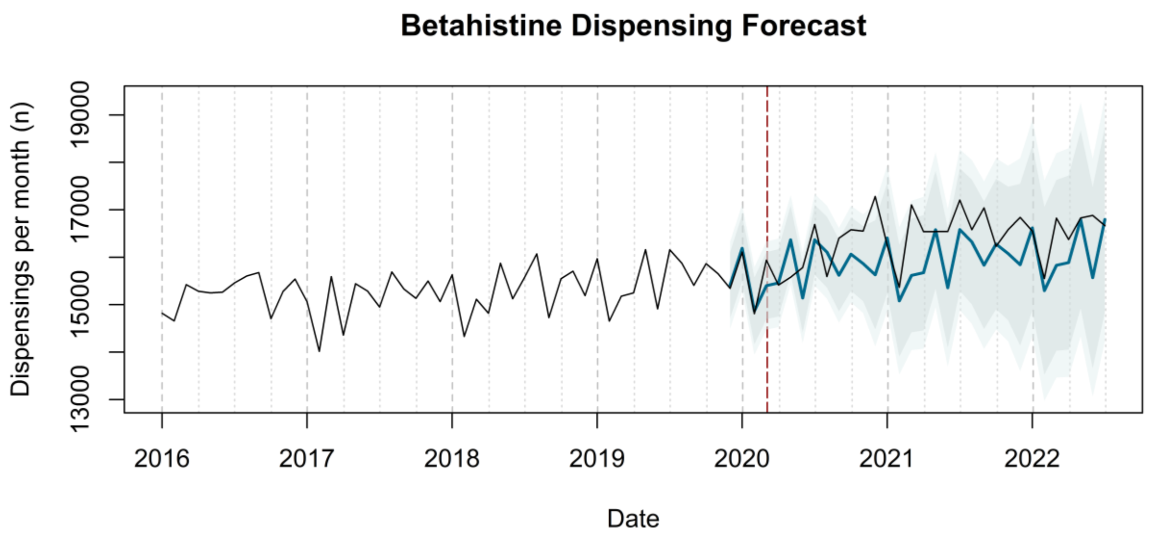

Betahistine Dispensing Forecast
Dispensings per month (n)
13000
15000
17000
19000
2016
2017
2018
2019
2020
2021
2022
Date

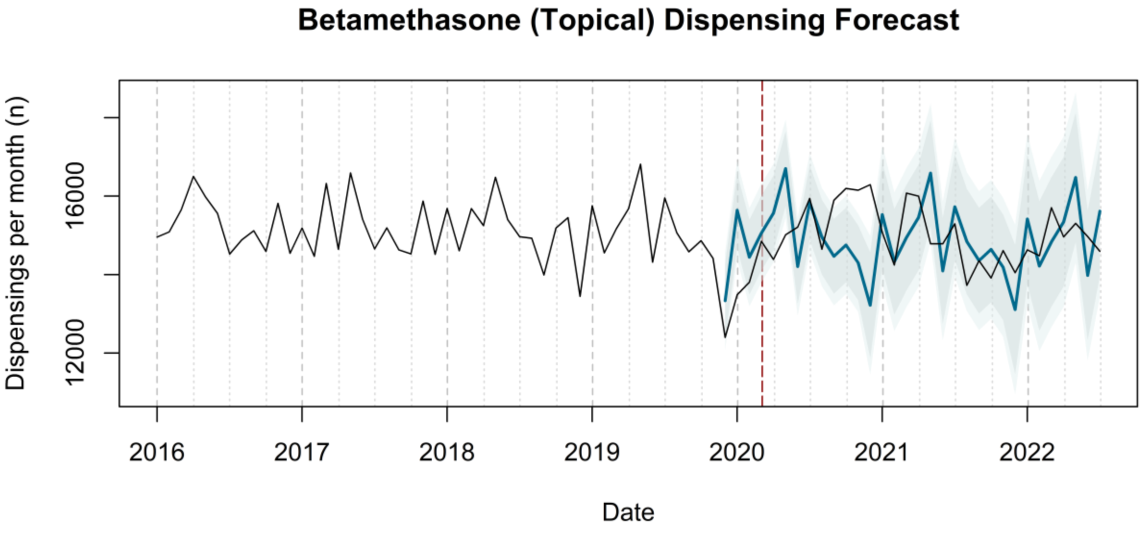
Betamethasone (Topical) Dispensing Forecast
Dispensings per month (n)
12000
16000
2016
2017
2018
2019
2020
2021
2022
Date

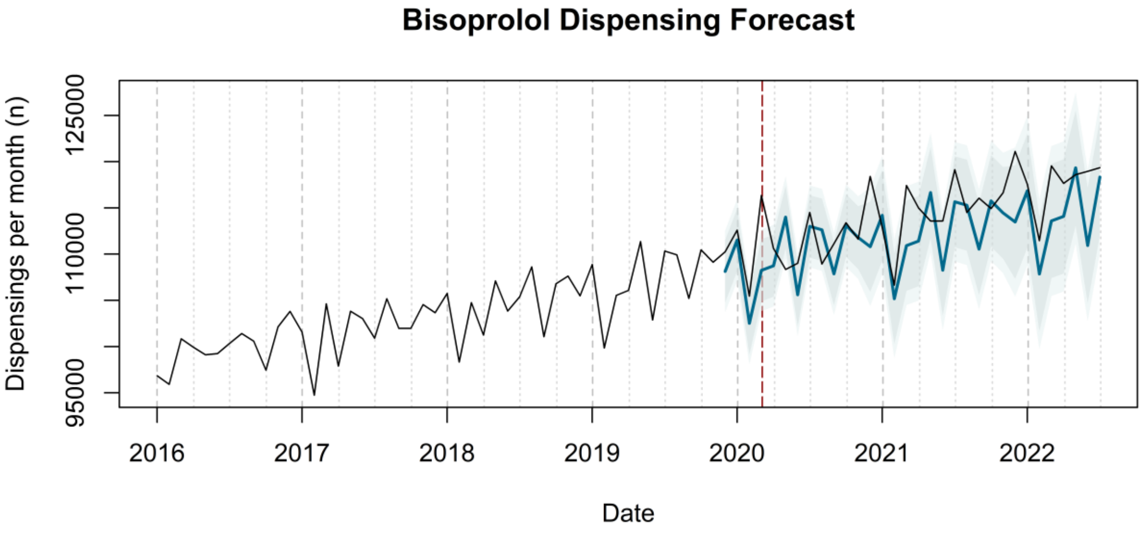
Bisoprolol Dispensing Forecast
Dispensings per month (n)
95000
110000
125000
2016
2017
2018
2019
2020
2021
2022
Date

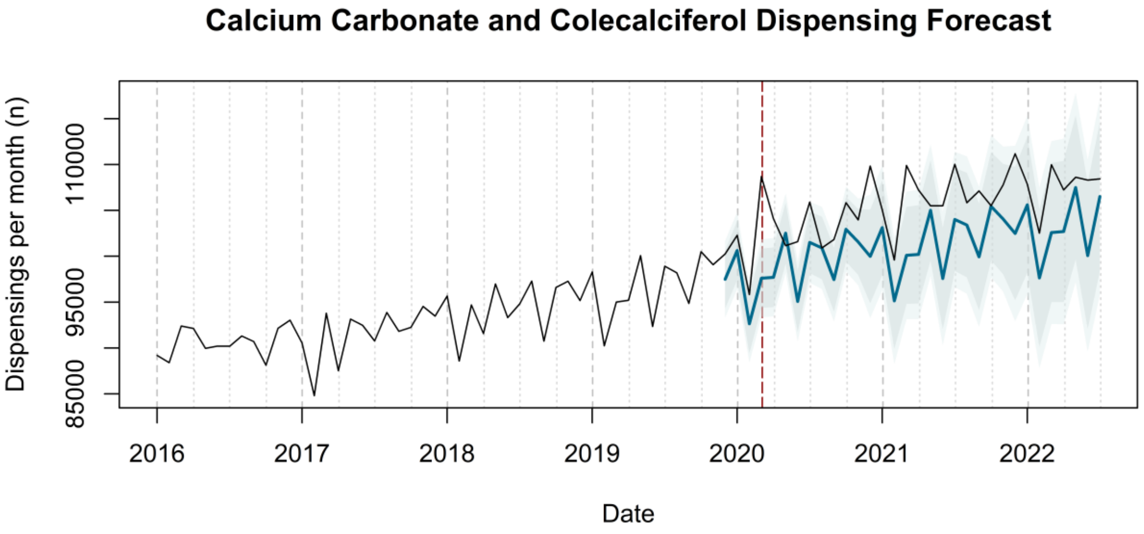
Calcium Carbonate and Colecalciferol Dispensing Forecast
Dispensings per month (n)
85000
95000
110000
2016
2017
2018
2019
2020
2021
2022
Date

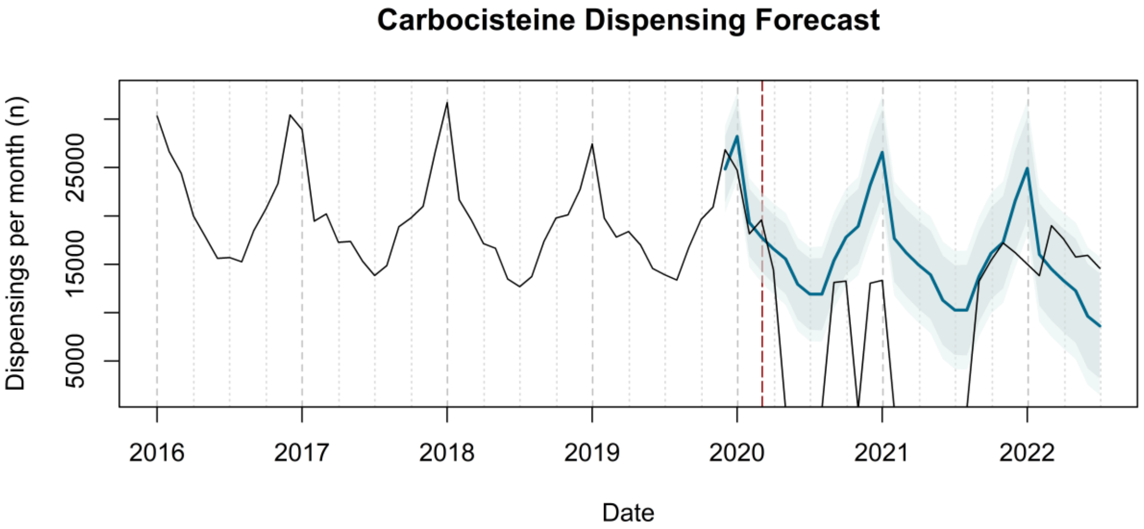
Carbocisteine Dispensing Forecast
Dispensings per month (n)
5000
15000
25000
2016
2017
2018
2019
2020
2021
2022
Date

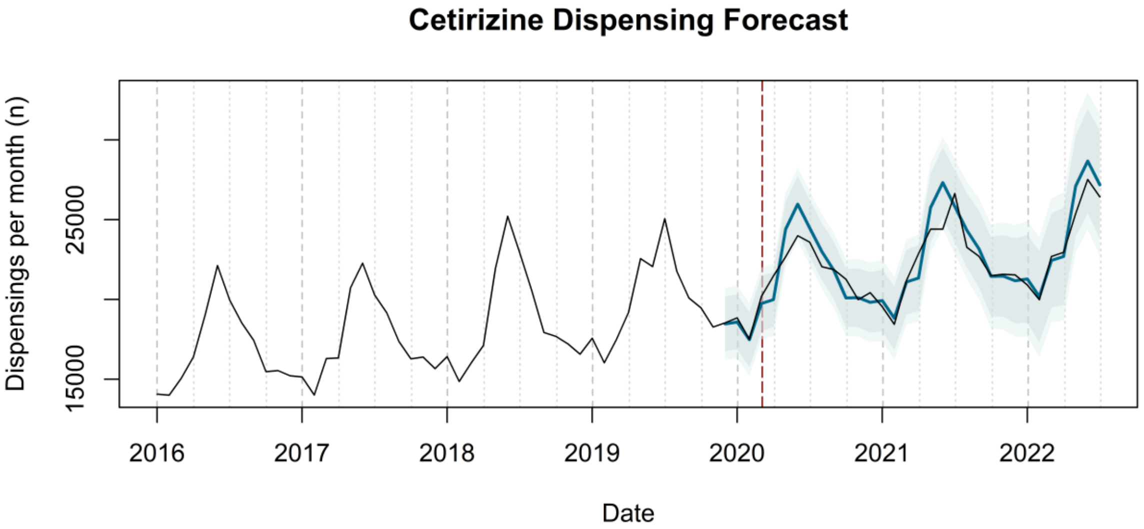
Cetirizine Dispensing Forecast
Dispensings per month (n)
15000
25000
2016
2017
2018
2019
2020
2021
2022
Date

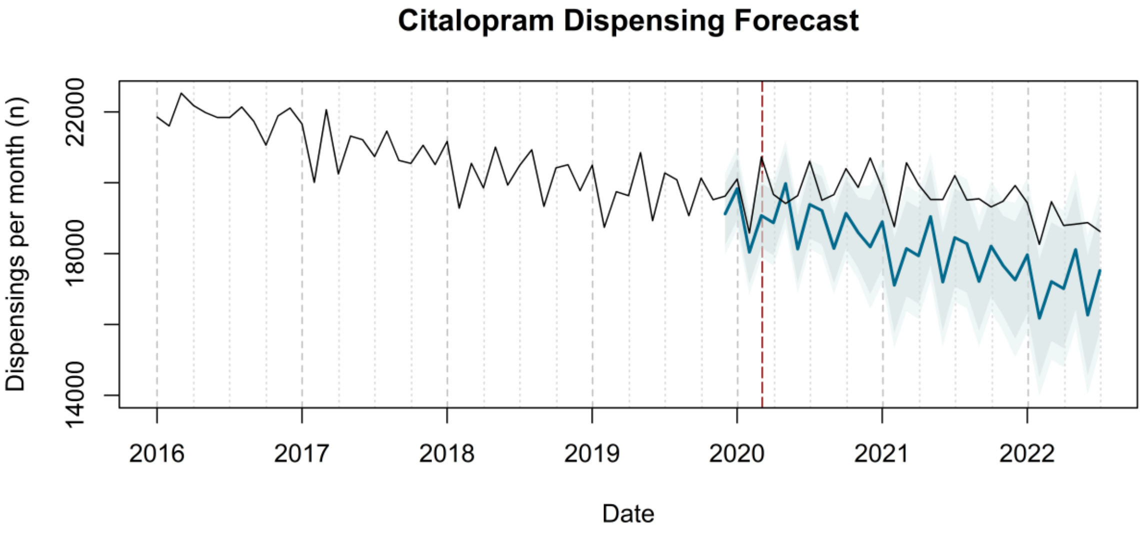
Citalopram Dispensing Forecast
Dispensings per month (n)
14000
18000
22000
2016
2017
2018
2019
2020
2021
2022
Date

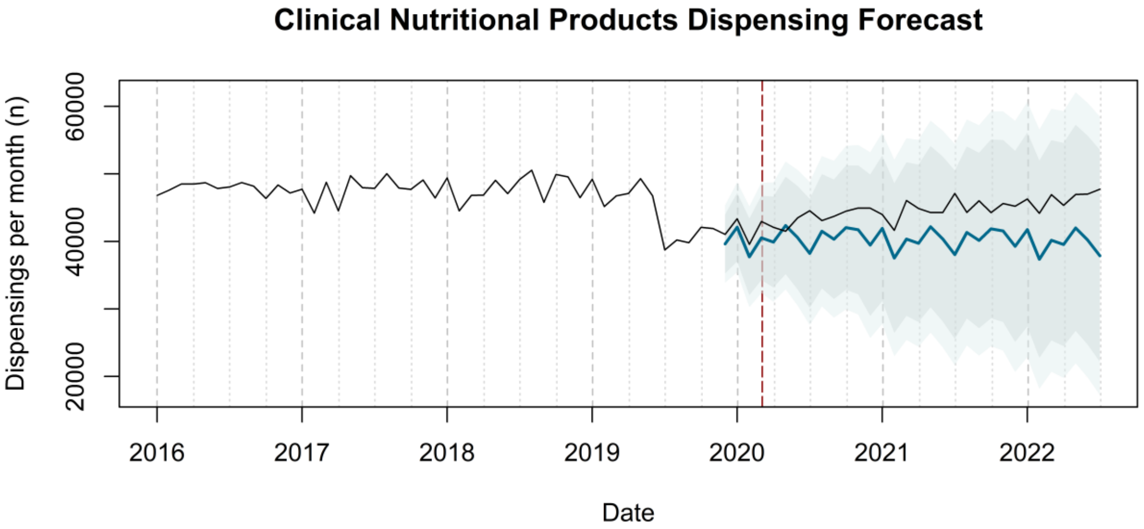
Clinical Nutritional Products Dispensing Forecast
Dispensings per month (n)
20000
40000
60000
2016
2017
2018
2019
2020
2021
2022
Date

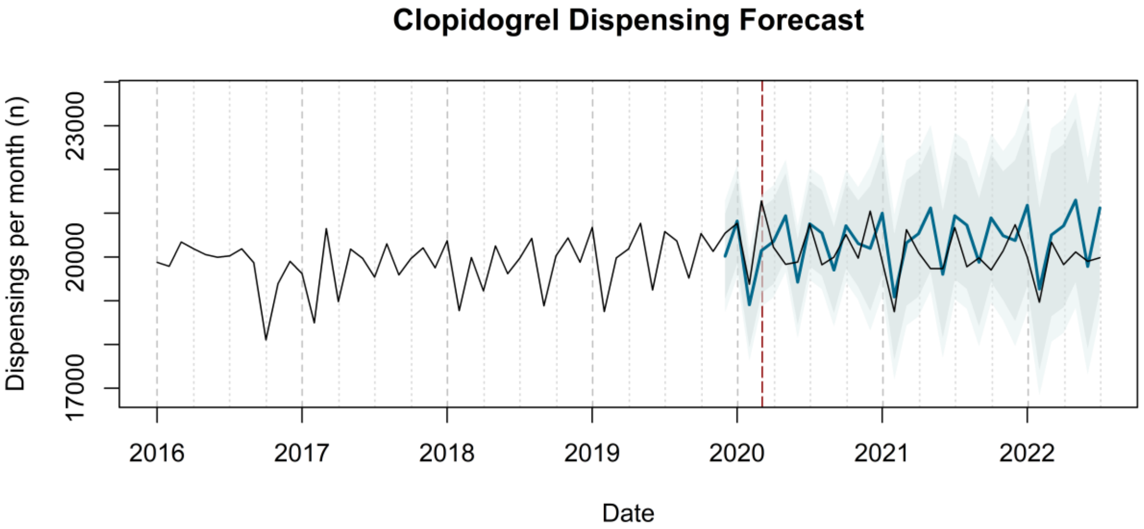
Clopidogrel Dispensing Forecast
Dispensings per month (n)
17000
20000
23000
2016
2017
2018
2019
2020
2021
2022
Date

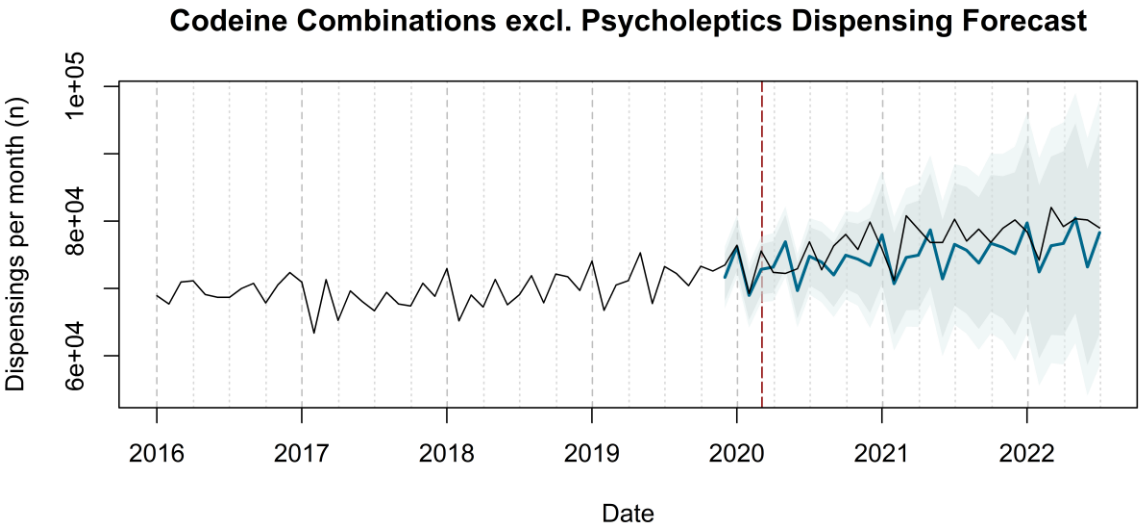
Codeine Combinations excl. Psycholeptics Dispensing Forecast
Dispensings per month (n)
6e+04
8e+04
1e+05
2016
2017
2018
2019
2020
2021
2022
Date

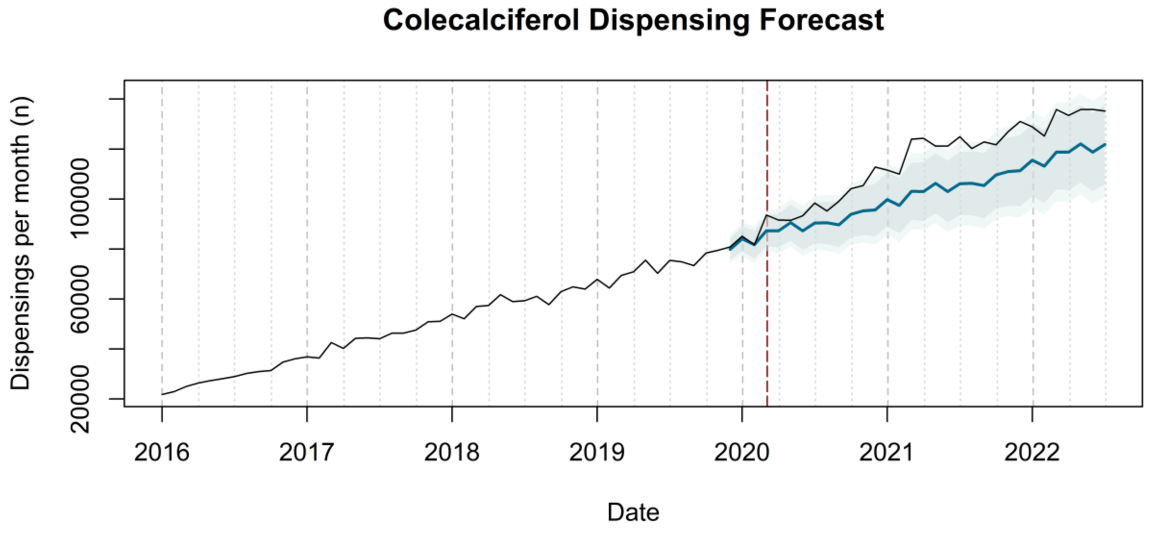
Colecalciferol Dispensing Forecast
Dispensings per month (n)
20000
60000
100000
2016
2017
2018
2019
2020
2021
2022
Date

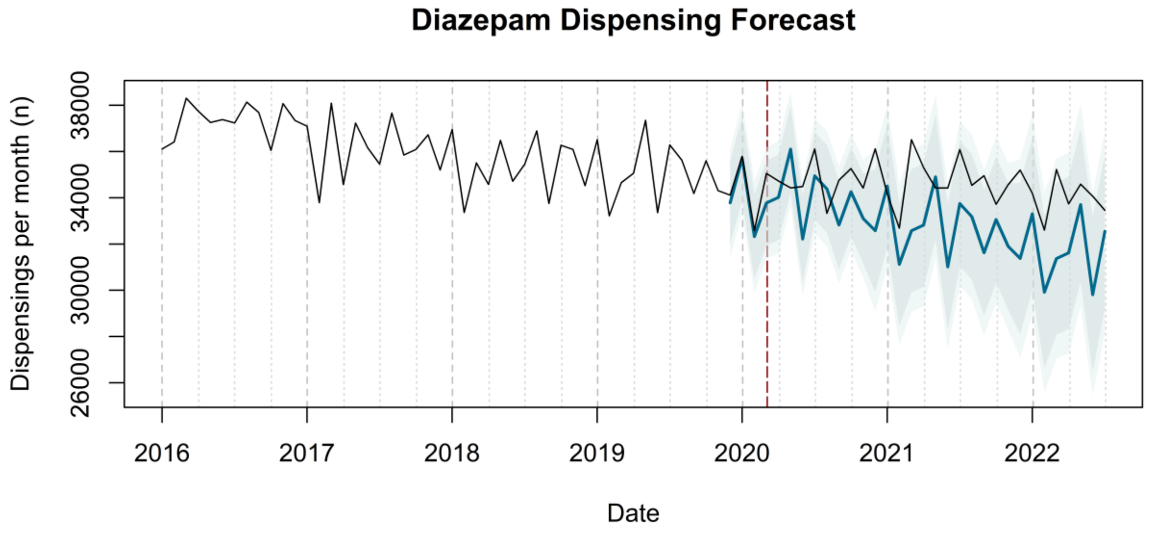
Diazepam Dispensing Forecast
Dispensings per month (n)
26000
30000
34000
38000
2016
2017
2018
2019
2020
2021
2022
Date

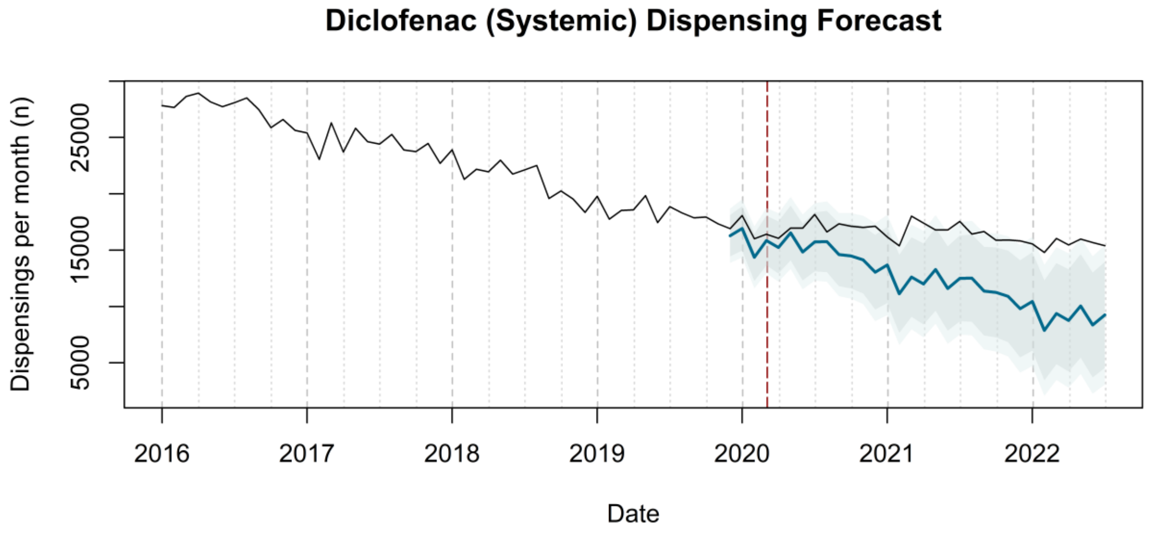
Diclofenac (Systemic) Dispensing Forecast
Dispensings per month (n)
5000
15000
25000
2016
2017
2018
2019
2020
2021
2022
Date

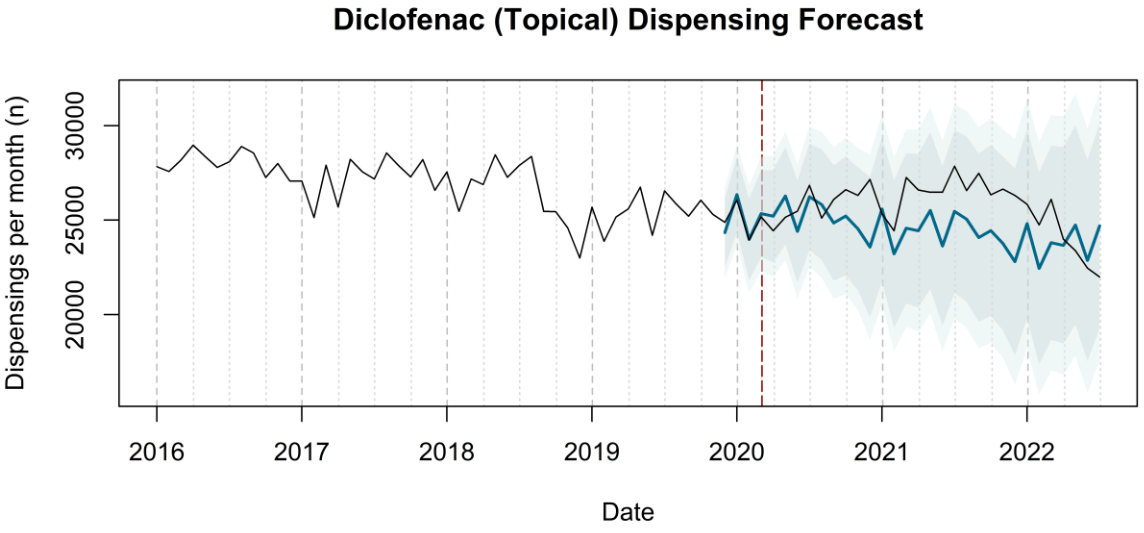
Diclofenac (Topical) Dispensing Forecast
Dispensings per month (n)
20000
25000
30000
2016
2017
2018
2019
2020
2021
2022
Date

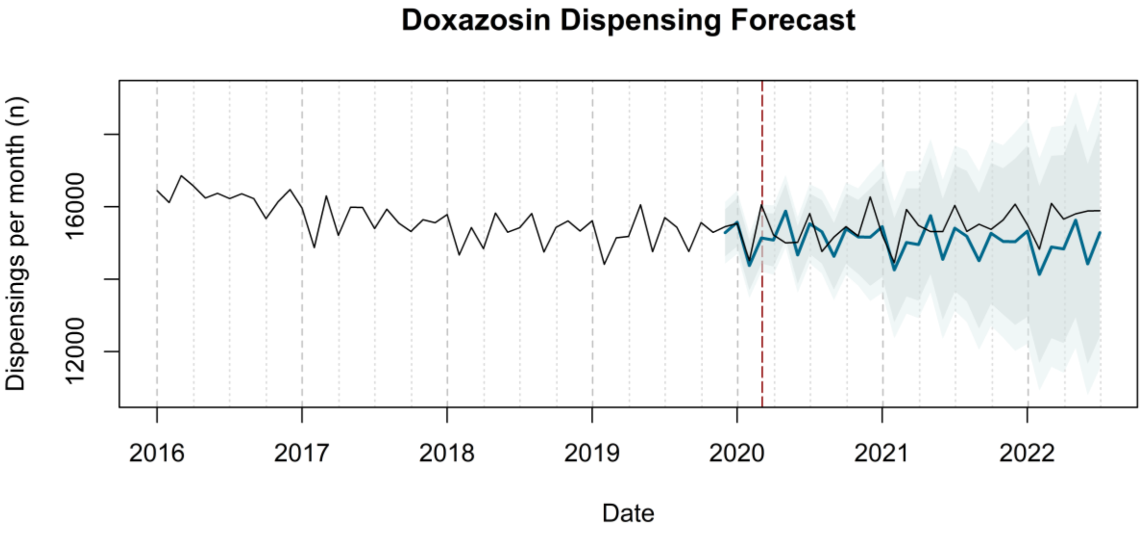
Doxazosin Dispensing Forecast
Dispensings per month (n)
12000
16000
2016
2017
2018
2019
2020
2021
2022
Date

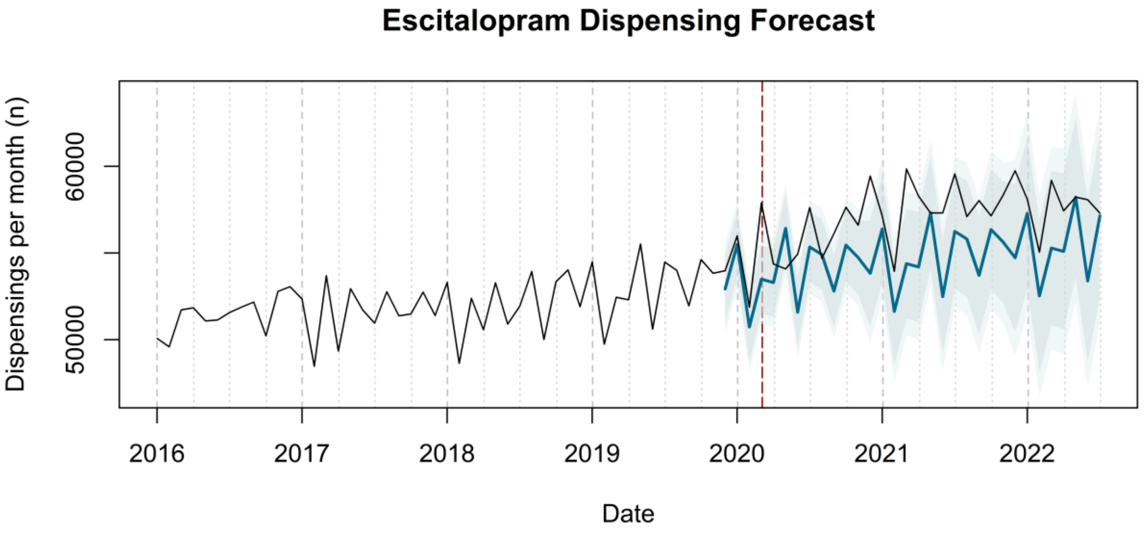
Escitalopram Dispensing Forecast
Dispensings per month (n)
50000
60000
2016
2017
2018
2019
2020
2021
2022
Date

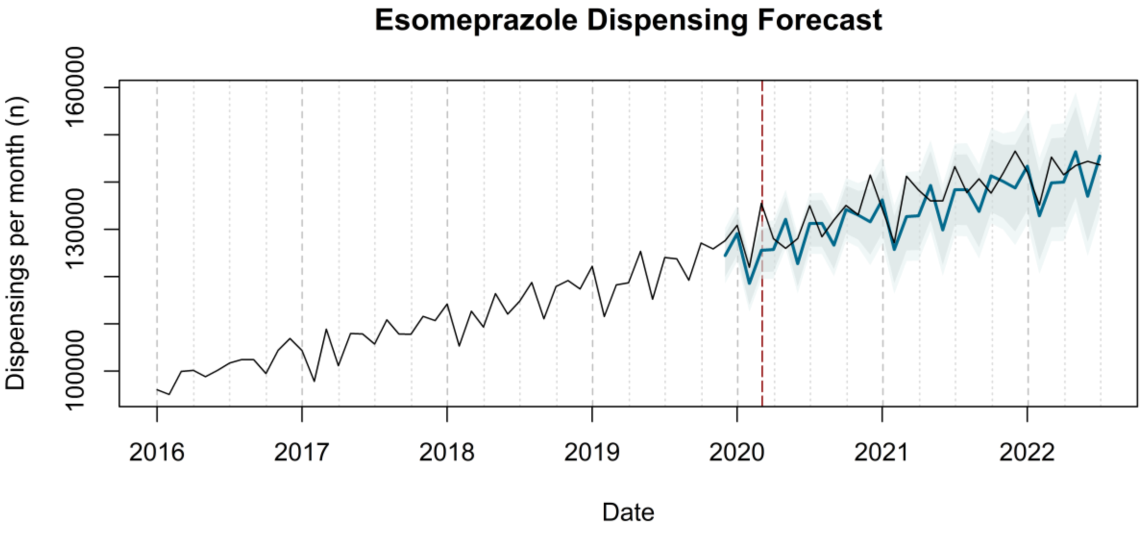
Esomeprazole Dispensing Forecast
Dispensings per month (n)
100000
130000
160000
2016
2017
2018
2019
2020
2021
2022
Date

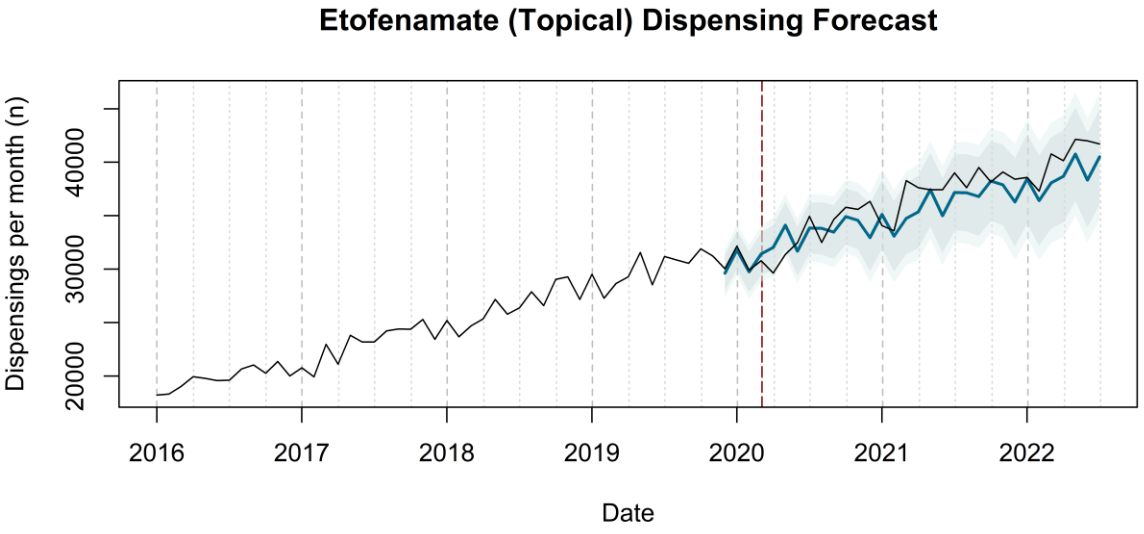
Etofenamate (Topical) Dispensing Forecast
Dispensings per month (n)
20000
30000
40000
2016
2017
2018
2019
2020
2021
2022
Date

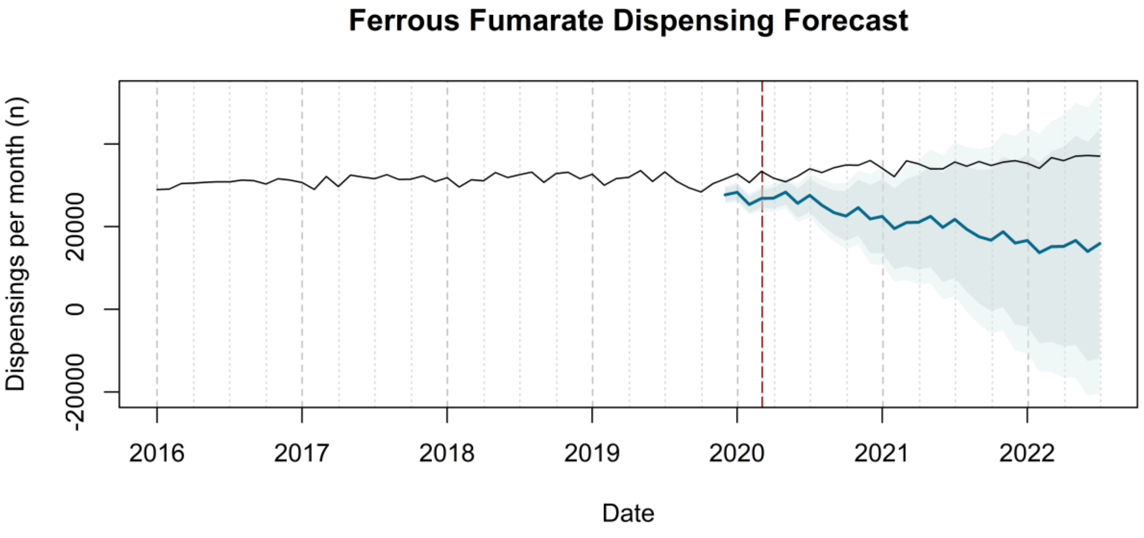
Ferrous Fumarate Dispensing Forecast
Dispensings per month (n)
-20000
0
20000
2016
2017
2018
2019
2020
2021
2022
Date

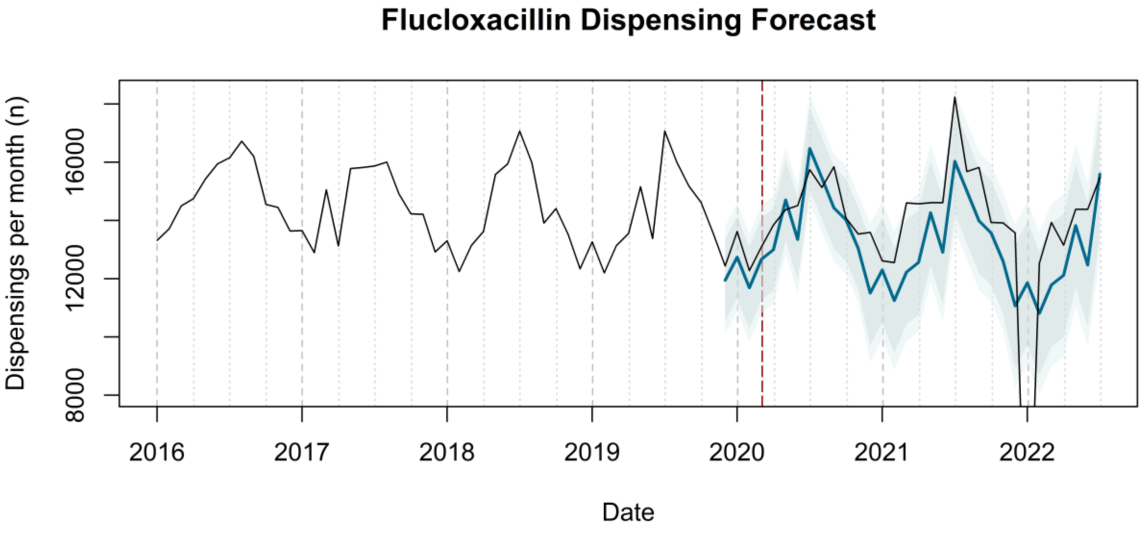
Flucloxacillin Dispensing Forecast
Dispensings per month (n)
8000
12000
16000
2016
2017
2018
2019
2020
2021
2022
Date

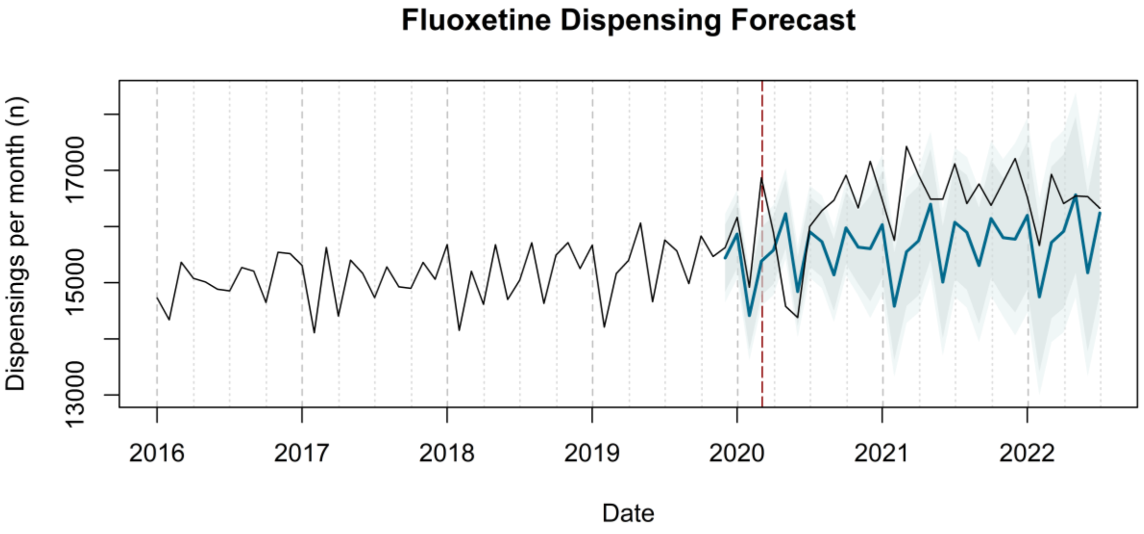
Fluoxetine Dispensing Forecast
Dispensings per month (n)
13000
15000
17000
2016
2017
2018
2019
2020
2021
2022
Date

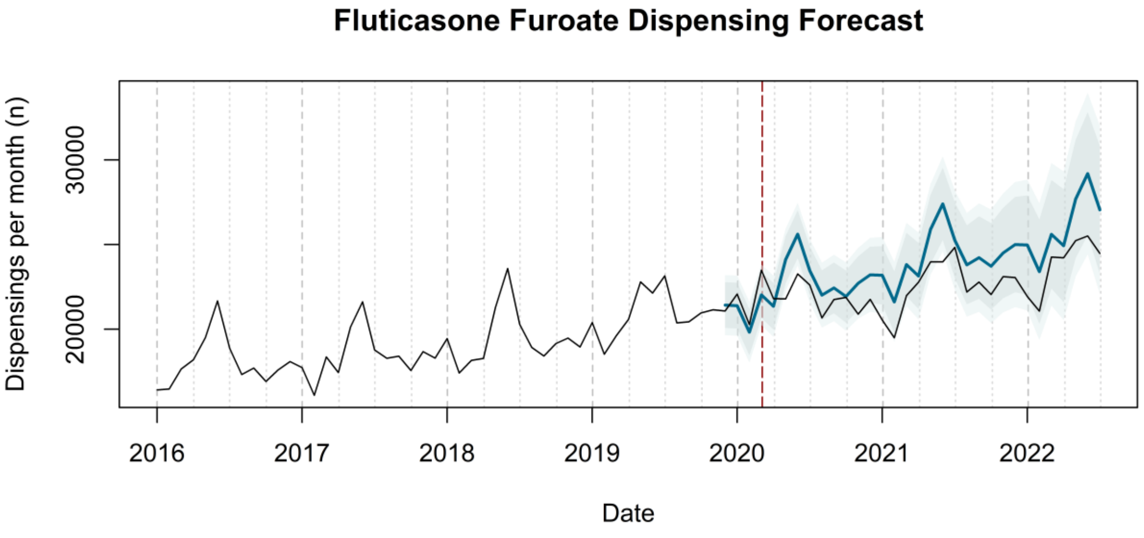
Fluticasone Furoate Dispensing Forecast
Dispensings per month (n)
20000
30000
2016
2017
2018
2019
2020
2021
2022
Date

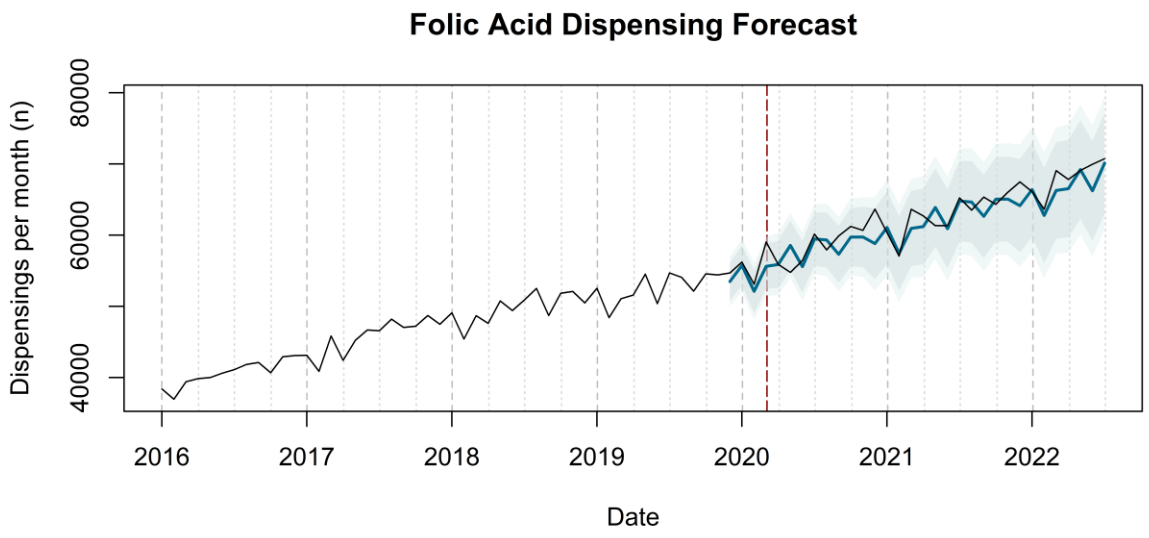
Folic Acid Dispensing Forecast
Dispensings per month (n)
40000
60000
80000
2016
2017
2018
2019
2020
2021
2022
Date

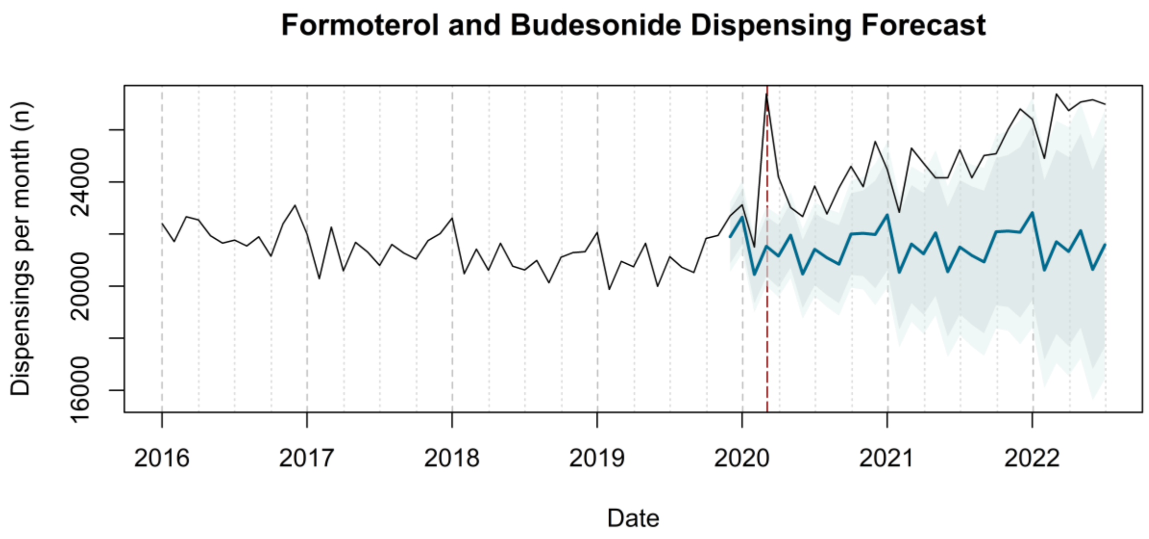
Formoterol and Budesonide Dispensing Forecast
Dispensings per month (n)
16000
20000
24000
2016
2017
2018
2019
2020
2021
2022
Date

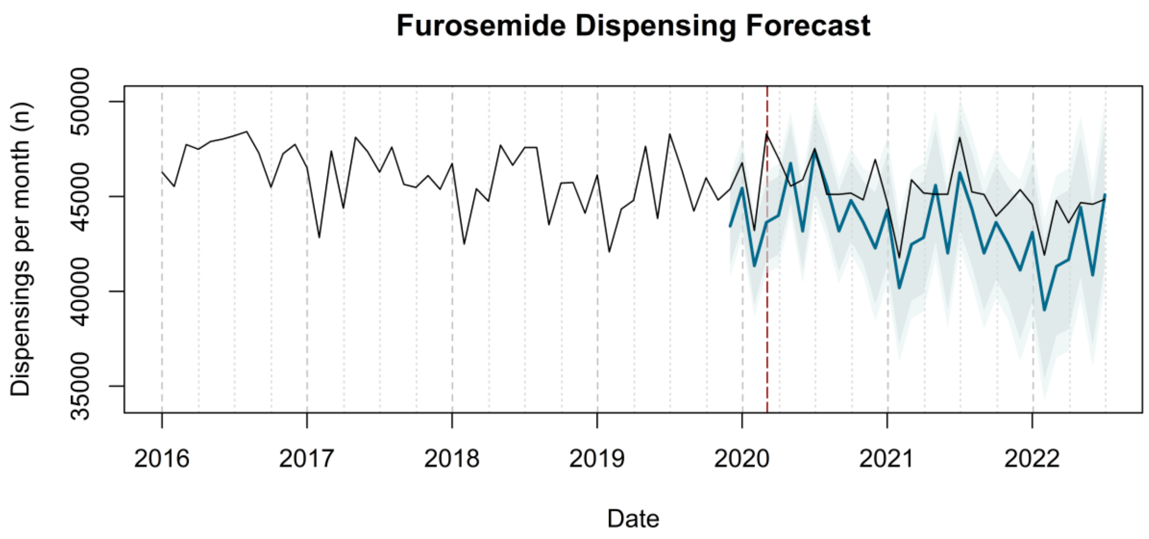
Furosemide Dispensing Forecast
Dispensings per month (n)
35000
40000
45000
50000
2016
2017
2018
2019
2020
2021
2022
Date

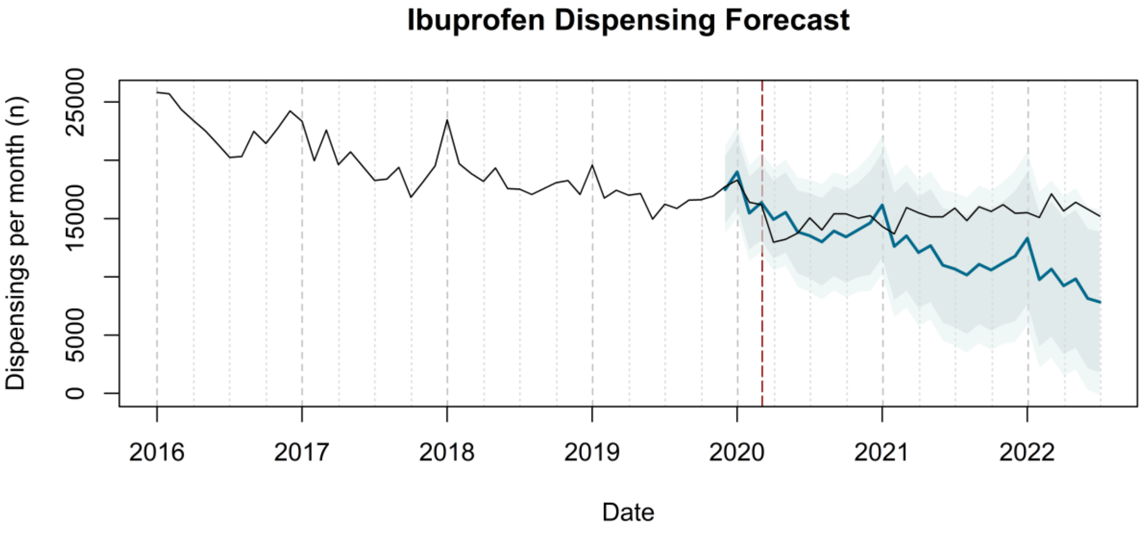
Ibuprofen Dispensing Forecast
Dispensings per month (n)
0
5000
15000
25000
2016
2017
2018
2019
2020
2021
2022
Date

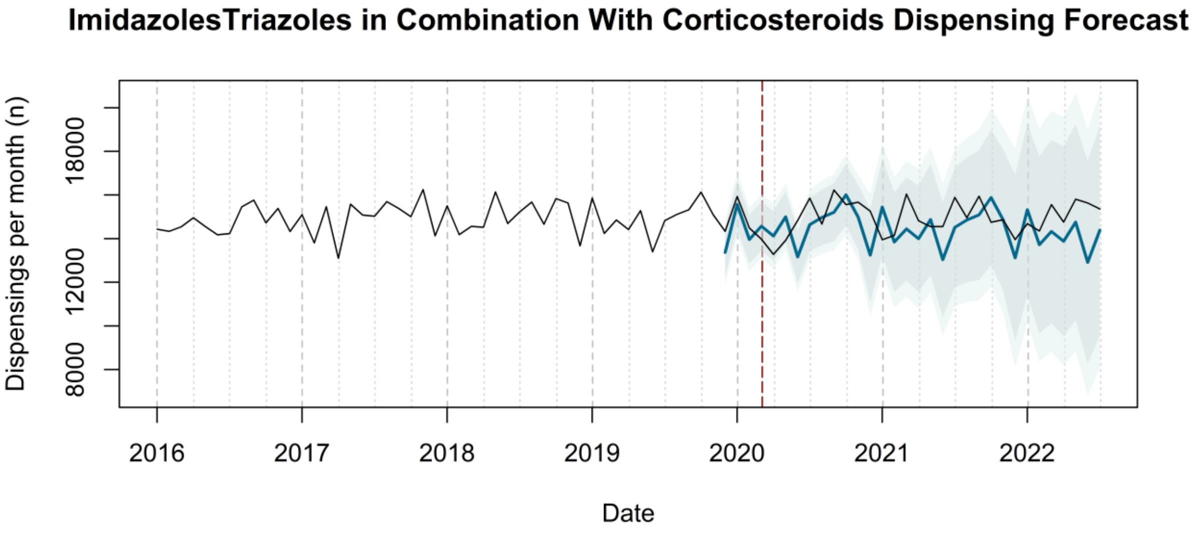
ImidazolesTriazoles in Combination With Corticosteroids Dispensing Forecast
Dispensings per month (n)
8000
12000
18000
2016
2017
2018
2019
2020
2021
2022
Date

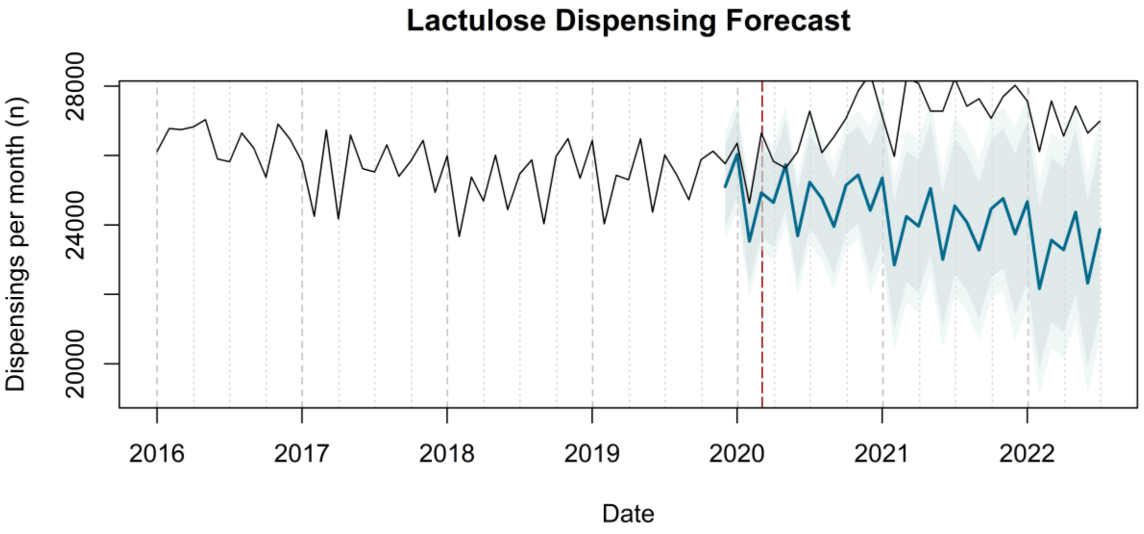
Lactulose Dispensing Forecast
Dispensings per month (n)
20000
24000
28000
2016
2017
2018
2019
2020
2021
2022
Date

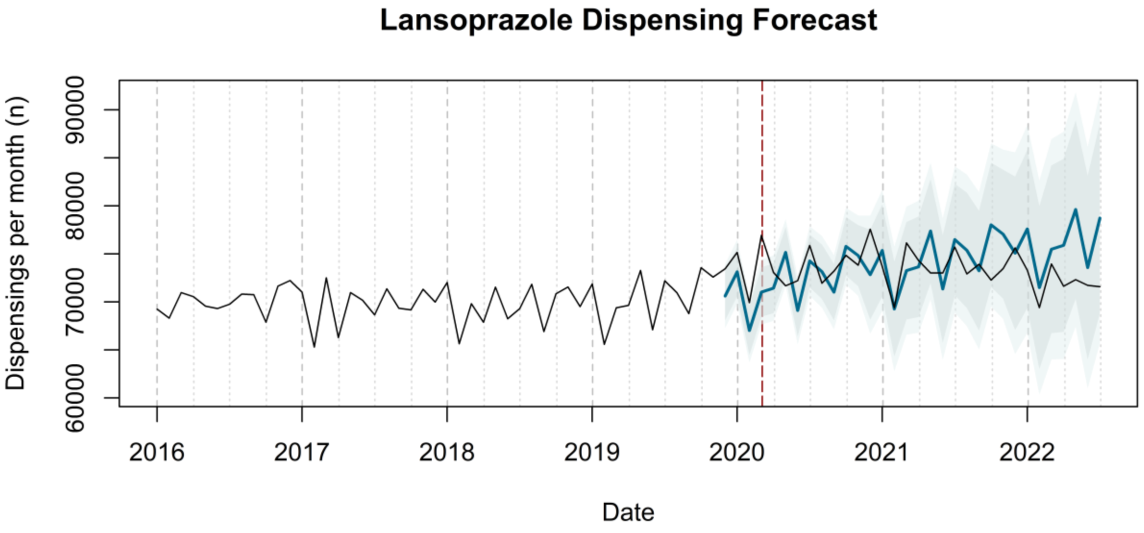

Lansoprazole Dispensing Forecast
Dispensings per month (n)
60000
70000
80000
90000
2016
2017
2018
2019
2020
2021
2022
Date


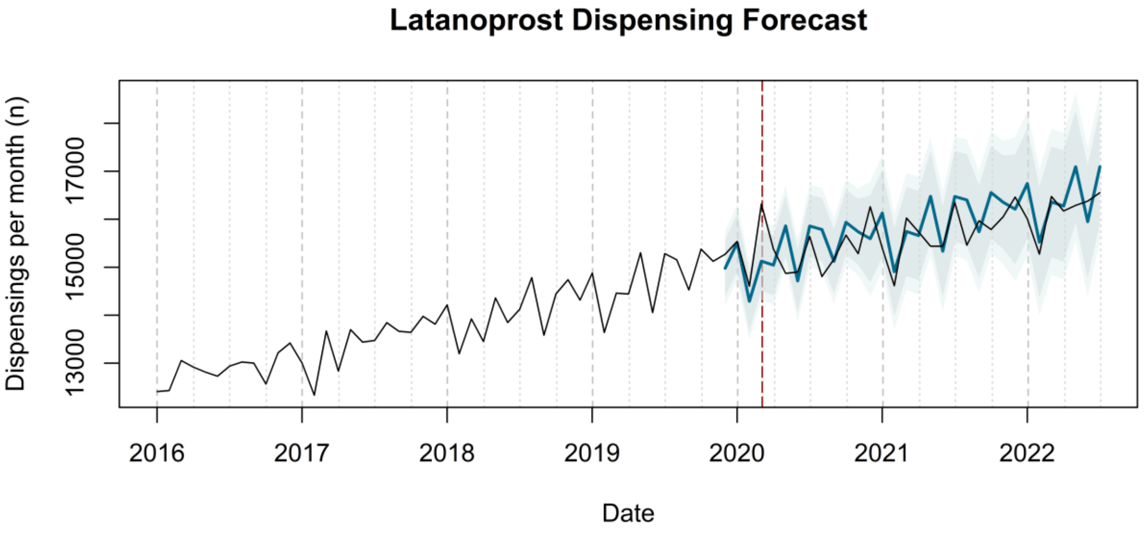

Latanoprost Dispensing Forecast
Dispensings per month (n)
13000
15000
17000
2016
2017
2018
2019
2020
2021
2022
Date


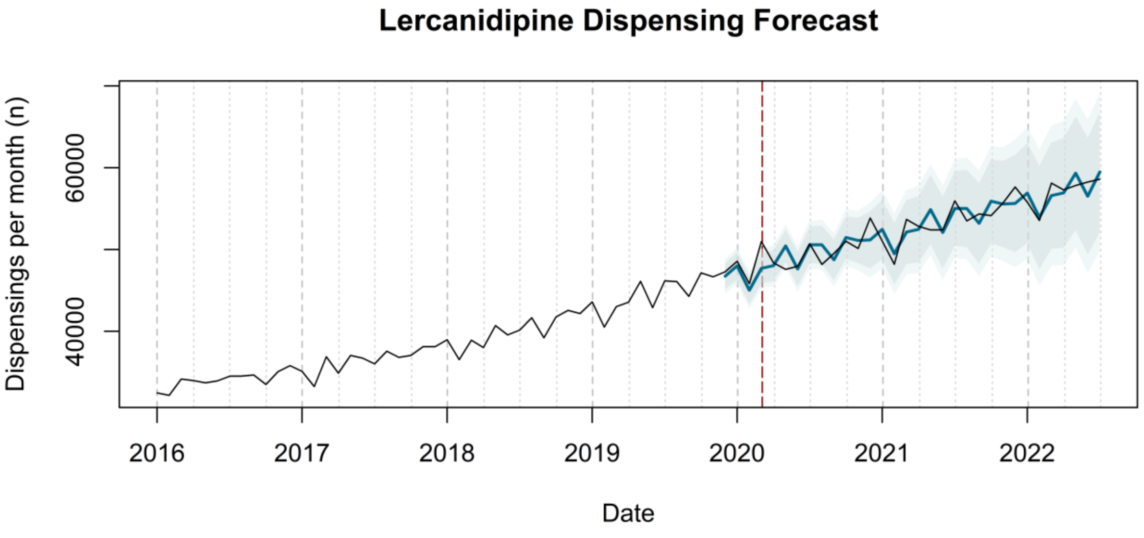

Lercanidipine Dispensing Forecast
Dispensings per month (n)
40000
60000
2016
2017
2018
2019
2020
2021
2022
Date

**Levonorgestrel and Ethinylestradiol Dispensing Forecast**

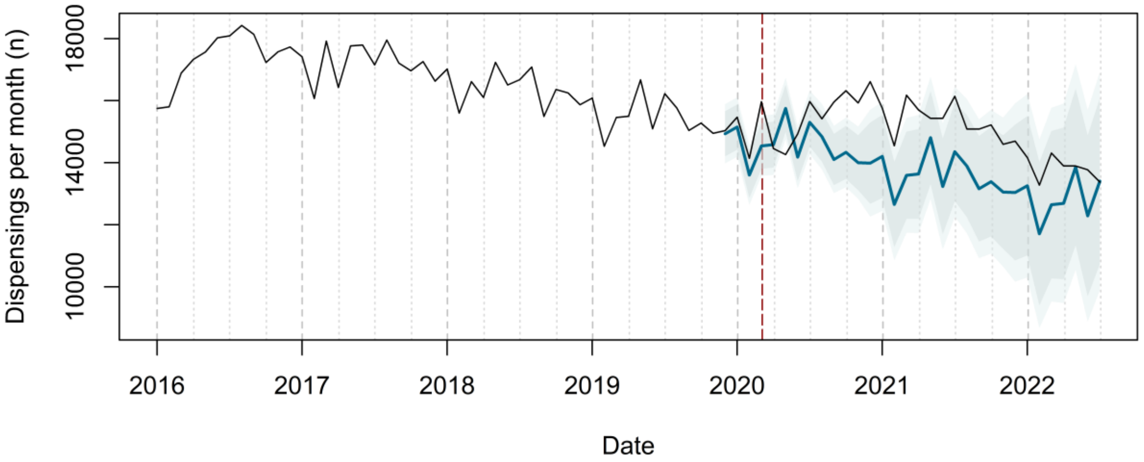


**Levothyroxine Sodium Dispensing Forecast**

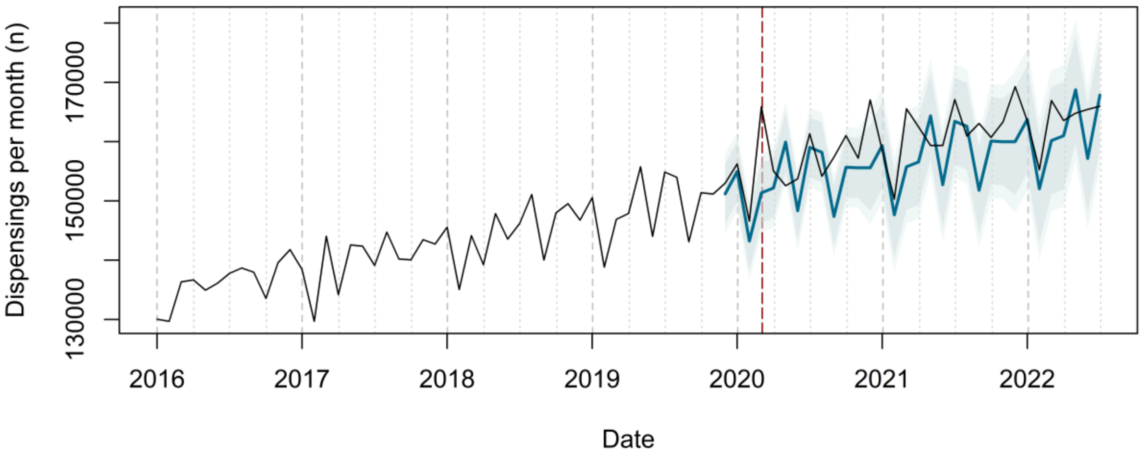

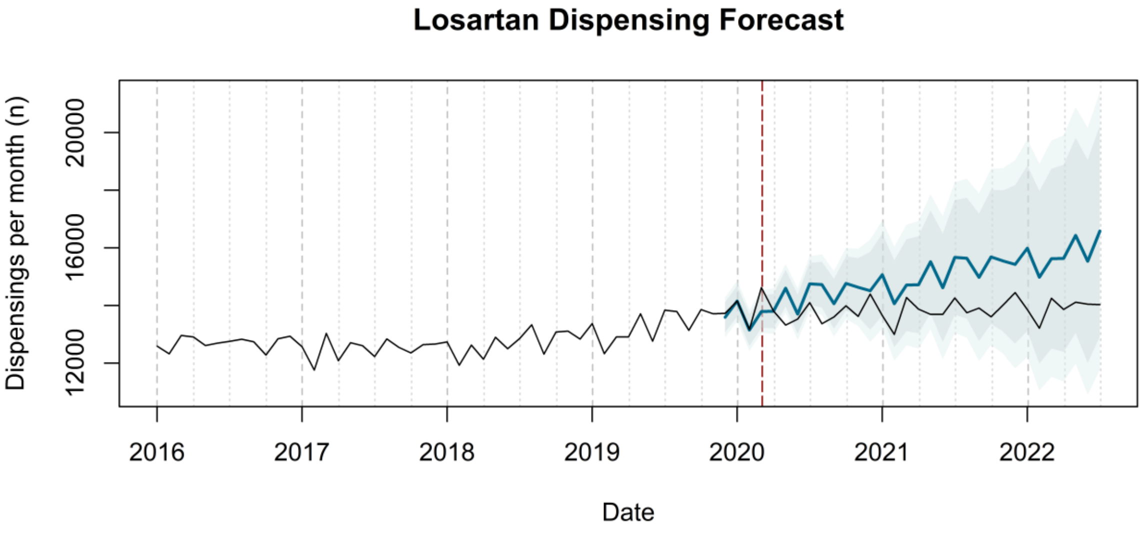
Losartan Dispensing Forecast
Dispensings per month (n)
12000
16000
20000
2016
2017
2018
2019
2020
2021
2022
Date

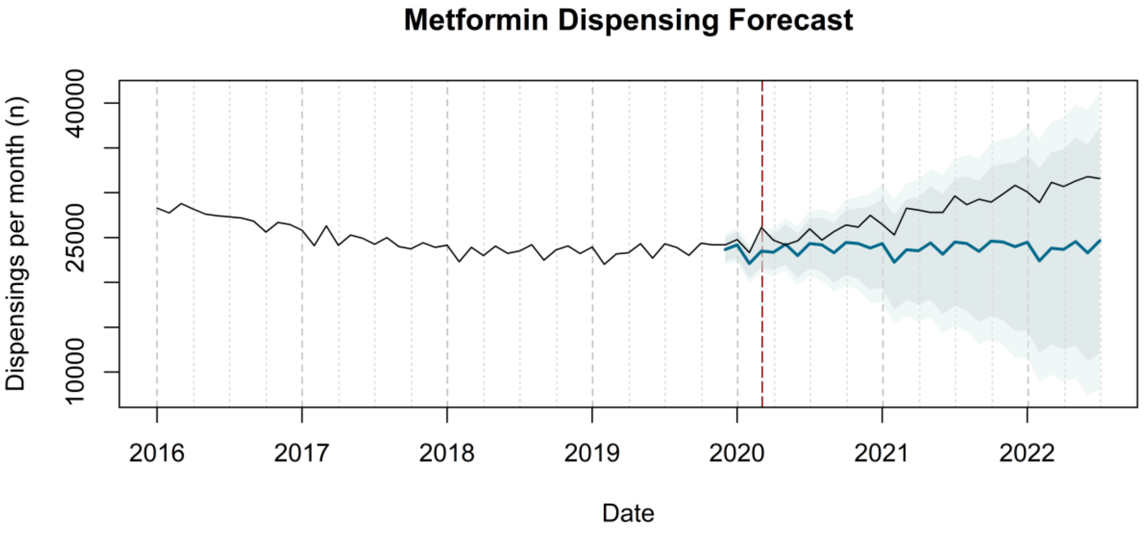
Metformin Dispensing Forecast
Dispensings per month (n)
10000
25000
40000
2016
2017
2018
2019
2020
2021
2022
Date

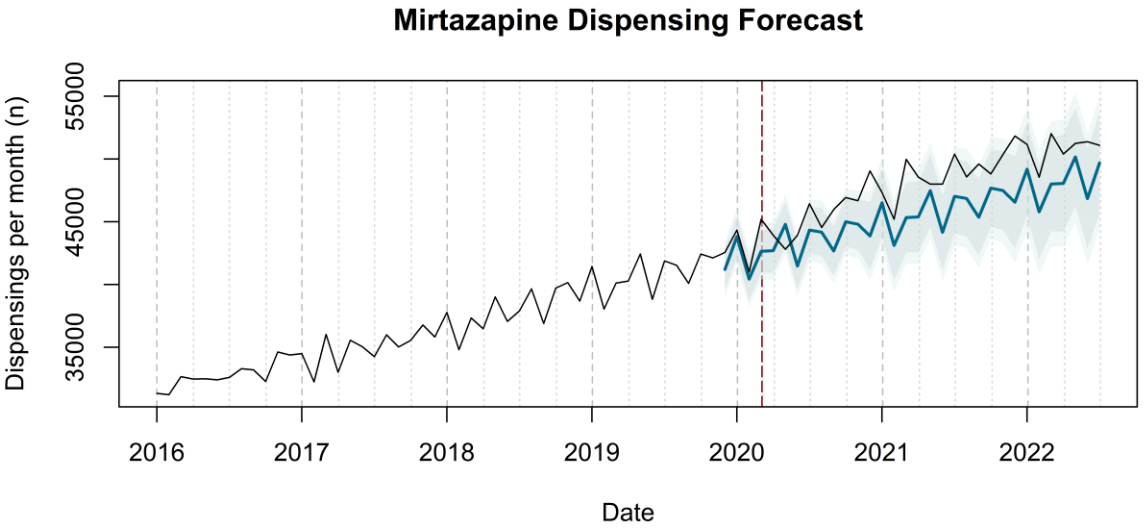
Mirtazapine Dispensing Forecast
Dispensings per month (n)
35000
45000
55000
2016
2017
2018
2019
2020
2021
2022
Date

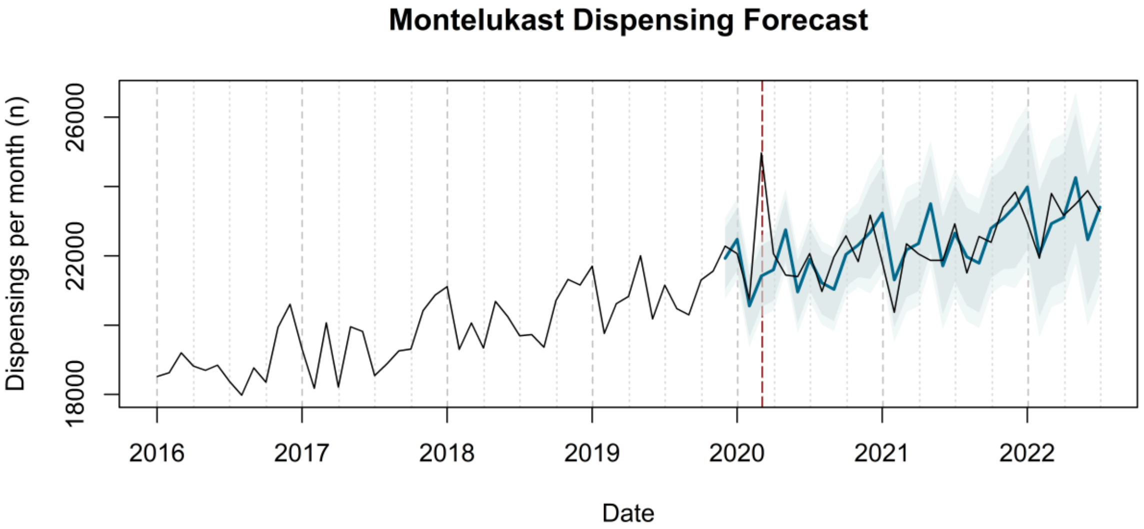
Montelukast Dispensing Forecast
Dispensings per month (n)
18000
22000
26000
2016
2017
2018
2019
2020
2021
2022
Date

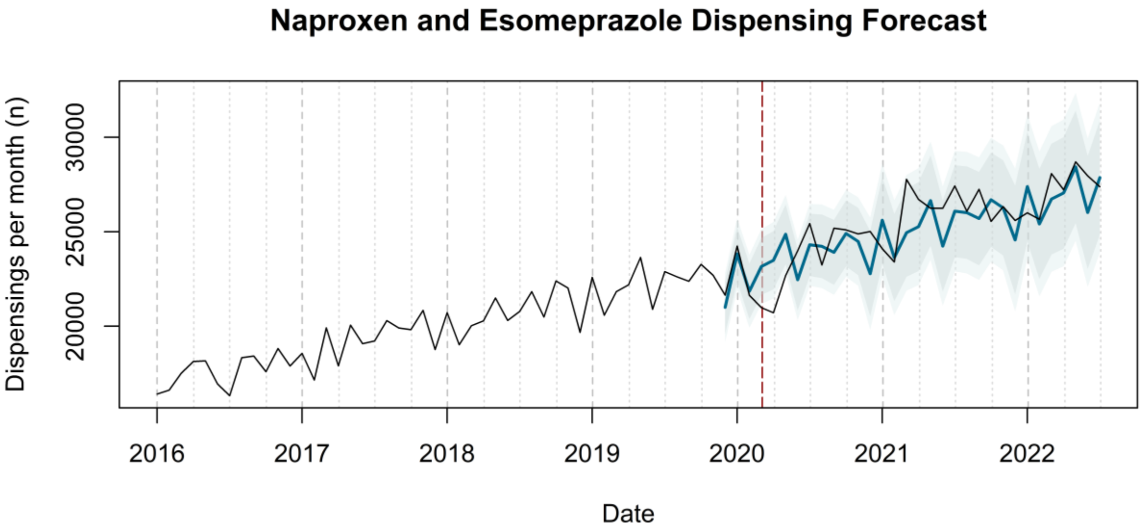
Naproxen and Esomeprazole Dispensing Forecast
Dispensings per month (n)
20000
25000
30000
2016
2017
2018
2019
2020
2021
2022
Date

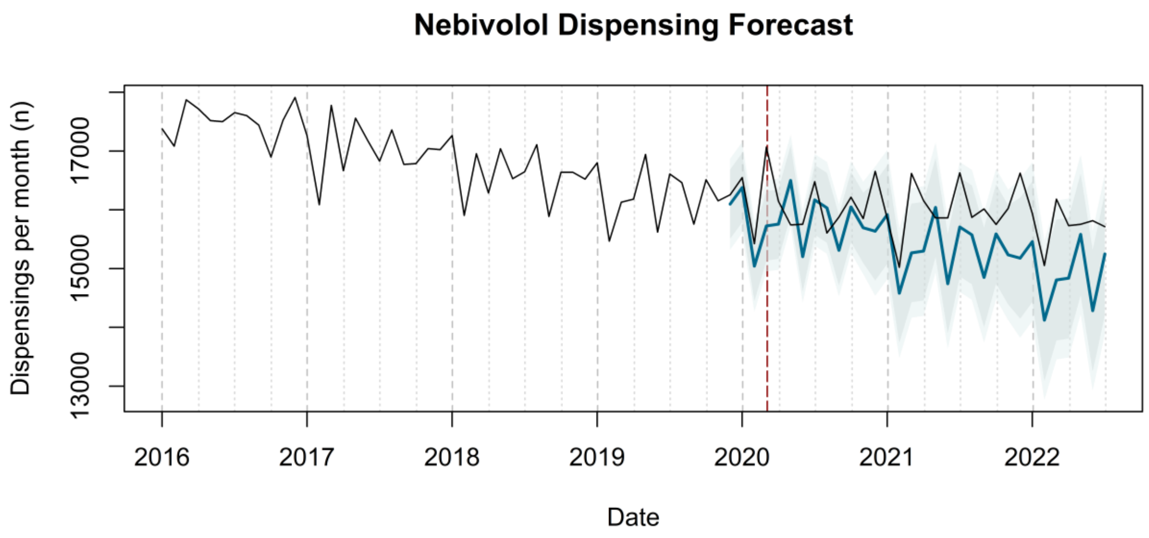
Nebivolol Dispensing Forecast
Dispensings per month (n)
13000
15000
17000
2016
2017
2018
2019
2020
2021
2022
Date

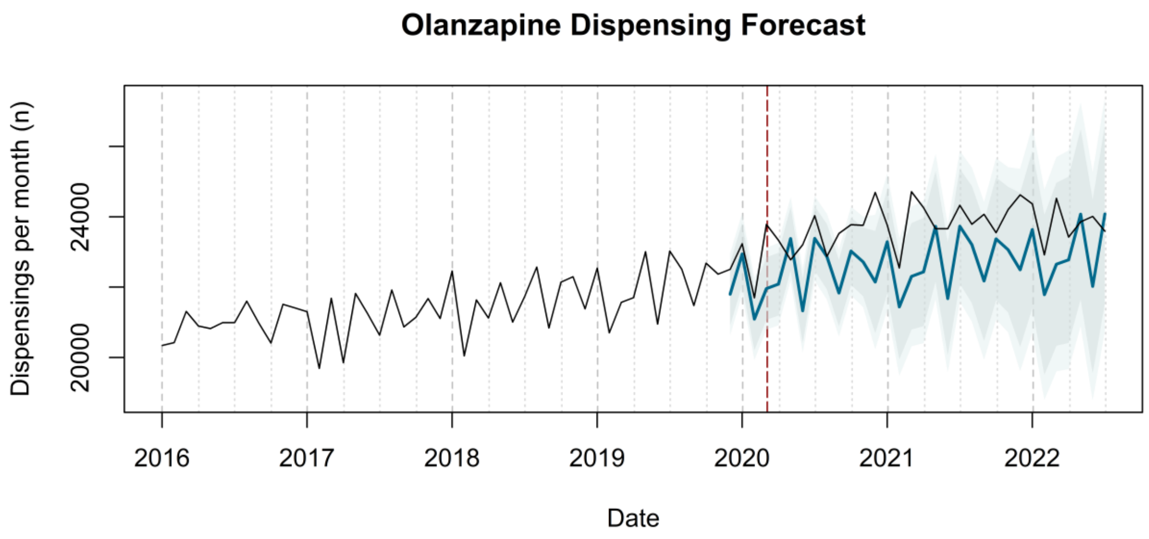
Olanzapine Dispensing Forecast
Dispensings per month (n)
20000
24000
2016
2017
2018
2019
2020
2021
2022
Date

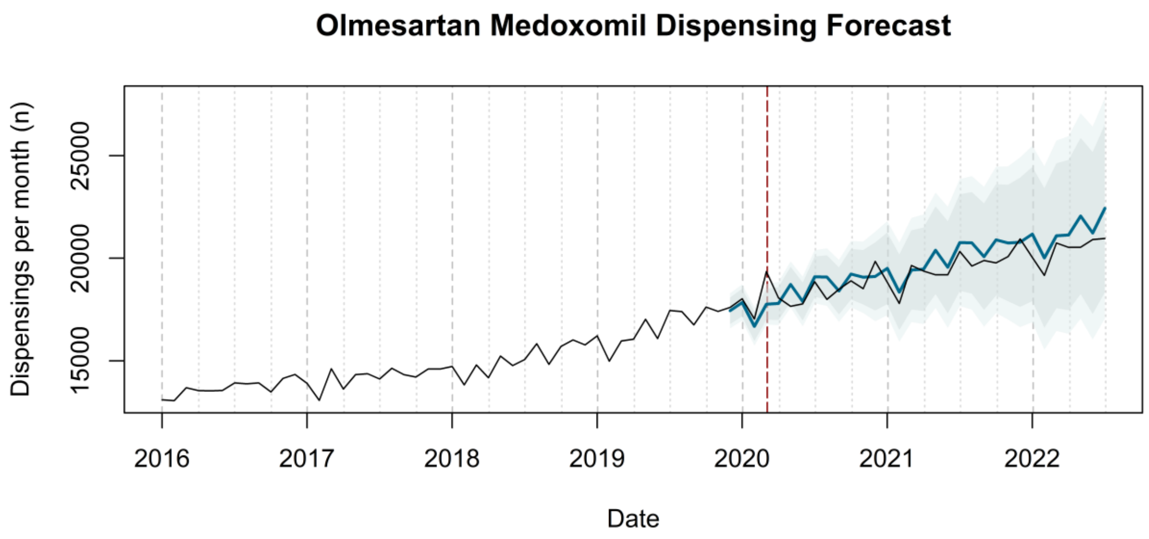
Olmesartan Medoxomil Dispensing Forecast
Dispensings per month (n)
15000
20000
25000
2016
2017
2018
2019
2020
2021
2022
Date

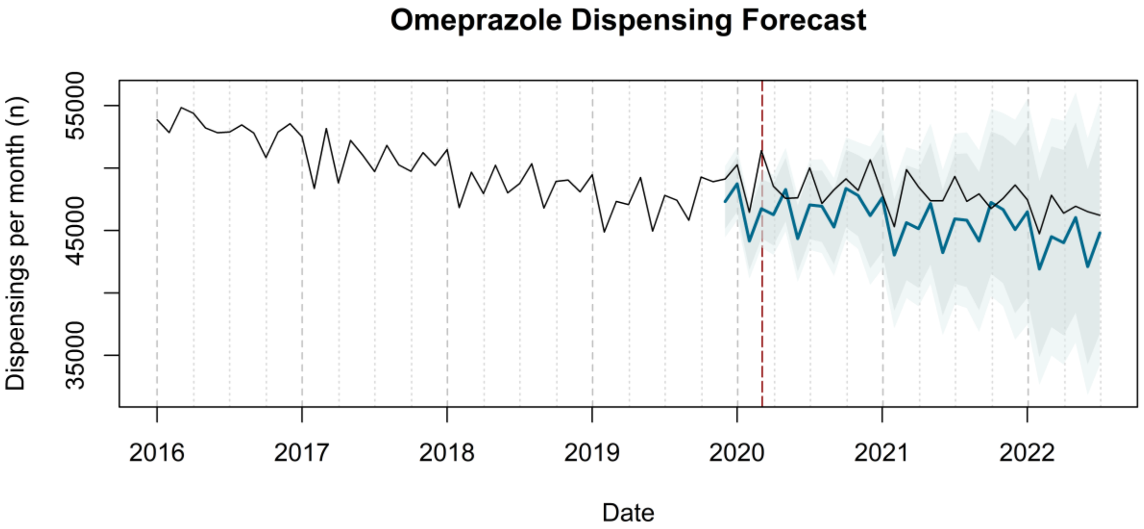
Omeprazole Dispensing Forecast
Dispensings per month (n)
35000
45000
55000
2016
2017
2018
2019
2020
2021
2022
Date

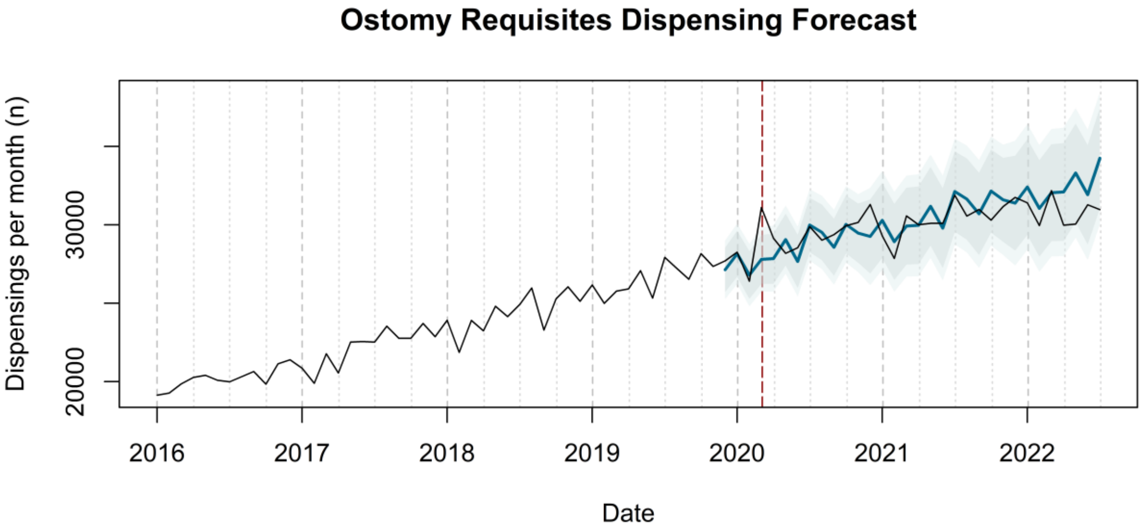
Ostomy Requisites Dispensing Forecast
Dispensings per month (n)
20000
30000
2016
2017
2018
2019
2020
2021
2022
Date

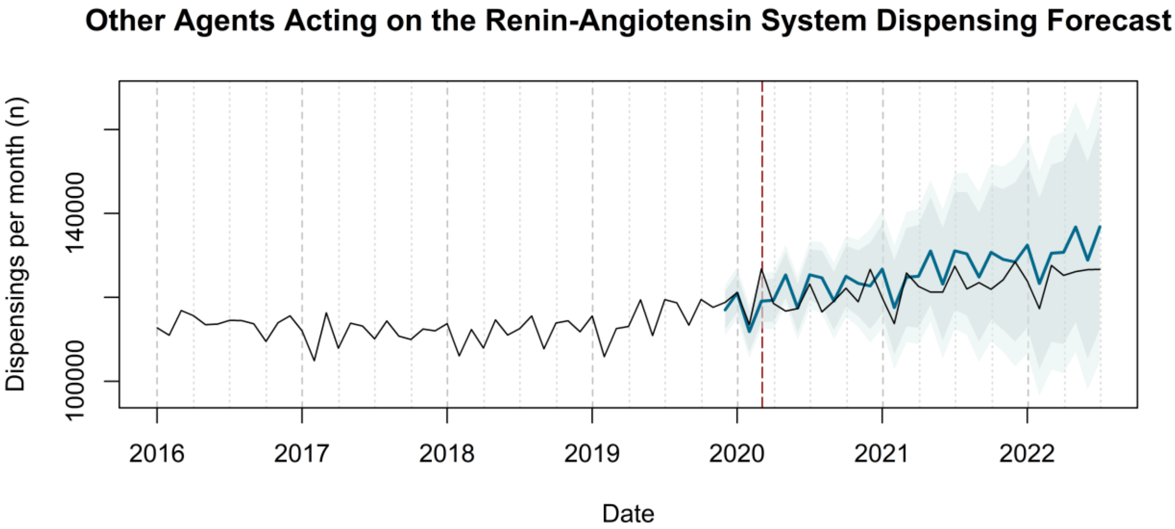
Other Agents Acting on the Renin-Angiotensin System Dispensing Forecast
Dispensings per month (n)
100000
140000
2016
2017
2018
2019
2020
2021
2022
Date

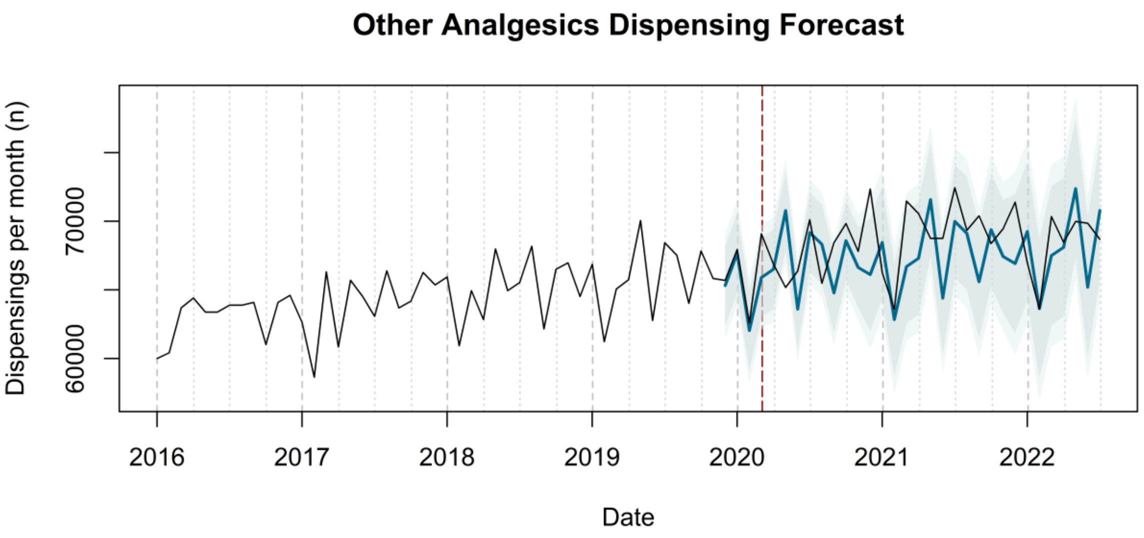

Other Analgesics Dispensing Forecast
Dispensings per month (n)
60000
70000
2016
2017
2018
2019
2020
2021
2022
Date


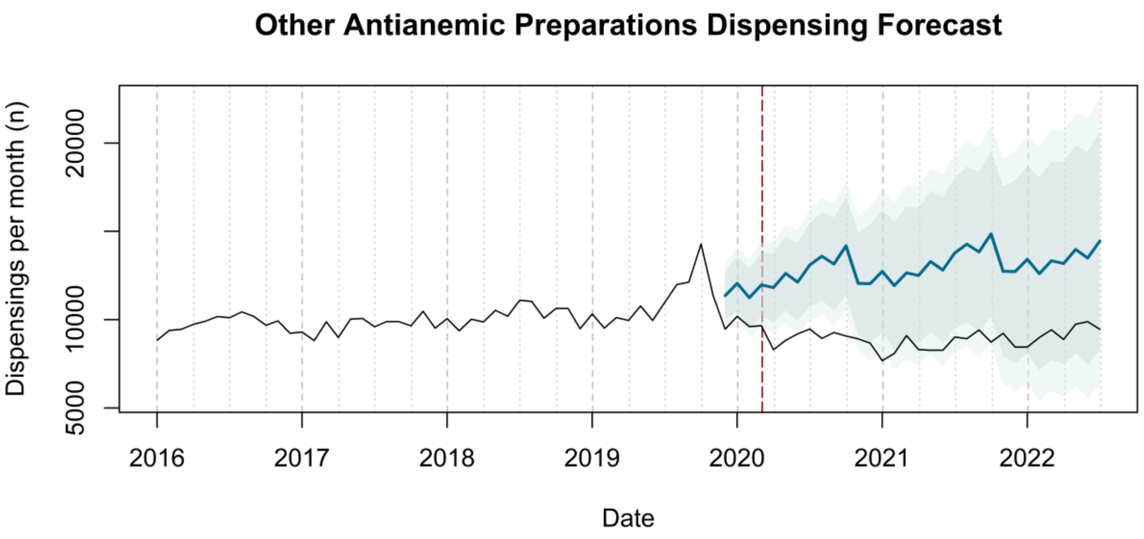

Other Antianemic Preparations Dispensing Forecast
Dispensings per month (n)
5000
10000
20000
2016
2017
2018
2019
2020
2021
2022
Date


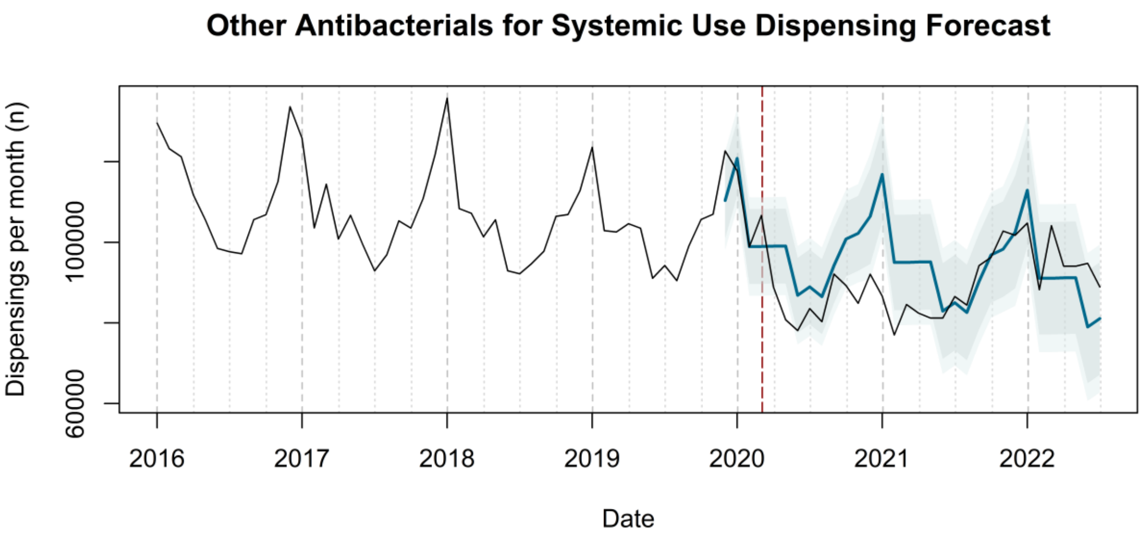

Other Antibacterials for Systemic Use Dispensing Forecast
Dispensings per month (n)
60000
100000
2016
2017
2018
2019
2020
2021
2022
Date

### Other Antiinflammatory and Antirheumatic Products Dispensing Forecast

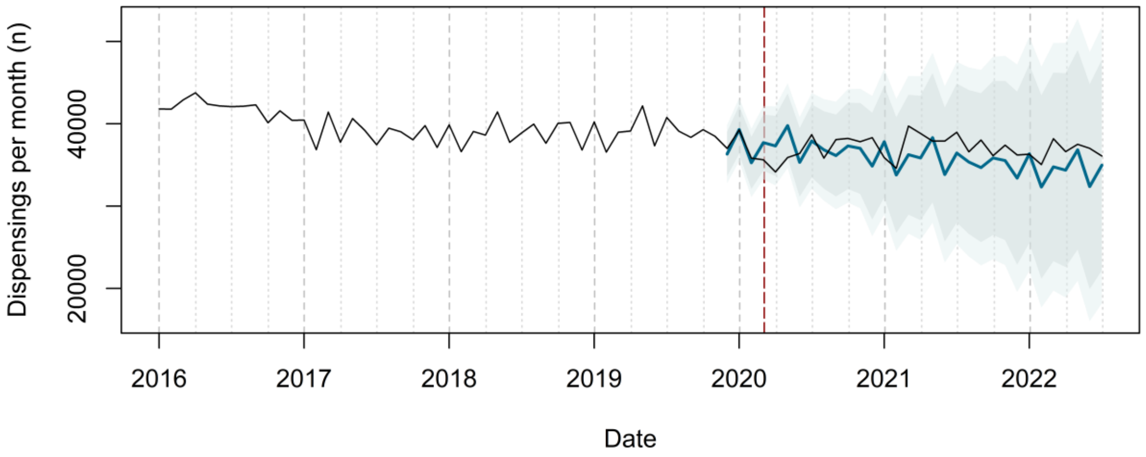


### Other Antithrombotic Agents Dispensing Forecast

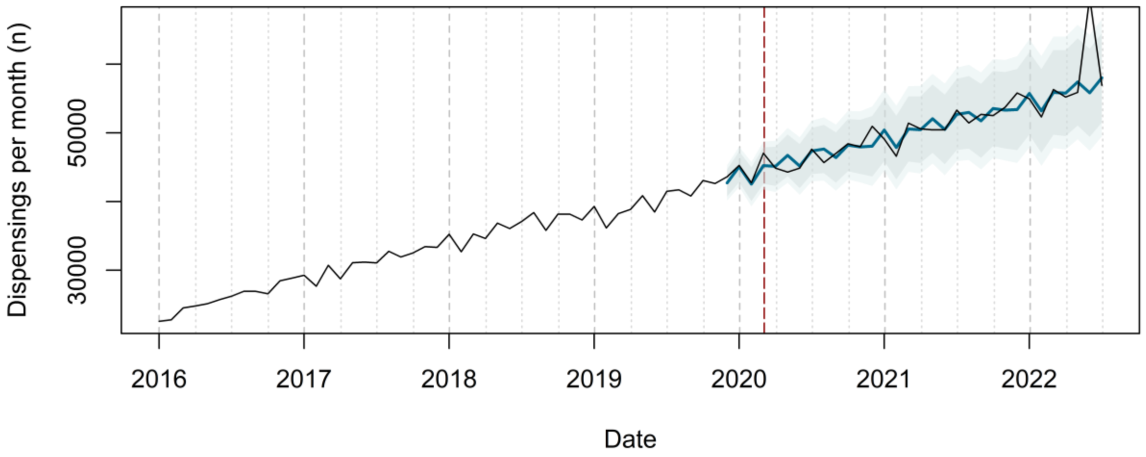


### Other Beta Blocking Agents Dispensing Forecast

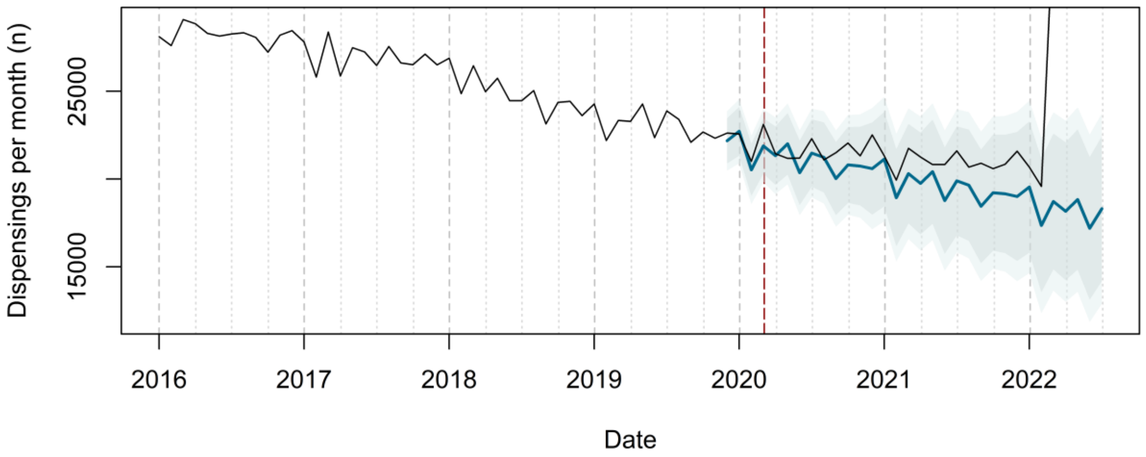

**Other Calcium Channel Blockers Dispensing Forecast**

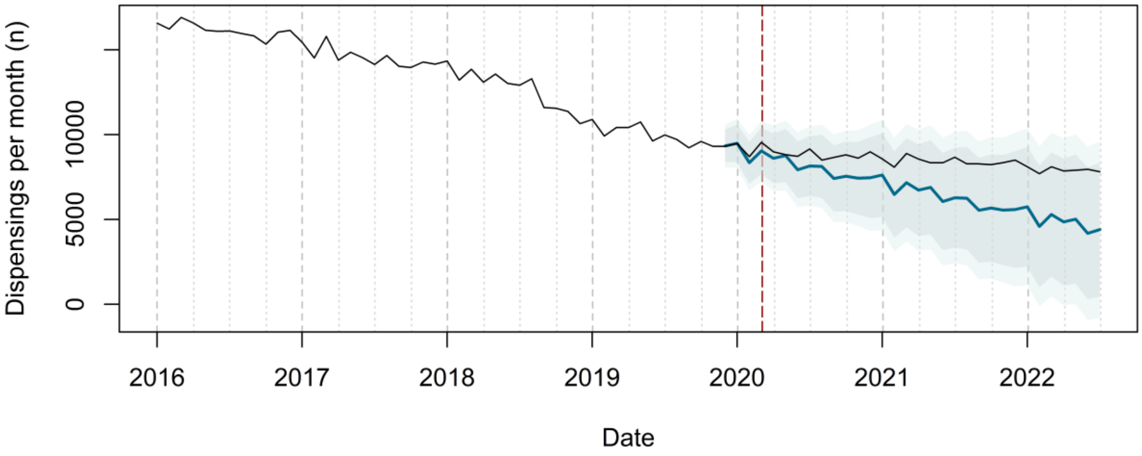


**Other Drugs for Acid Related Disorders Dispensing Forecast**

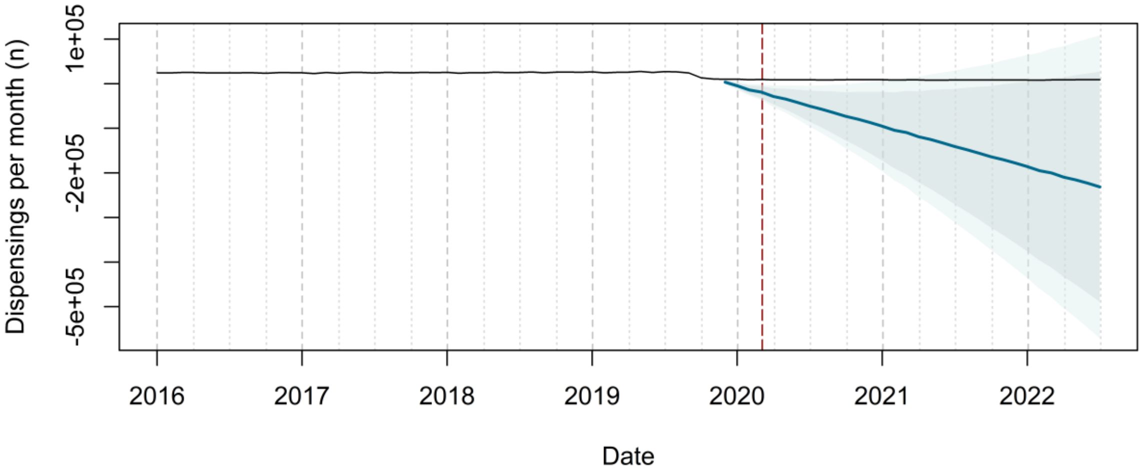


**Other Drugs for Obstructive Airway Diseases Dispensing Forecast**

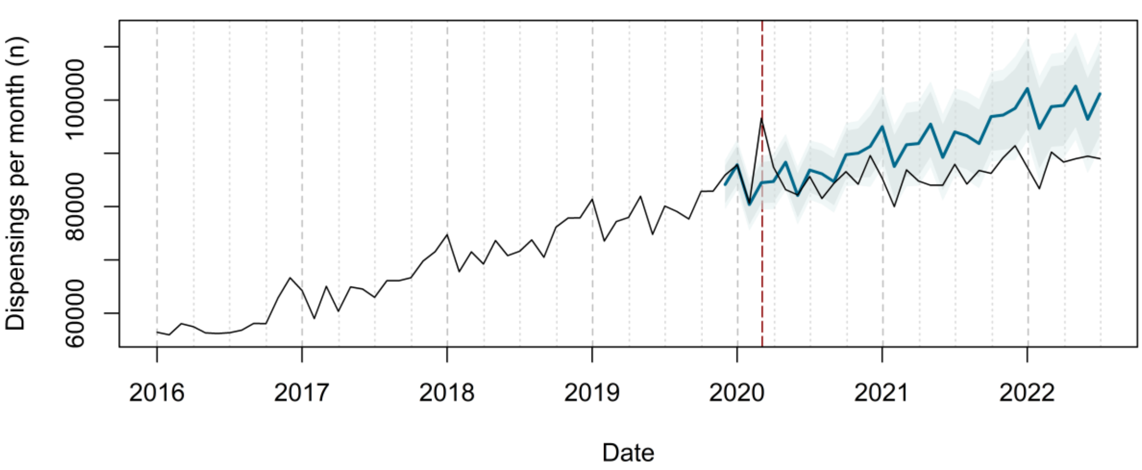

### Other Emollients and Protectives Dispensing Forecast

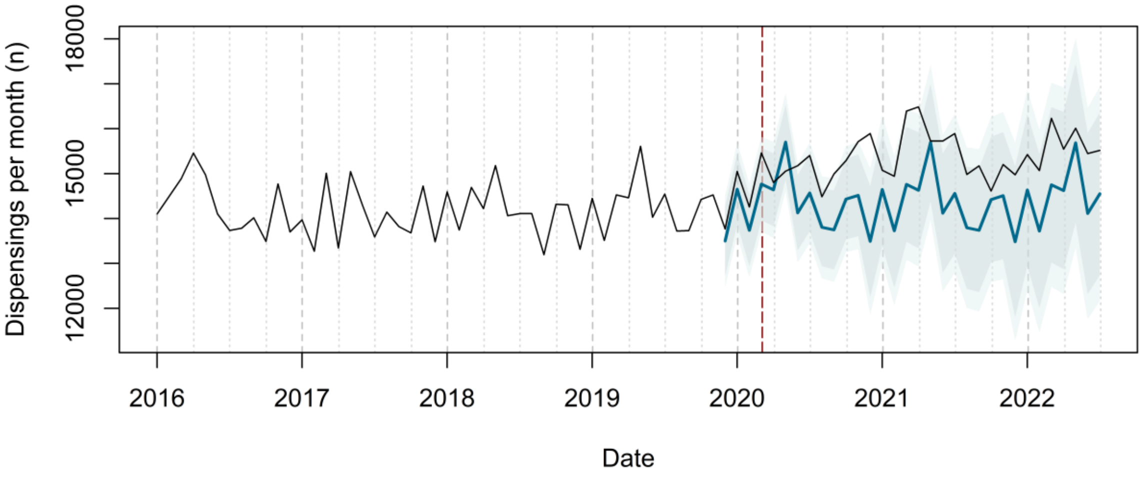


### Other Laxatives Dispensing Forecast

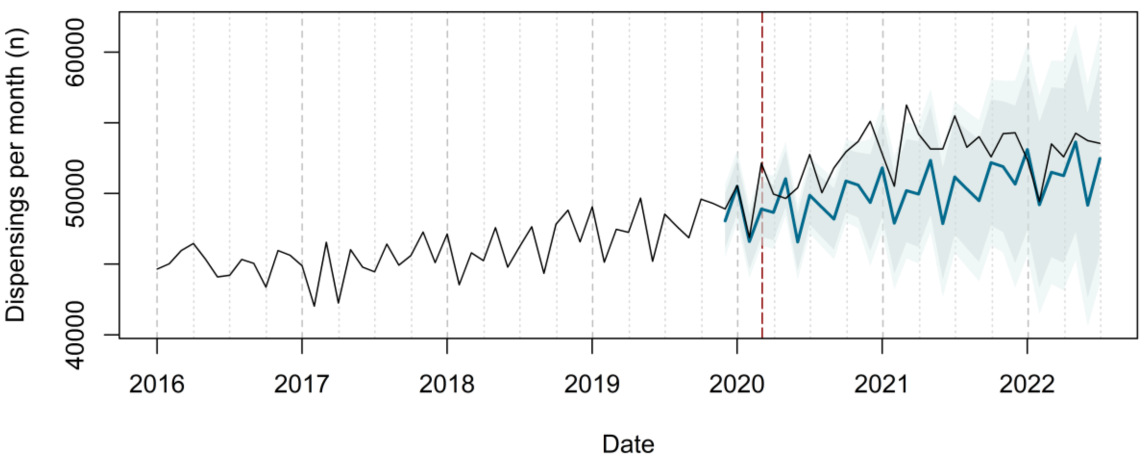


### Other Ophthalmologicals Dispensing Forecast

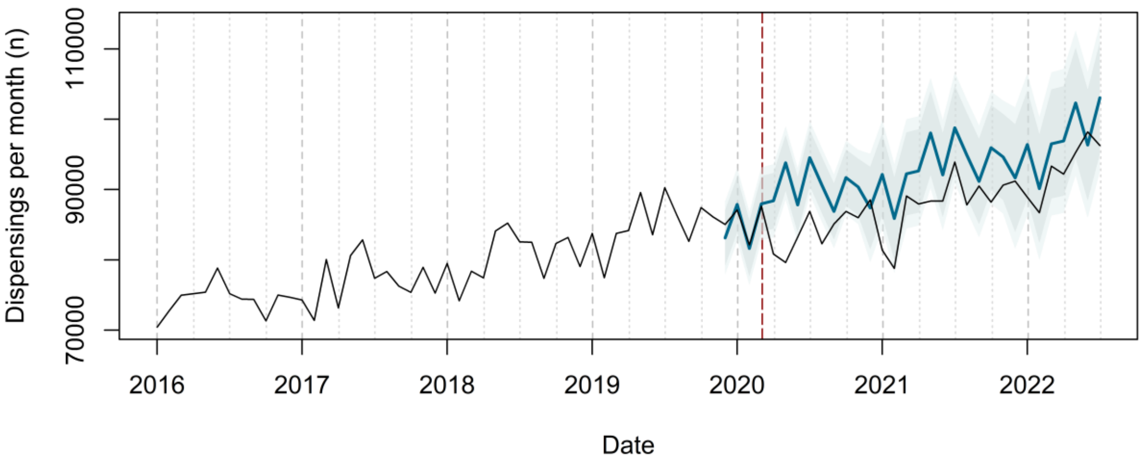

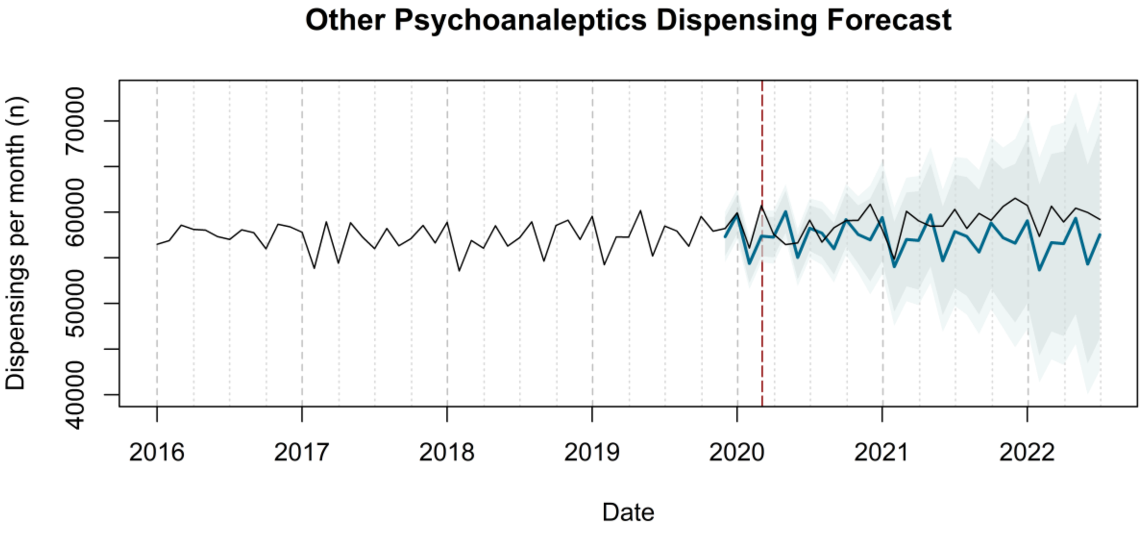
Other Psychoanaleptics Dispensing Forecast
Dispensings per month (n)
40000
50000
60000
70000
2016
2017
2018
2019
2020
2021
2022
Date

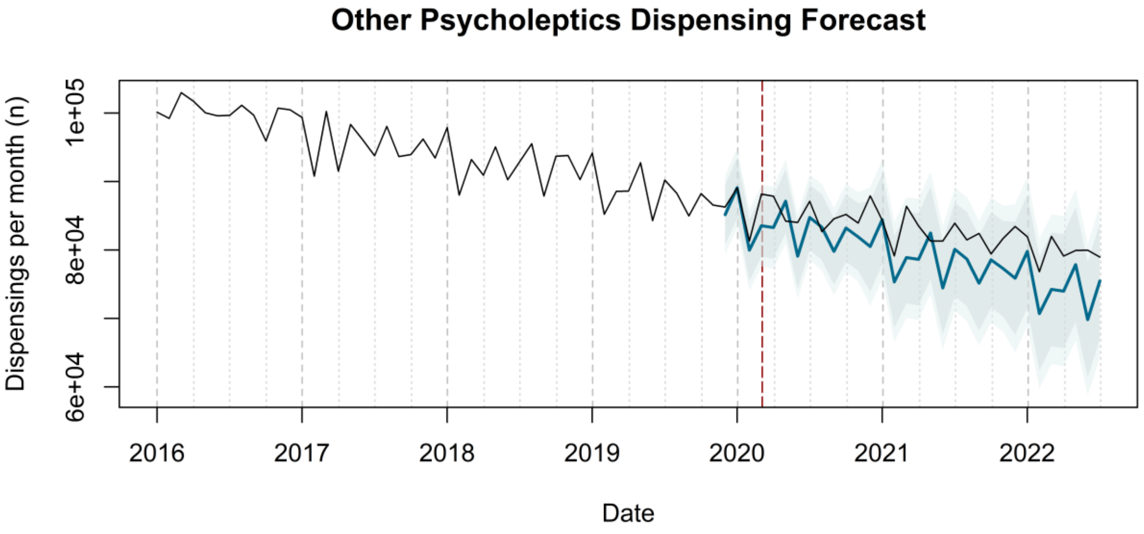
Other Psycholeptics Dispensing Forecast
Dispensings per month (n)
6e+04
8e+04
1e+05
2016
2017
2018
2019
2020
2021
2022
Date

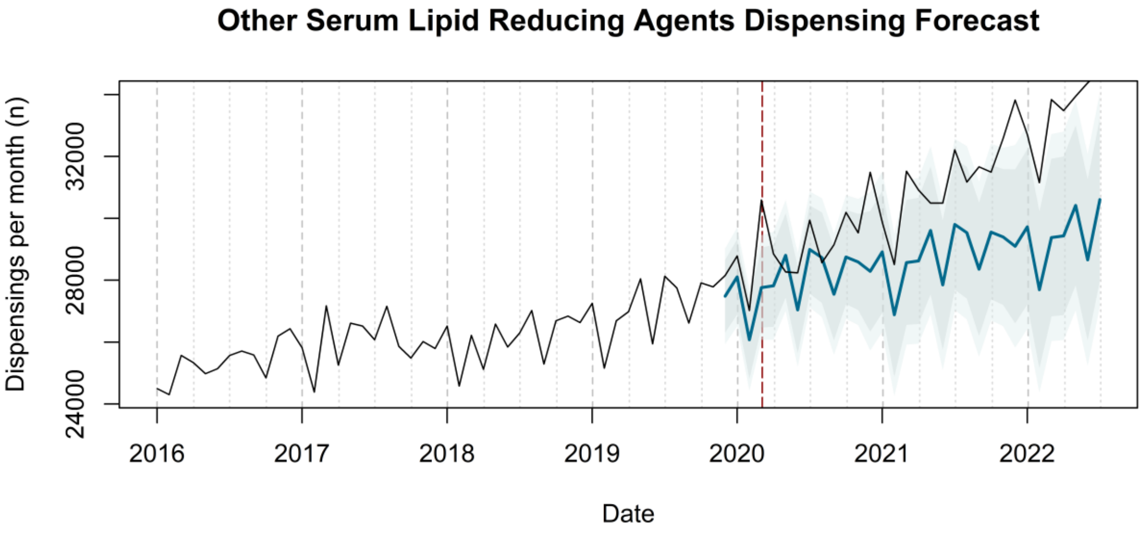
Other Serum Lipid Reducing Agents Dispensing Forecast
Dispensings per month (n)
24000
28000
32000
2016
2017
2018
2019
2020
2021
2022
Date

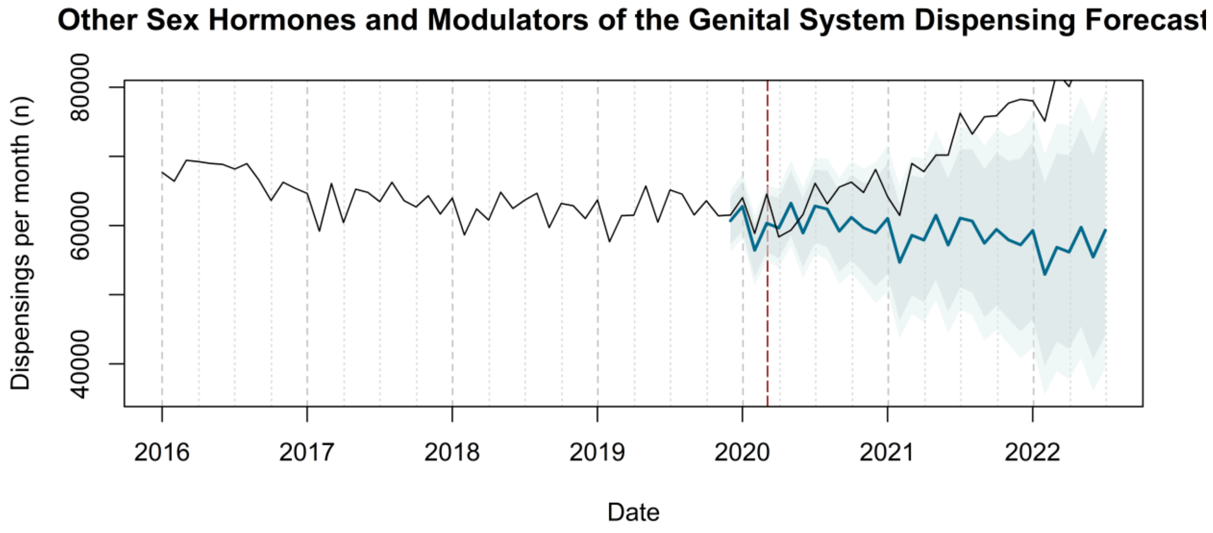

Other Sex Hormones and Modulators of the Genital System Dispensing Forecast
Dispensings per month (n)
40000
60000
80000
2016
2017
2018
2019
2020
2021
2022
Date


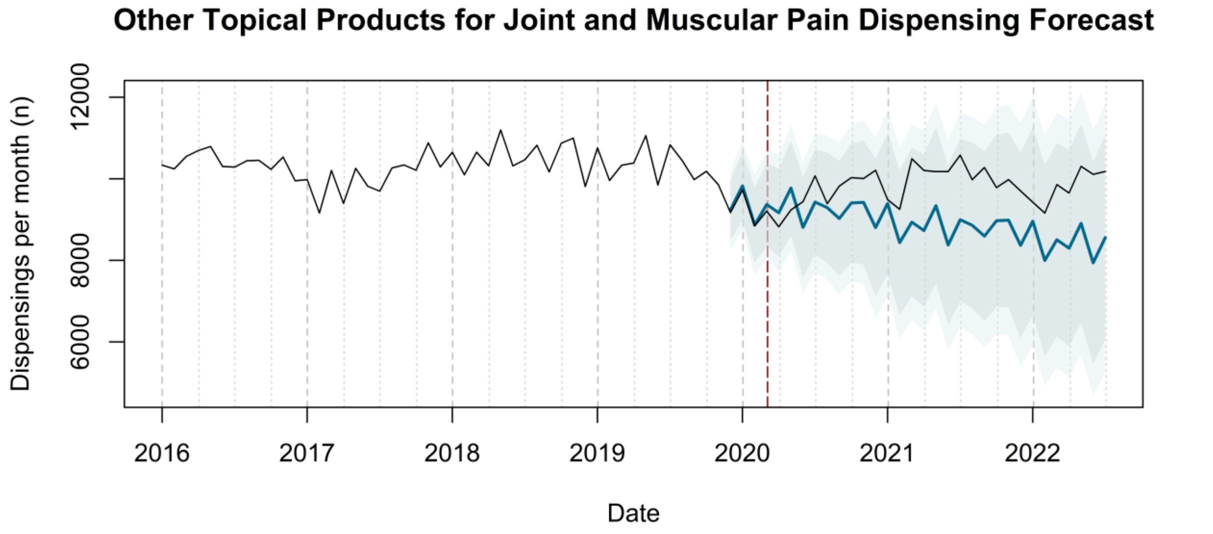

Other Topical Products for Joint and Muscular Pain Dispensing Forecast
Dispensings per month (n)
6000
8000
12000
2016
2017
2018
2019
2020
2021
2022
Date


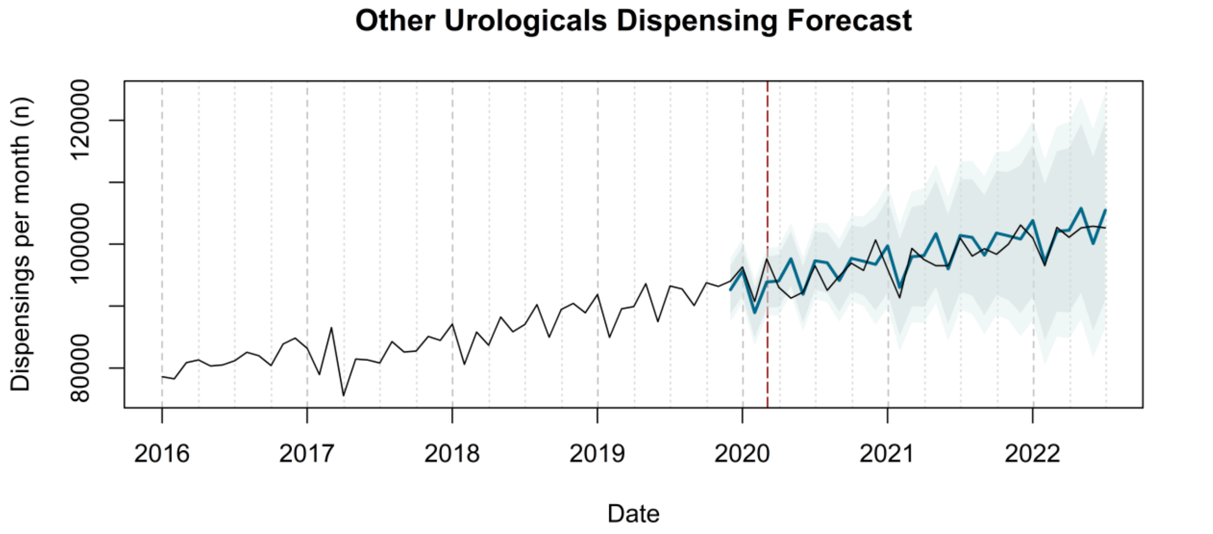

Other Urologicals Dispensing Forecast
Dispensings per month (n)
80000
100000
120000
2016
2017
2018
2019
2020
2021
2022
Date

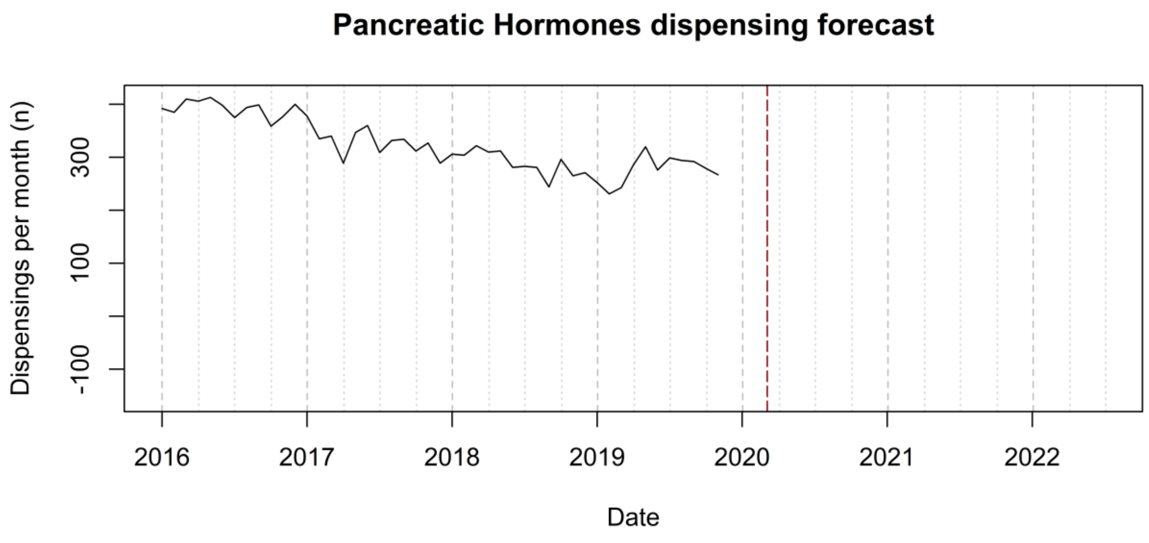

Pancreatic Hormones dispensing forecast
Dispensings per month (n)
-100
100
300
2016
2017
2018
2019
2020
2021
2022
Date


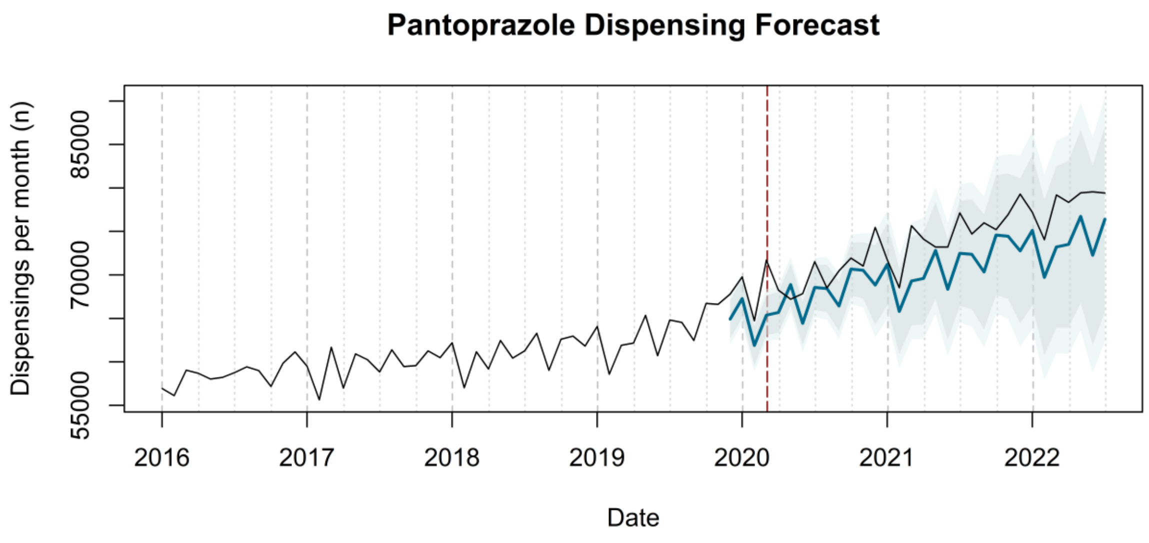

Pantoprazole Dispensing Forecast
Dispensings per month (n)
55000
70000
85000
2016
2017
2018
2019
2020
2021
2022
Date


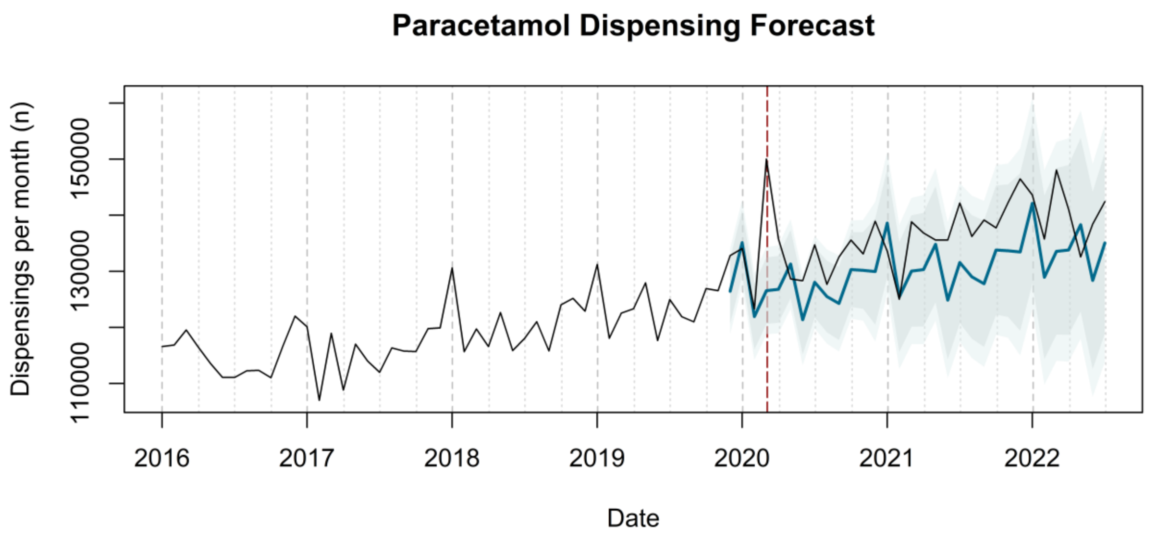

Paracetamol Dispensing Forecast
Dispensings per month (n)
110000
130000
150000
2016
2017
2018
2019
2020
2021
2022
Date

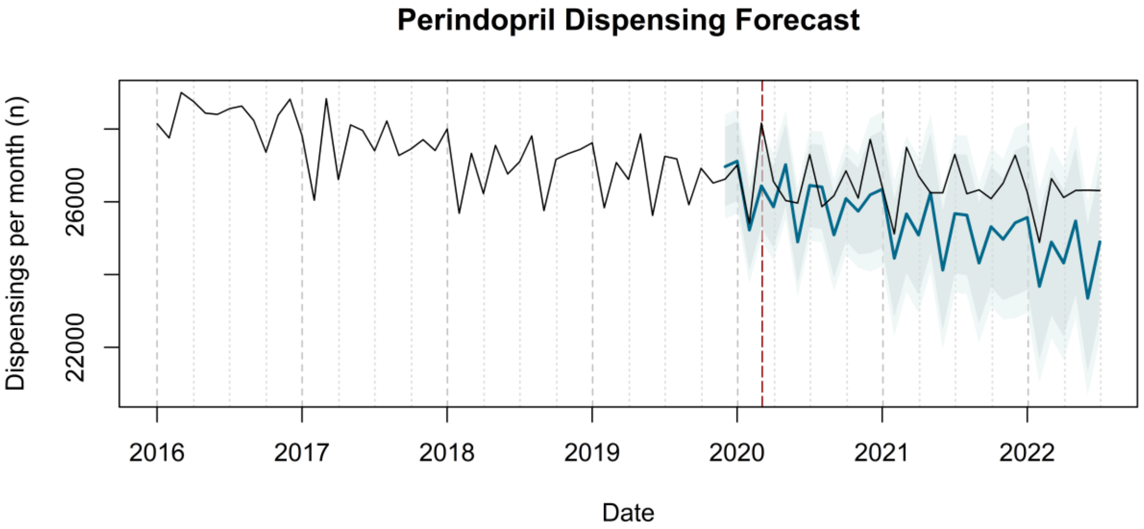
Perindopril Dispensing Forecast
Dispensings per month (n)
22000
26000
2016
2017
2018
2019
2020
2021
2022
Date

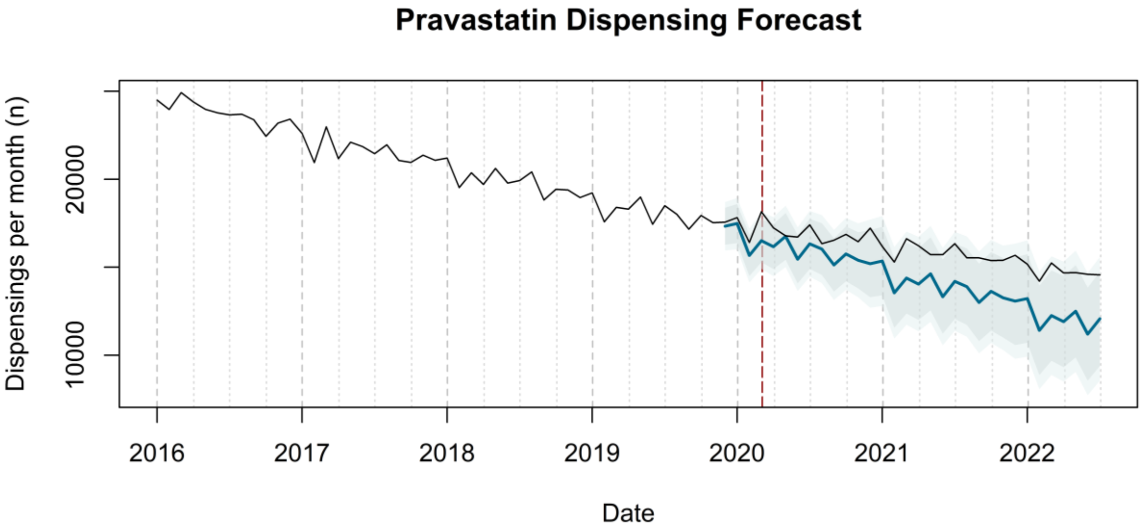
Pravastatin Dispensing Forecast
Dispensings per month (n)
10000
20000
2016
2017
2018
2019
2020
2021
2022
Date

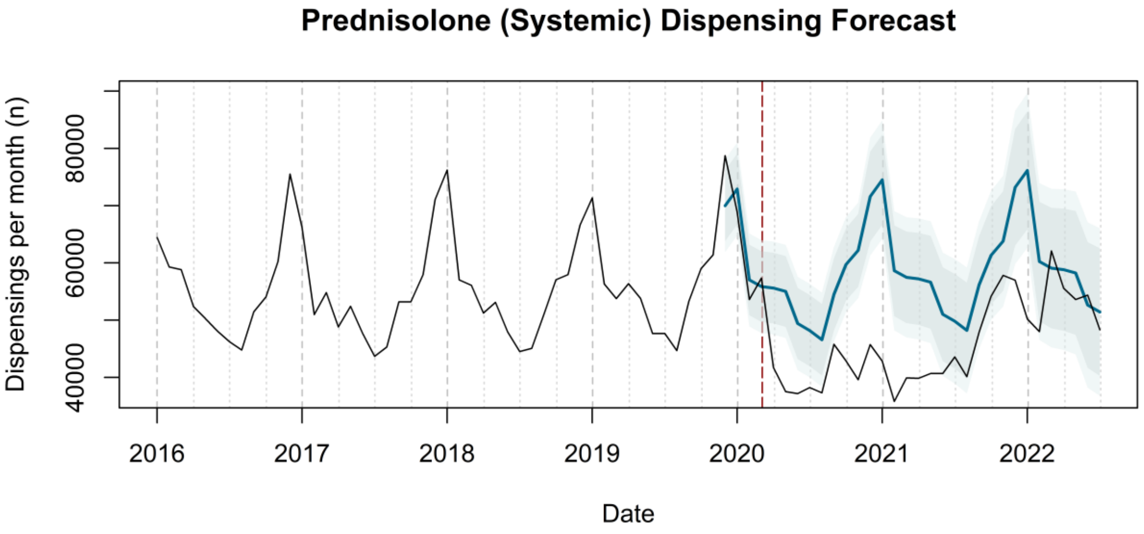
Prednisolone (Systemic) Dispensing Forecast
Dispensings per month (n)
40000
60000
80000
2016
2017
2018
2019
2020
2021
2022
Date

**Pregabalin Dispensing Forecast**

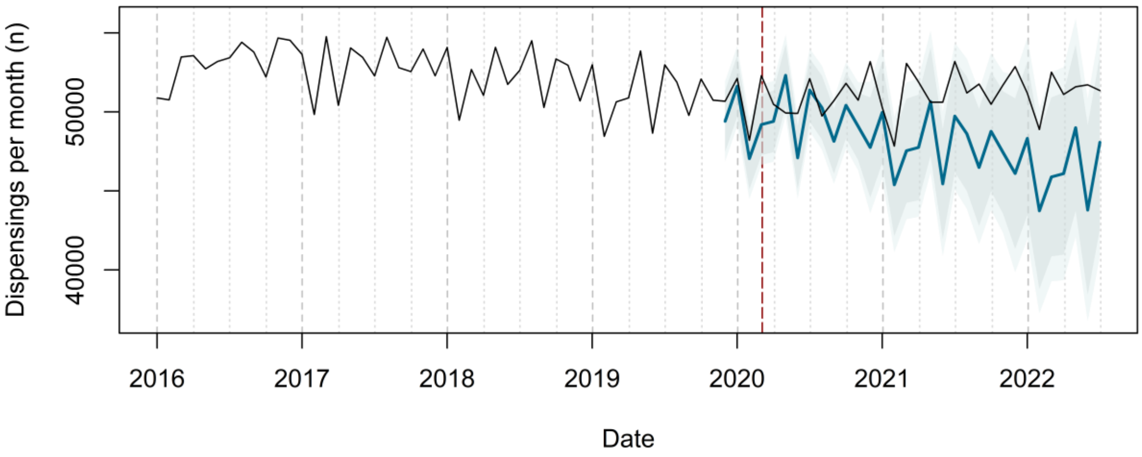


**Quetiapine Dispensing Forecast**

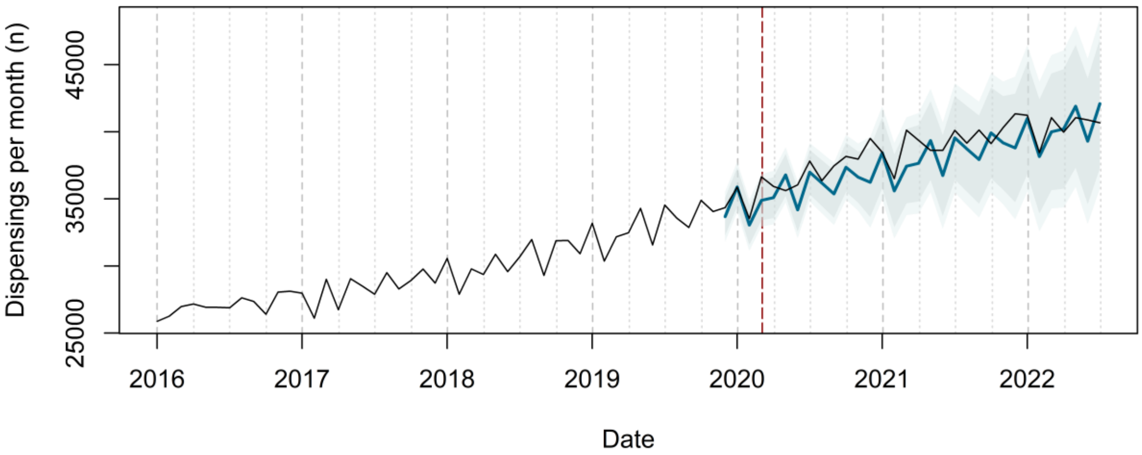


**Ramipril Dispensing Forecast**

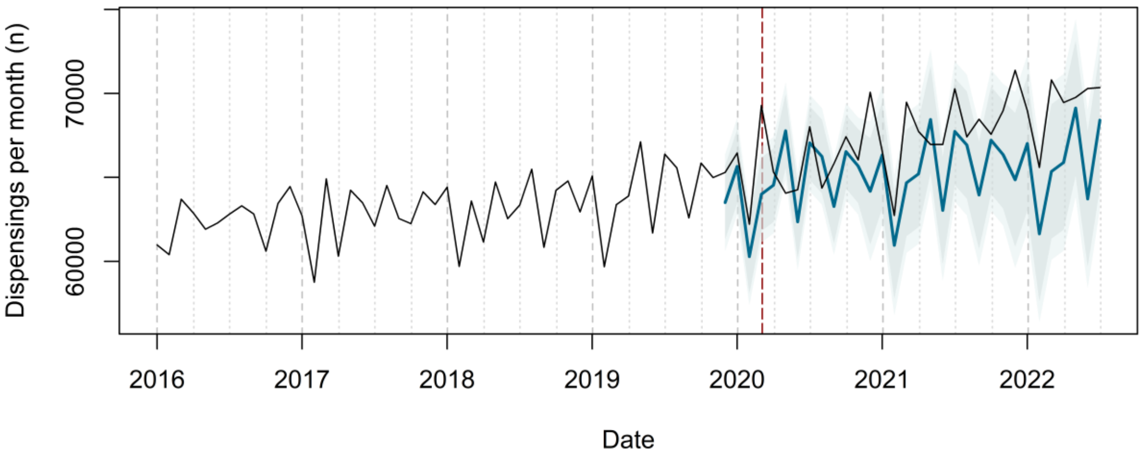

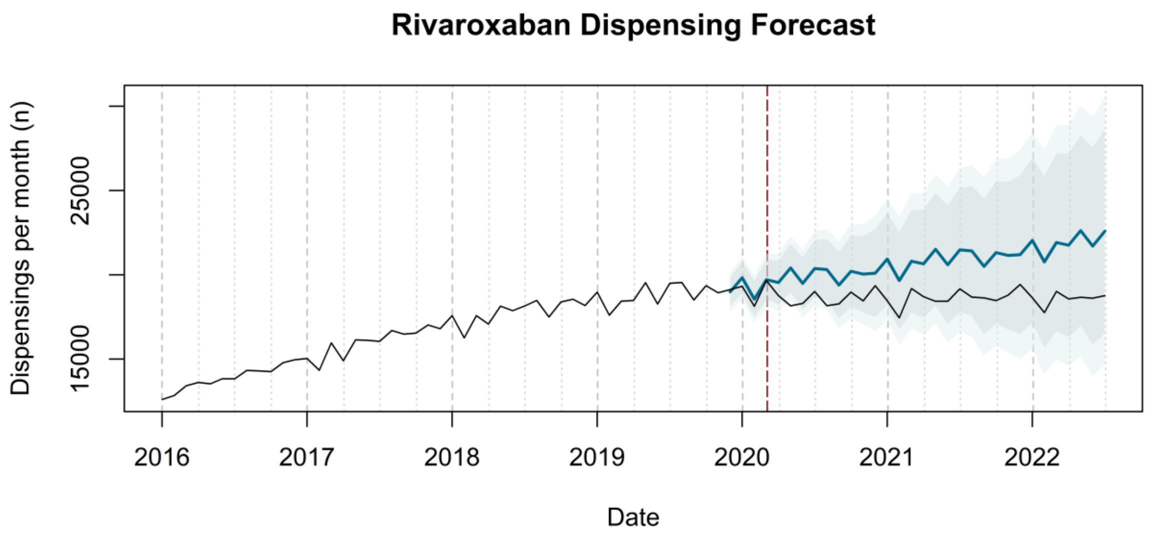
Rivaroxaban Dispensing Forecast
Dispensings per month (n)
15000
25000
2016
2017
2018
2019
2020
2021
2022
Date

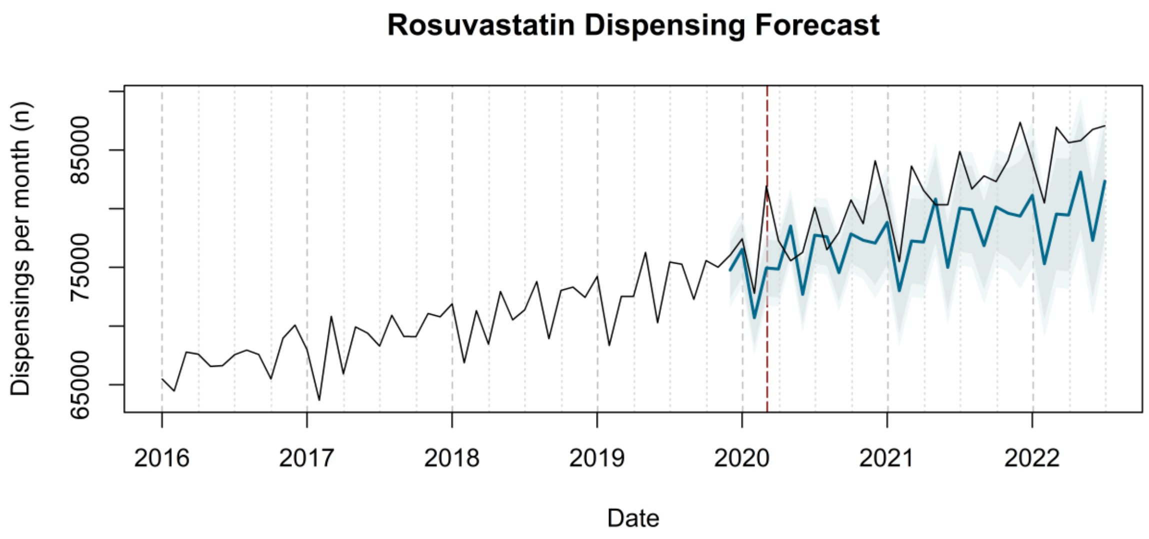
Rosuvastatin Dispensing Forecast
Dispensings per month (n)
65000
75000
85000
2016
2017
2018
2019
2020
2021
2022
Date

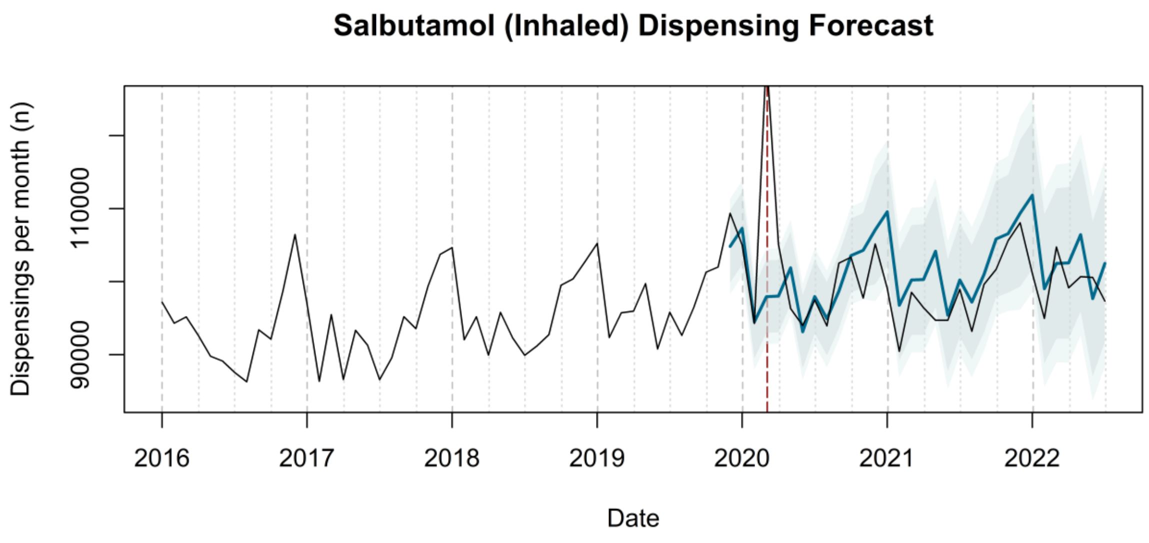
Salbutamol (Inhaled) Dispensing Forecast
Dispensings per month (n)
90000
110000
2016
2017
2018
2019
2020
2021
2022
Date

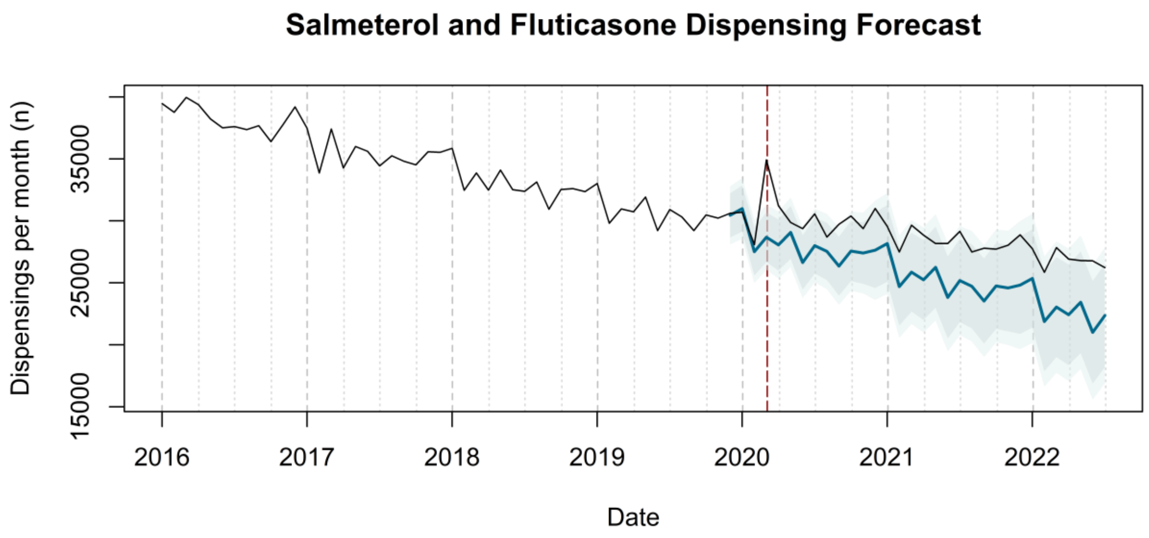
Salmeterol and Fluticasone Dispensing Forecast
Dispensings per month (n)
15000
25000
35000
2016
2017
2018
2019
2020
2021
2022
Date

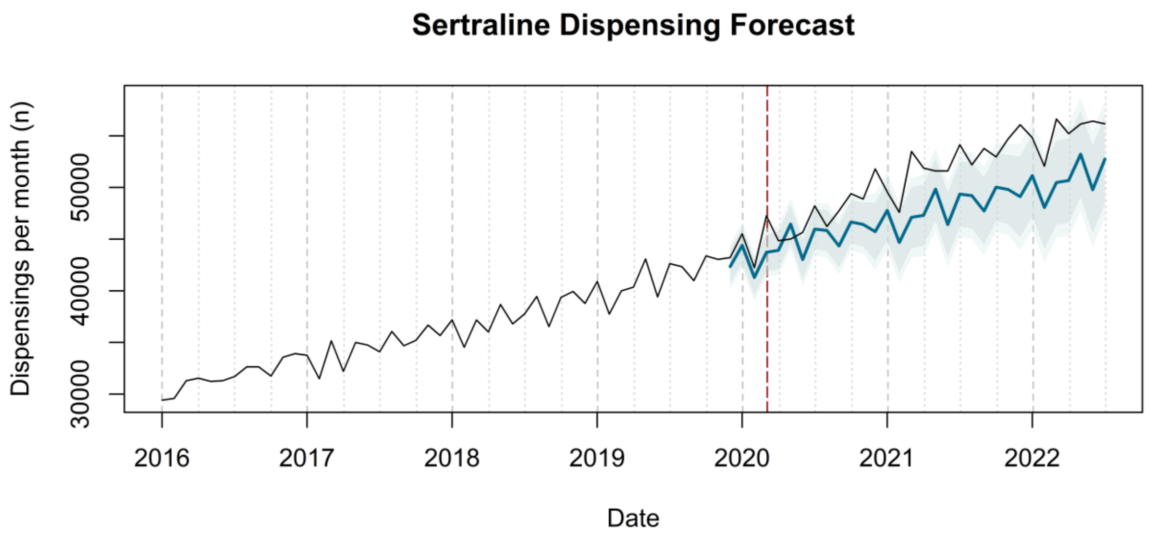
Sertraline Dispensing Forecast
Dispensings per month (n)
30000
40000
50000
2016
2017
2018
2019
2020
2021
2022
Date

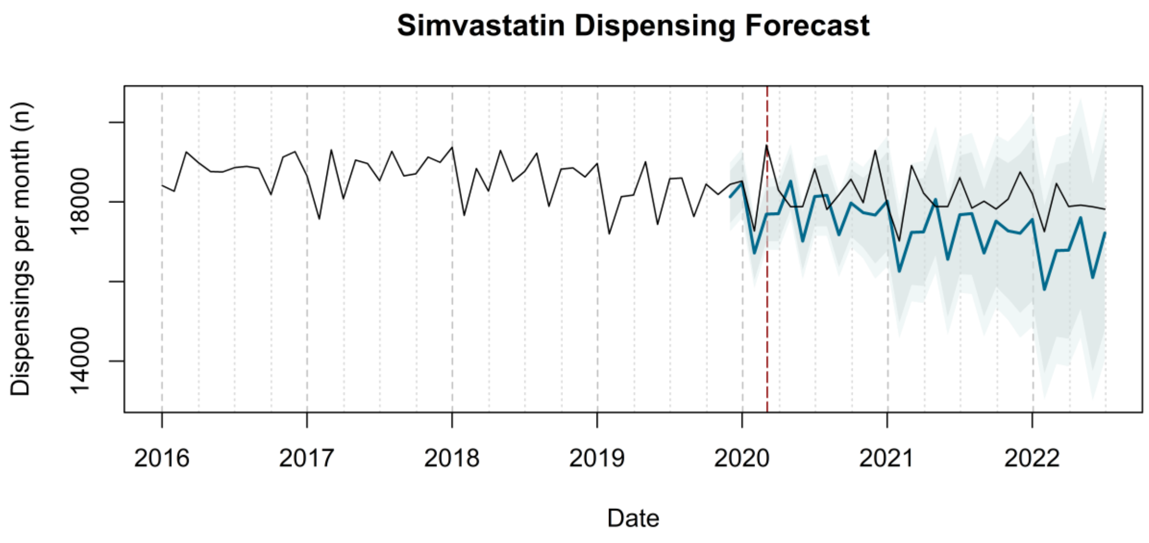
Simvastatin Dispensing Forecast
Dispensings per month (n)
14000
18000
2016
2017
2018
2019
2020
2021
2022
Date

### Tamsulosin Dispensing Forecast

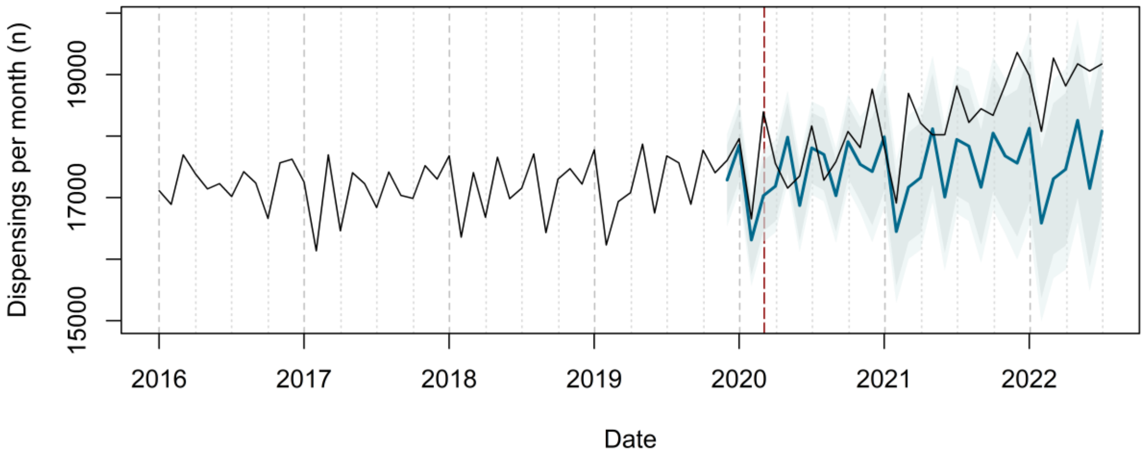


### Tiotropium Bromide Dispensing Forecast

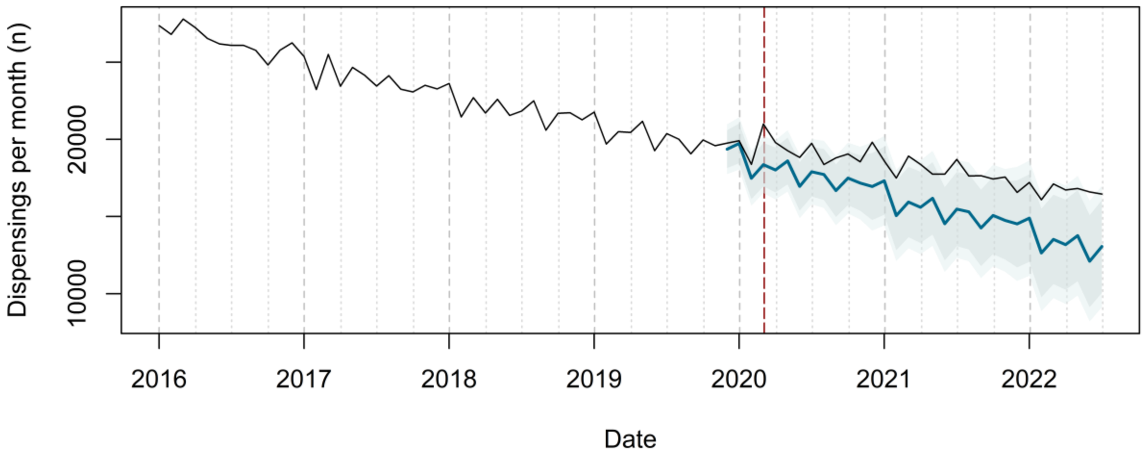


### Tramadol Dispensing Forecast

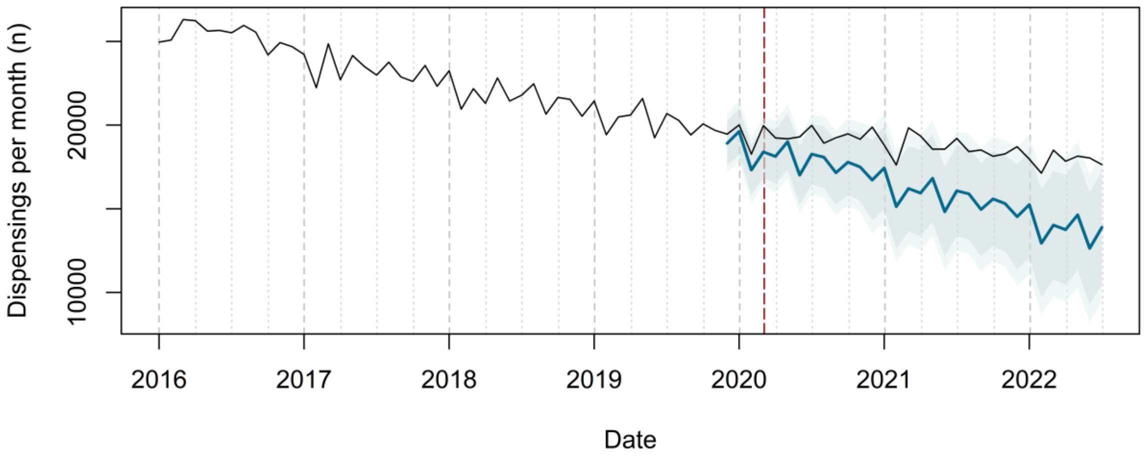

### Valsartan Dispensing Forecast

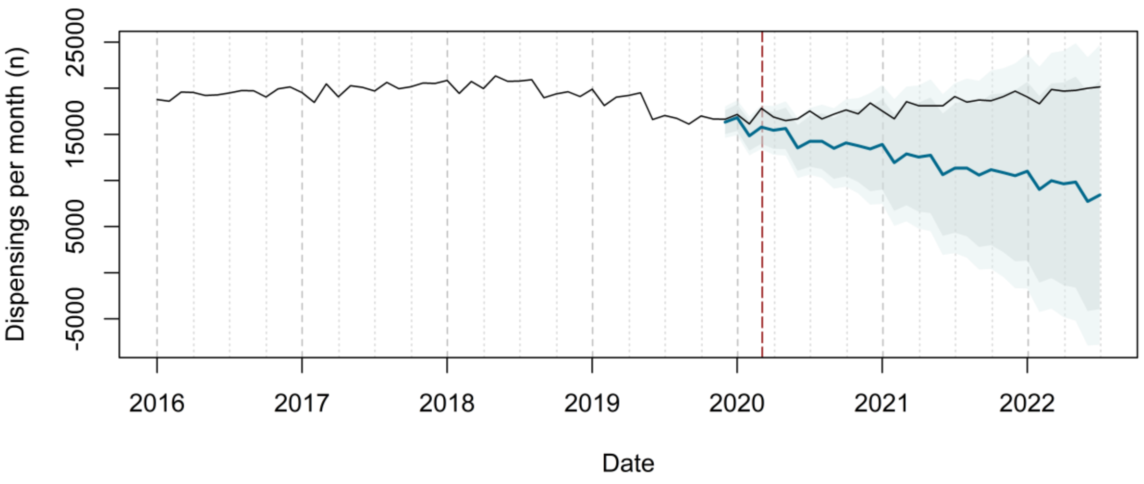


### Venlafaxine Dispensing Forecast

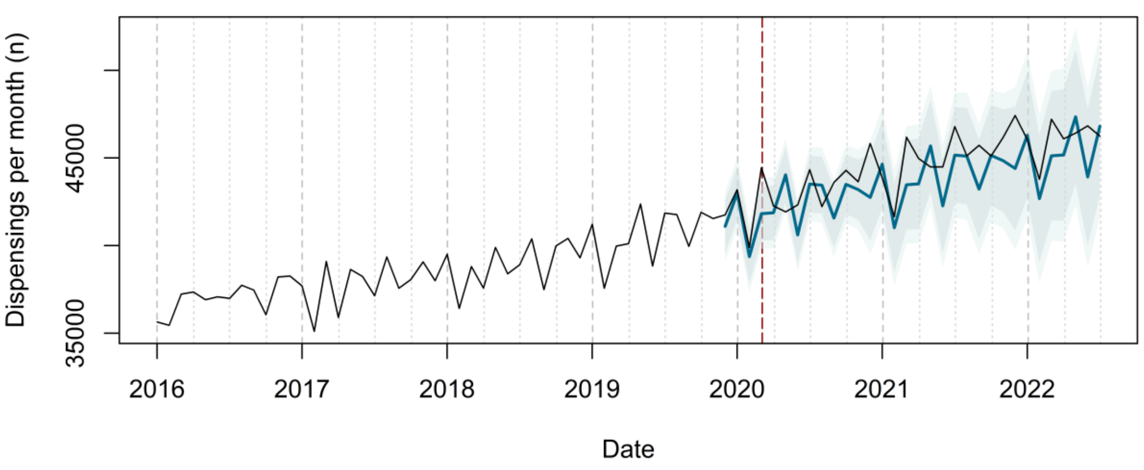


### Warfarin Dispensing Forecast

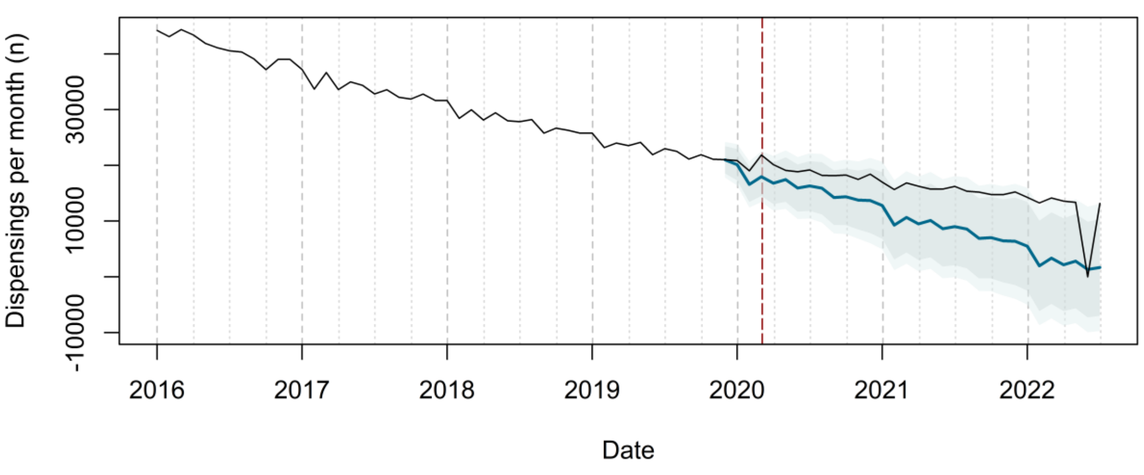

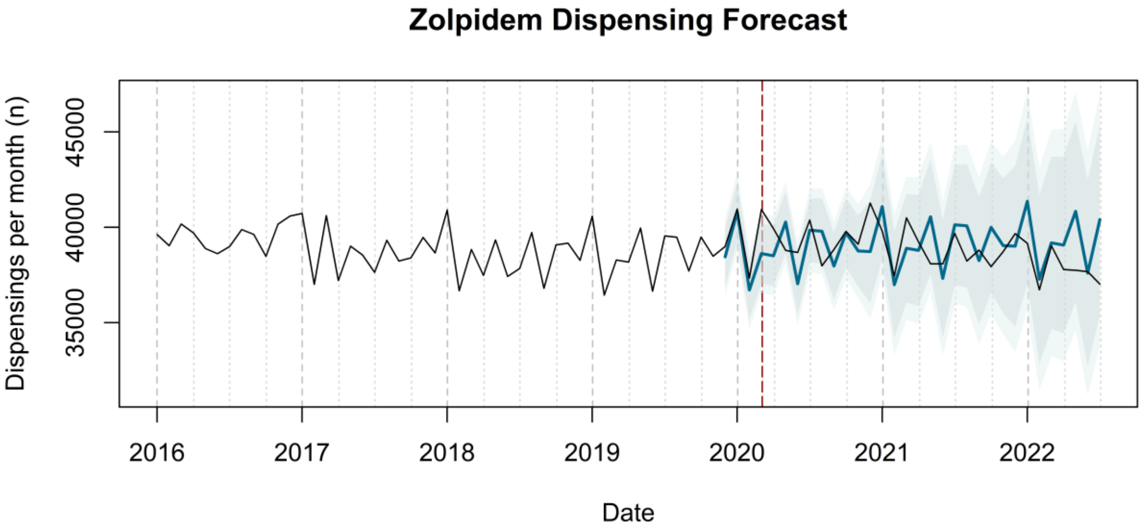

Zolpidem Dispensing Forecast
Dispensings per month (n)
35000
40000
45000
2016
2017
2018
2019
2020
2021
2022
Date


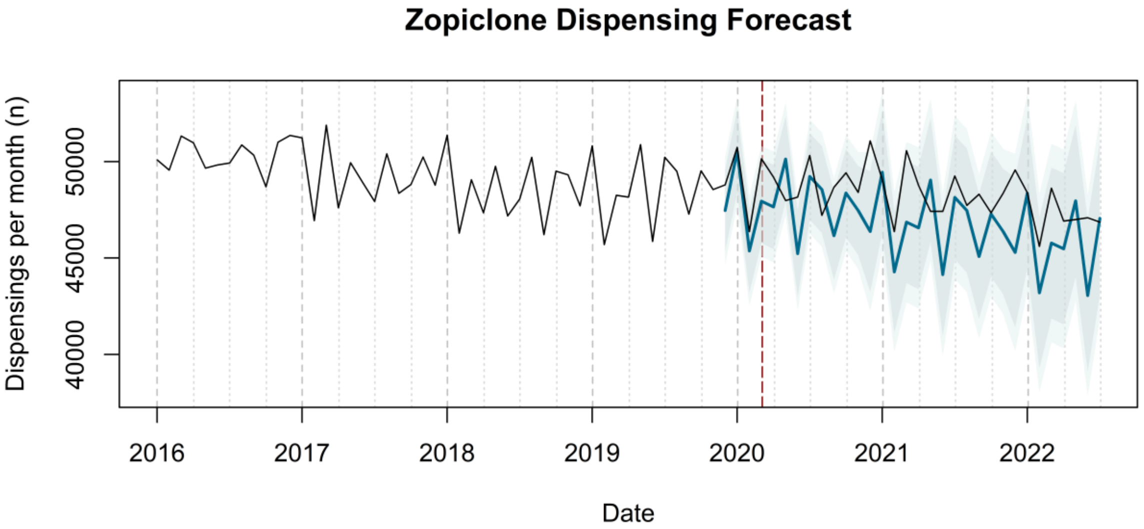

Zopiclone Dispensing Forecast
Dispensings per month (n)
40000
45000
50000
2016
2017
2018
2019
2020
2021
2022
Date
